# The shared genetic risk architecture of neurological and psychiatric disorders: a genome-wide analysis

**DOI:** 10.1101/2023.07.21.23292993

**Authors:** Olav B. Smeland, Gleda Kutrolli, Shahram Bahrami, Vera Fominykh, Nadine Parker, Guy F. L. Hindley, Linn Rødevand, Piotr Jaholkowski, Markos Tesfaye, Pravesh Parekh, Torbjørn Elvsåshagen, Andrew D. Grotzinger, The International Multiple Sclerosis Genetics Consortium (IMSGC), The International Headache Genetics Consortium (IHGC), Nils Eiel Steen, Dennis van der Meer, Kevin S. O’Connell, Srdjan Djurovic, Anders M. Dale, Alexey A. Shadrin, Oleksandr Frei, Ole A. Andreassen

**Affiliations:** NORMENT, Division of Mental Health and Addiction, Oslo University Hospital & Institute of Clinical Medicine, University of Oslo, Oslo, Norway; Psychosis Studies, Institute of Psychiatry, Psychology and Neurosciences, King’s College London, London, United Kingdom; Department of Psychiatry, St. Paul’s Hospital Millennium Medical College, Addis Ababa, Ethiopia; Department of Neurology, Division of Clinical Neuroscience, Oslo University Hospital, Oslo, Norway; Department of Psychology and Neuroscience, University of Colorado at Boulder, Boulder, CO, USA; Institute for Behavioral Genetics, University of Colorado at Boulder, Boulder, CO, USA; School of Mental Health and Neuroscience, Faculty of Health, Medicine and Life Sciences, Maastricht University, The Netherlands; Department of Medical Genetics, Oslo University Hospital, Oslo, Norway; NORMENT, Department of Clinical Science, University of Bergen, Bergen, Norway; Multimodal Imaging Laboratory, University of California San Diego, La Jolla, USA; Department of Psychiatry, University of California, San Diego, La Jolla, USA; Department of Neurosciences, University of California San Diego, La Jolla, USA; Department of Radiology, University of California, San Diego, La Jolla, USA; KG Jebsen Centre for Neurodevelopmental disorders, University of Oslo, Oslo, Norway; Center for Bioinformatics, Department of Informatics, University of Oslo, Oslo, Norway

## Abstract

While neurological and psychiatric disorders have historically been considered to reflect distinct pathogenic entities, recent findings suggest shared pathobiological mechanisms. However, the extent to which these heritable disorders share genetic influences remains unclear. Here, we performed a comprehensive analysis of GWAS data, involving nearly 1 million cases across ten neurological diseases and ten psychiatric disorders, to compare their common genetic risk and biological underpinnings. Using complementary statistical tools, we demonstrate widespread genetic overlap across the disorders, even in the absence of genetic correlations. This indicates that a large set of common variants impact risk of multiple neurological and psychiatric disorders, but with divergent effect sizes. Furthermore, biological interrogation revealed a range of biological processes associated with neurological diseases, while psychiatric disorders consistently implicated neuronal biology. Altogether, the study indicates that neurological and psychiatric disorders share key etiological aspects, which has important implications for disease classification, precision medicine, and clinical practice.

## Introduction

Neurological and psychiatric disorders rank among the leading causes of disability and mortality worldwide^1^. Despite their shared link to the nervous system, the disorders have generally been considered to reflect distinct pathogenic entities, as emphasized by their separate classification in the International Classification of Diseases^2^. The clinical division was driven by progress in brain research during the 19^th^ and 20^th^ century^3,4^. While neurology laid claim on the disorders with demonstrable neuropathology, such as Alzheimer’s disease (ALZ), psychiatry focused on the mental disorders without recognizable pathology, such as schizophrenia (SCZ). However, findings in neuroscience over the past decades, combined with clinical and epidemiological observations, have challenged the validity of this clinical distinction^3–8^. Various therapeutic interventions are effective in both groups of disorders, for example transcranial magnetic stimulation in Parkinson’s disease (PD) and depression^9^ and anticonvulsants in epilepsy and bipolar disorder (BD)^10^. Neurological and psychiatric disorders also share clinical features, notably cognitive impairment, a key functional determinant^2,11^. Additionally, debilitating psychiatric symptoms such as hallucinations, delusions and mood disturbances are prominent across neurological diseases^12–14^, while classical neurological symptoms such as movement abnormalities are observed in psychiatric disorders^15^. Moreover, environmental risk factors such as pollutants increase risk of both neurological and psychiatric illnesses^8^, and epidemiological studies demonstrate high comorbidity between several neurological and psychiatric disorders^14,16–18^, including a higher incidence of dementia among individuals with psychotic disorders^18^. Furthermore, *in vivo* neuroimaging^19^ and postmortem^20^ investigations report systematic brain abnormalities in psychiatric disorders, indicating that mental disorders have a neural basis akin to neurological disease. Altogether, the existing clinical dichotomy inadequately reflects the interconnected nature of neurological and psychiatric disorders, emphasizing the need for a more unified clinical approach^3–8^. However, the extent to which these conditions share an etiological basis still remains largely unclear.

The significant heritability of neurological and psychiatric disorders indicates that genomic research could provide new insights into their etiology^21^. This could bridge the nosological gap by forming the basis for an etiology-driven approach to disease classification, reveal novel treatment targets, and inform the development of precision medicine approaches. In recent years, genome-wide association studies (GWAS) have identified multiple common genetic variants for neurological and psychiatric disorders. Two key findings have emerged: the conditions are polygenic and genetic overlap is ubiquitous^22,23^. Genetic overlap has mainly been assessed by estimating pairwise genetic correlations using tools such as linkage disequilibrium (LD) score regression (LDSC)^24^, demonstrating that the genetic risk of psychiatric disorders is highly intercorrelated^25–28^. On the contrary, there are fewer pairwise genetic correlations among neurological diseases^25,29,30^ and between neurological and psychiatric disorders^25,31^. Accordingly, neurological diseases have been considered to be genetically disparate from psychiatric disorders^25^, in line with their clinical distinction^2^. However, estimates of genetic correlation are sensitive to low GWAS power and do not provide a complete picture of the genetic relationship between complex human phenotypes^22,32^. Importantly, they may conceal genetic overlap involving a mixture of concordant and discordant effect directions^33,34^, and they do not account for differences in polygenicity^33^, which governs the extent to which phenotypes may share genetic variants. Moreover, recent analyses using LAVA^34^ and MiXeR^33^ have demonstrated extensive genetic overlap across complex human phenotypes irrespective of the genetic correlations, along with differences in their polygenic architetures^22,27,33–35^. Additionally, genetic analyses have identified overlapping common variants, rare variants and expression profiles between psychiatric and neurological disorders^8,31,32,36–41^, indicative of a partially shared pathobiological basis.

In the present study, we aimed to provide a novel genetic perspective on the clinical distinction between neurological and psychiatric disorders. To this end, we conducted a comprehensive cross-disorder analysis of recent large-scale GWAS datasets to characterize their shared genetic architecture. We applied novel statistical tools that capture distinct forms of genetic overlap and extensive follow up analyses to link the genomic findings to biological pathways and relevant tissue and cell types.

## Results

### Study design (Fig.1)

We curated a collection of well-powered GWAS summary statistics, resulting in data on ten psychiatric disorders (attention-deficit/hyperactivity disorder (ADHD)^42^, anorexia nervosa (AN)^43^, autism spectrum disorder (ASD)^44^, anxiety disorders (ANX)^45^, BD^46^, major depressive disorder (MDD)^47^, obsessive-compulsive disorder (OCD)^48^, post-traumatic stress disorder (PTSD)^49^, SCZ^50^ and Tourette Syndrome (TS)^51^), and ten neurological diseases (ALZ^52^, amyotrophic lateral sclerosis (ALS)^53^, essential tremor (ET)^54^, Lewy body dementia (LBD)^55^, migraine (MIG)^56^, multiple sclerosis (MS)^57^, PD^58–60^, stroke^61^ and the epilepsy subtypes focal epilepsy (FE)^62^ and genetic generalized epilepsy (GGE)^62^). Additionally, we included GWAS data on brain-related traits (general cognitive ability (COG)^63^ and cortical surface area (CRT-SA) and thickness (CRT-TH)^64^), four somatic diseases (chronic kidney disease (CKD)^65^, coronary artery disease (CAD)^66^, inflammatory bowel disease (IBD)^67^ and Type 2 Diabetes (T2D)^68^) and height^69^ as comparators. All GWAS data were limited to participants of European ancestry to avoid bias due to differences in LD structure across ancestries. Ascertainment and diagnostic criteria are described in the Supplementary Note.

**Fig. 1.**
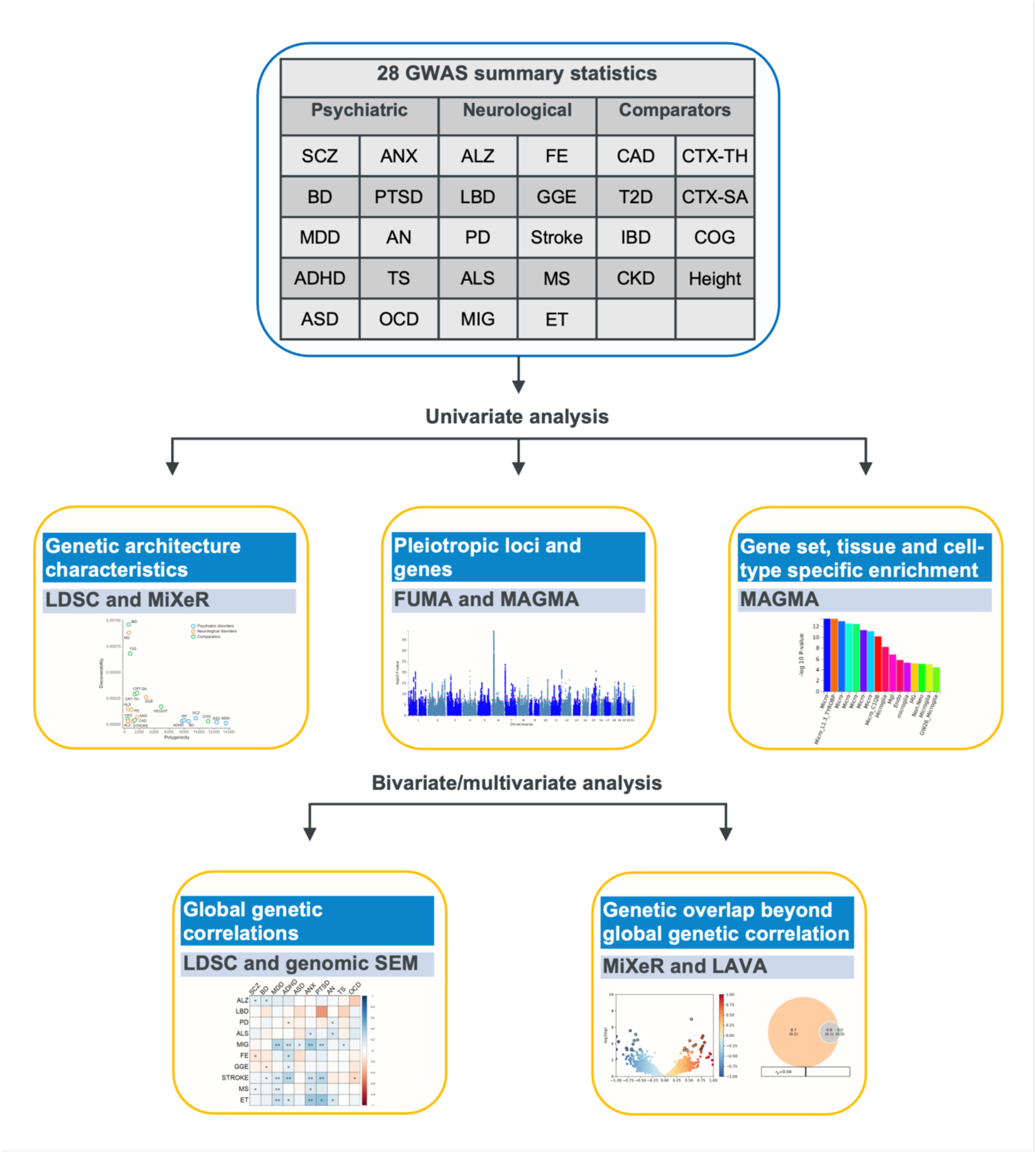
Study design. Overview of the GWAS summary statistics and analyses performed in the study. Abbreviations psychiatric disorders: Attention-deficit/hyperactivity disorder (ADHD), anorexia nervosa (AN), autism spectrum disorder (ASD), anxiety disorders (ANX), bipolar disorder (BD), major depressive disorder (MDD), obsessive-compulsive disorder (OCD), post-traumatic stress disorder (PTSD), schizophrenia (SCZ), Tourette syndrome (TS); neurological diseases: Alzheimer’s disease (ALZ), amyotrophic lateral sclerosis (ALS), Essential Tremor (ET), Lewy body dementia (LBD), migraine (MIG), multiple sclerosis (MS), Parkinson disease (PD), focal epilepsy (FE), genetic generalized epilepsy (GGE); comparators: general cognitive ability (COG), total cortical surface area (CRT-SA) and average cortical thickness (CRT-TH), coronary artery disease (CAD), chronic kidney disease (CKD), inflammatory bowel disease (IBD) and Type 2 Diabetes (T2D); methods: linkage disequilibrium score regression (LDSC), genomic structural equation modeling (genomic SEM).

After data harmonization and pre-processing of the GWAS summary data, we conducted systematic cross-trait analyses and biological interrogation (Fig. 1). We first provide information on the characteristics of the genetic architecture distinguishing each phenotype. Next, we provide an overview of the overlapping genome-wide significant loci and implicated genes. Third, we present the patterns of global genetic correlations across the phenotypes, and clusters of inter-related brain disorders. Fourth, we provide estimates of genetic overlap beyond genetic correlation. Finally, we interrogate the implicated biological pathways, tissues and cell types across the included GWAS.

### Individual genetic architecture characteristics

The genetic architecture of complex human phenotypes differs in terms of the heritability accounted for by single-nucleotide polymorphisms (SNP-heritability), the estimated number of SNPs influencing the phenotype (the polygenicity), and the variance of effect sizes across the associated SNPs (the discoverability)^22,32^. For each phenotype (trait or disorder), we estimated the SNP-heritability using LDSC^70^ (Table 1, Fig. 2a). On average, the estimated SNP-heritability on the liability scale was almost twice as large for psychiatric disorders (14.6%, range 5.3-29.3%) compared to neurological diseases (8.2%, range 1.4-23.8%). Regardless of disease category, however, illnesses with typical onset during childhood or adolescence had the highest estimated SNP-heritability, specifically OCD, GGE, SCZ and TS, all of putative neurodevelopmental origin. The average estimated SNP-heritability for non-brain related diseases was 9.8% (range 1.6-17.9%).

**Fig. 2.**
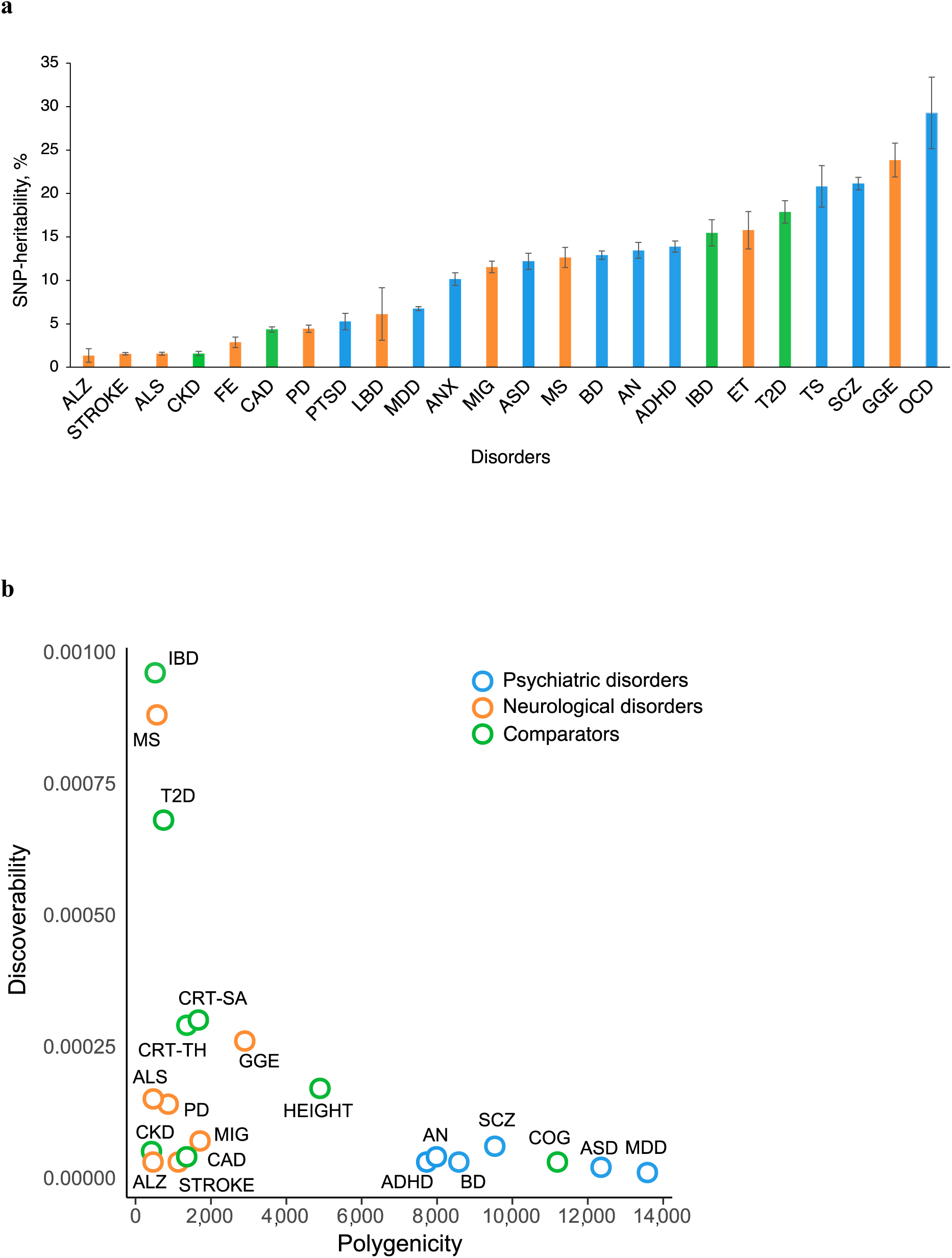
Individual genetic architecture characteristics. **a,** SNP-based heritability on the liability scale for all disorders estimated using LD score regression^70^. **b,** Polygenicity and discoverability of all phenotypes estimated using MiXeR^71^, excluding GWAS with poor model fit. For full univariate MiXeR results, see Supplementary Table 1.

**Table 1.**
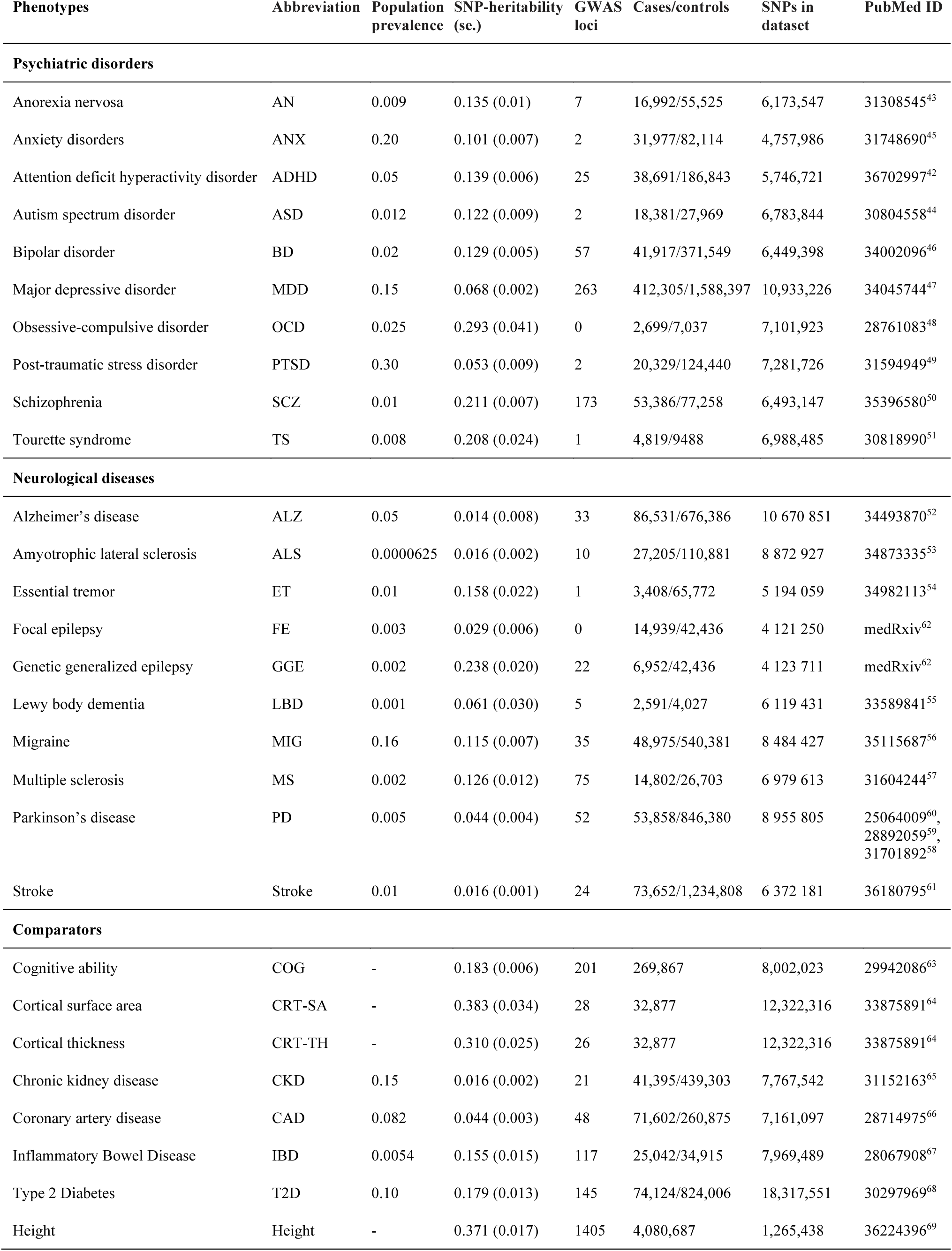
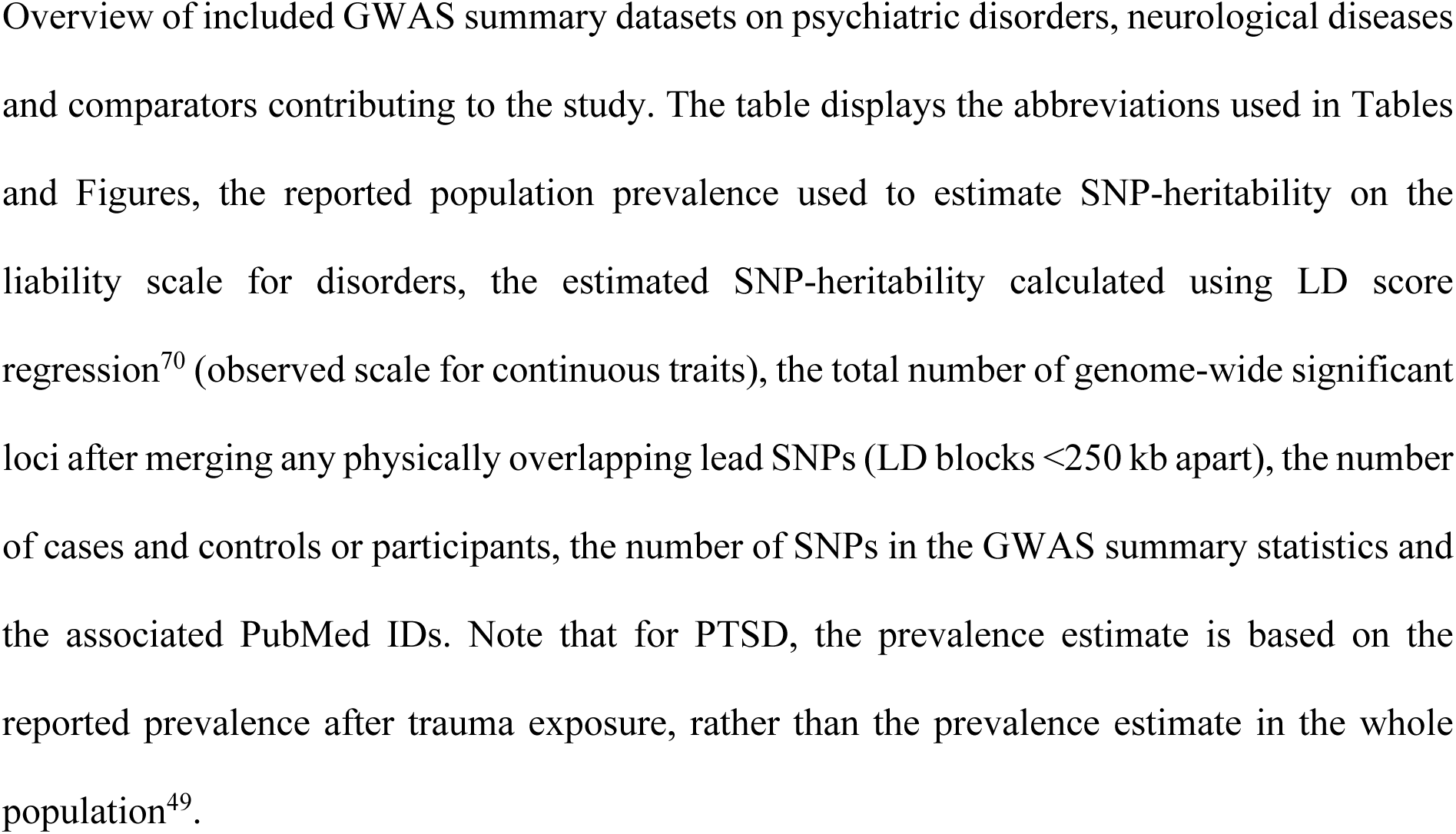
Overview of the GWAS contributing to the study.

| Phenotypes | Abbreviation | Population prevalence | SNP-heritability (se.) | GWAS loci | Cases/controls | SNPs in dataset | PubMed ID |
| --- | --- | --- | --- | --- | --- | --- | --- |
| **Psychiatric disorders** |  |  |  |  |  |  |  |
| Anorexia nervosa | AN | 0.009 | 0.135 (0.01) | 7 | 16,992/55,525 | 6,173,547 | 31308545<sup>43</sup> |
| Anxiety disorders | ANX | 0.20 | 0.101 (0.007) | 2 | 31,977/82,114 | 4,757,986 | 31748690<sup>45</sup> |
| Attention deficit hyperactivity disorder | ADHD | 0.05 | 0.139 (0.006) | 25 | 38,691/186,843 | 5,746,721 | 36702997<sup>42</sup> |
| Autism spectrum disorder | ASD | 0.012 | 0.122 (0.009) | 2 | 18,381/27,969 | 6,783,844 | 30804558<sup>44</sup> |
| Bipolar disorder | BD | 0.02 | 0.129 (0.005) | 57 | 41,917/371,549 | 6,449,398 | 34002096<sup>46</sup> |
| Major depressive disorder | MDD | 0.15 | 0.068 (0.002) | 263 | 412,305/1,588,397 | 10,933,226 | 34045744<sup>47</sup> |
| Obsessive-compulsive disorder | OCD | 0.025 | 0.293 (0.041) | 0 | 2,699/7,037 | 7,101,923 | 28761083<sup>48</sup> |
| Post-traumatic stress disorder | PTSD | 0.30 | 0.053 (0.009) | 2 | 20,329/124,440 | 7,281,726 | 31594949<sup>49</sup> |
| Schizophrenia | SCZ | 0.01 | 0.211 (0.007) | 173 | 53,386/77,258 | 6,493,147 | 35396580<sup>50</sup> |
| Tourette syndrome | TS | 0.008 | 0.208 (0.024) | 1 | 4,819/9488 | 6,988,485 | 30818990<sup>51</sup> |
| **Neurological diseases** |  |  |  |  |  |  |  |
| Alzheimer's disease | ALZ | 0.05 | 0.014 (0.008) | 33 | 86,531/676,386 | 10 670 851 | 34493870<sup>52</sup> |
| Amyotrophic lateral sclerosis | ALS | 0.0000625 | 0.016 (0.002) | 10 | 27,205/110,881 | 8 872 927 | 34873335<sup>53</sup> |
| Essential tremor | ET | 0.01 | 0.158 (0.022) | 1 | 3,408/65,772 | 5 194 059 | 34982113<sup>54</sup> |
| Focal epilepsy | FE | 0.003 | 0.029 (0.006) | 0 | 14,939/42,436 | 4 121 250 | medRxiv<sup>62</sup> |
| Genetic generalized epilepsy | GGE | 0.002 | 0.238 (0.020) | 22 | 6,952/42,436 | 4 123 711 | medRxiv<sup>62</sup> |
| Lewy body dementia | LBD | 0.001 | 0.061 (0.030) | 5 | 2,591/4,027 | 6 119 431 | 33589841<sup>55</sup> |
| Migraine | MIG | 0.16 | 0.115 (0.007) | 35 | 48,975/540,381 | 8 484 427 | 35115687<sup>56</sup> |
| Multiple sclerosis | MS | 0.002 | 0.126 (0.012) | 75 | 14,802/26,703 | 6 979 613 | 31604244<sup>57</sup> |
| Parkinson's disease | PD | 0.005 | 0.044 (0.004) | 52 | 53,858/846,380 | 8 955 805 | 25064009<sup>60</sup>,<br>28892059<sup>59</sup>,<br>31701892<sup>58</sup> |
| Stroke | Stroke | 0.01 | 0.016 (0.001) | 24 | 73,652/1,234,808 | 6 372 181 | 36180795<sup>61</sup> |
| **Comparators** |  |  |  |  |  |  |  |
| Cognitive ability | COG | - | 0.183 (0.006) | 201 | 269,867 | 8,002,023 | 29942086<sup>63</sup> |
| Cortical surface area | CRT-SA | - | 0.383 (0.034) | 28 | 32,877 | 12,322,316 | 33875891<sup>64</sup> |
| Cortical thickness | CRT-TH | - | 0.310 (0.025) | 26 | 32,877 | 12,322,316 | 33875891<sup>64</sup> |
| Chronic kidney disease | CKD | 0.15 | 0.016 (0.002) | 21 | 41,395/439,303 | 7,767,542 | 31152163<sup>65</sup> |
| Coronary artery disease | CAD | 0.082 | 0.044 (0.003) | 48 | 71,602/260,875 | 7,161,097 | 28714975<sup>66</sup> |
| Inflammatory Bowel Disease | IBD | 0.0054 | 0.155 (0.015) | 117 | 25,042/34,915 | 7,969,489 | 28067908<sup>67</sup> |
| Type 2 Diabetes | T2D | 0.10 | 0.179 (0.013) | 145 | 74,124/824,006 | 18,317,551 | 30297969<sup>68</sup> |
| Height | Height | - | 0.371 (0.017) | 1405 | 4,080,687 | 1,265,438 | 36224396<sup>69</sup> |

Overview of included GWAS summary datasets on psychiatric disorders, neurological diseases and comparators contributing to the study. The table displays the abbreviations used in Tables and Figures, the reported population prevalence used to estimate SNP-heritability on the liability scale for disorders, the estimated SNP-heritability calculated using LD score regression<sup>70</sup> (observed scale for continuous traits), the total number of genome-wide significant loci after merging any physically overlapping lead SNPs (LD blocks <250 kb apart), the number of cases and controls or participants, the number of SNPs in the GWAS summary statistics and the associated PubMed IDs. Note that for PTSD, the prevalence estimate is based on the reported prevalence after trauma exposure, rather than the prevalence estimate in the whole population<sup>49</sup>.

Using MiXeR^71^, we estimated the polygenicity and discoverability for each phenotype (Fig. 2b; Supplementary Table 1), except for seven GWAS displaying poor model fit due to insufficient statistical power (ANX, PTSD, TS, OCD, FE, ET and LBD). The polygenicity estimates for all psychiatric disorders (7,725 (SD=349) – 13,582 (SD=387)) and COG (11,195 (SD=369)) exceeded those of neurological diseases (464 (SD=43) – 2,898 (SD=220)), somatic disorders (423 (SD=55) – 1,358 (SD=85)), height (4,894 (SD=90)) and cortical imaging measures (1,361 (SD=100) – 1,666 (SD=125)). For example, the least polygenic psychiatric disorder ADHD (7,725 (SD=349)) was estimated to be influenced by ∼2.7 times more genetic variants than the most polygenic neurological disease GGE (2,898 (SD=220)). In line with prior work^22,71^, the most polygenic phenotypes were characterized by relatively low discoverability, indicating a larger fraction of trait-influencing variants with smaller effect sizes. In Supplementary Fig. 1, we present GWAS power plots displaying the estimated fraction of SNP-heritability explained by genome-wide significant SNPs as a function of sample size, demonstrating that the discovery trajectories for most of the GWAS are still in the early stages, except for height.

### Overlapping genome-wide significant loci and genes

We estimated the number and fraction of significantly associated loci and genes shared across the phenotype categories (Table 2) with results for each phenotypic pair provided in Supplementary Tables 2-3. For each GWAS, we identified genome-wide significant loci according to the FUMA protocol^72^. We subsequently grouped physically overlapping loci, resulting in a total number of 1,988 distinct loci. Of these, 441 loci were linked to psychiatric disorders and 227 loci to neurological diseases. In total, 41 loci were overlapping between psychiatric and neurological disorders, constituting 9.3% and 18.1% of the total number of loci linked to these categories, respectively. Additionally, we mapped GWAS associations to protein-coding genes using MAGMA^73^, yielding a total number of 7,829 distinct genes. Of these, 796 genes were linked to psychiatric disorders and 497 to neurological diseases. A total of 51 genes were shared between psychiatric and neurological disorders, constituting 6.4% and 10.3% of the total number of genes linked to these categories, respectively. As expected, the pleiotropy across genome-wide significant loci and genes were largely driven by GWAS power, warranting cautious interpretation of these results. Most of the pleiotropy for psychiatric disorders were observed for SCZ and MDD, while the neurological GWAS were more evenly powered.

**Table 2.** Overview of pleiotropic loci and genes linked to psychiatric or neurological diseases at the genome-wide significant level.

|  | Loci |  | Genes |  |
| --- | --- | --- | --- | --- |
|  | Count | Fraction (%) | Count | Fraction (%) |
| **Psychiatric disorders** |  |  |  |  |
| **Total** | 441 |  | 796 |  |
| **Pleiotropic with** |  |  |  |  |
| Psychiatric disorders | 60 | 13.6% | 148 | 18.6% |
| Neurological diseases | 41 | 9.3% | 51 | 6.4% |
| Cognitive ability | 64 | 14.5% | 136 | 17.1% |
| Cortical thickness and surface area | 16 | 3.6% | 23 | 2.9% |
| Somatic disorders | 46 | 10.4% | 72 | 9.0% |
| Height | 162 | 36.7% | 463 | 58.2% |
| **Neurological diseases** |  |  |  |  |
| **Total** | 227 |  | 497 |  |
| **Pleiotropic with** |  |  |  |  |
| Psychiatric disorders | 41 | 18.1% | 51 | 10.3% |
| Neurological diseases | 16 | 7.0% | 16 | 3.2% |
| Cognitive ability | 19 | 8.4% | 35 | 7.0% |
| Cortical thickness and surface area | 10 | 4.4% | 19 | 3.8% |
| Somatic disorders | 53 | 23.3% | 58 | 11.7% |
| Height | 109 | 48.0% | 256 | 51.5% |

Count and fraction of pleiotropic genome-wide significant loci and genes linked to psychiatric and neurological disorders across the phenotype categories. After identifying genome-wide significant loci, physically overlapping loci were merged into grouped loci. Protein-coding genes were identified using MAGMA<sup>73</sup>. Across all phenotypes, 1,988 grouped loci and 7,829 genes were identified. The extended MHC-region (chr6: 25–37 Mb) was excluded from these analyses. Supplementary Tables 2-3 present the number of overlapping loci and genes for each pair of phenotypes.

**Table 3.** Summary of tissue and cell type specificity analyses.

|  | Tissue specific associations | Human brain cell type associations |
| --- | --- | --- |
| **Neurological diseases** |  |  |
| ALZ | Upregulated in the spleen, terminal ileum of the small intestine and whole blood | Microglia |
| GGE | - | GABAergic and excitatory neurons |
| MS | Upregulated in EBV-transformed lymphocytes, the spleen, whole blood and the terminal ileum of the small intestine | Microglia and endothelial cells |
| PD | Upregulated in the substantia nigra | - |
| **Psychiatric disorders** |  |  |
| ADHD | Upregulated in the cerebral cortex | GABAergic and excitatory neurons |
| AN | Downregulated in the putamen and hippocampus | - |
| BD | - | GABAergic and excitatory neurons |
| MDD | Upregulated in multiple brain tissues | GABAergic neurons and oligodendrocyte progenitor cells |
| SCZ | Upregulated in multiple brain tissues | GABAergic and excitatory neurons |
| **Comparators** |  |  |
| COG | Upregulated in multiple brain tissues<br>Downregulated in the renal cortex | GABAergic and excitatory neurons |
| CAD | - | Vascular cells |
| CRT-SA | - | Oligodendrocytes, astrocytes, stem cells, and microglia |
| CRT-TH | Upregulated in the cerebellar hemisphere | - |
| IBD | Upregulated in the spleen and whole blood<br>Downregulated in multiple central nervous tissues | Microglia and endothelial cells |
| Height | Up- and downregulated in multiple tissues body-wide | Vascular cells, endothelial cells, astrocytes, oligodendrocytes, and microglia |
| T2D | Downregulated in multiple central nervous tissues | Vascular cells, endothelial cells, astrocytes, oligodendrocytes, and microglia |

### Global genetic correlations

Using bivariate LDSC^24^, we estimated the global pairwise genetic correlations across all phenotypes (Supplementary Fig. 2; Supplementary Table 4). Our results corroborate prior findings of highly intercorrelated genetic risk among psychiatric disorders^25–28,31^. In total, 40 out of 45 genetic correlations among psychiatric disorders reached significance (FDR < 0.05). In comparison, 12 out of 45 correlations among neurological diseases reached significance (FDR < 0.05). As recently demonstrated^29,30^, the neurodegenerative disorders ALS, LBD, ALZ and PD formed a cluster of correlated disorders. Additionally, ET was correlated with both PD (*r*_g_=0.31, p=1.80×10^-7^) and MIG (*r*_g_=0.17, p=3.90×10^-3^), FE was correlated with stroke (*r*_g_=0.30, p=1.40×10^-3^), ALS (*r*_g_=0.32, p=7.10×10^-3^) and the other epilepsy subtype GGE (*r*_g_=0.61, p=8.04×10^-17^), while PD was negatively correlated with both MIG (*r*_g_=-0.08, p=1.40×10^-2^) and stroke (*r*_g_=-0.10, p=1.57×10^-2^).

In total, 30 out of 100 genetic correlations between neurological and psychiatric disorders reached significance at FDR < 0.05 (*r*_g_ range: −0.19 – 0.40; Fig. 3), adding further evidence that genetic risk transcends the categorical boundary between these disorders. We found that MIG, ET and stroke were positively correlated with several psychiatric disorders, in particular MDD, ADHD, ANX and PTSD. The same psychiatric disorders were also correlated with CAD, consistent with a connection between mental disorders and cardiovascular illness^74^. However, neither MIG or ET were significantly correlated with any somatic comparator or stroke, suggesting that their shared genetic effects with psychiatric disorders relate to other aspects. Furthermore, MS was significantly correlated with ANX (*r*_g_=0.17, p=6.00×10^-4^), MDD (*r*_g_=0.11, p=1.16×10^-5^) and SCZ (*r*_g_=0.07, p=1.02×10^-2^). All of these disorders were positively correlated with the immune-mediated disease IBD, indicating a common link to immunity. We also observed significant correlations between ALZ and both BD (*r*_g_=0.14, p=1.81×10^-2^) and SCZ (*r*_g_=0.11, p=1.14×10^-2^), in line with the comorbidity between dementia and psychosis^12,18,31^. Finally, we observed significant correlations between several comparators and psychiatric and neurological disorders, indicating body-wide effects of the involved genetic variants (Supplementary Note).

**Fig. 3.**
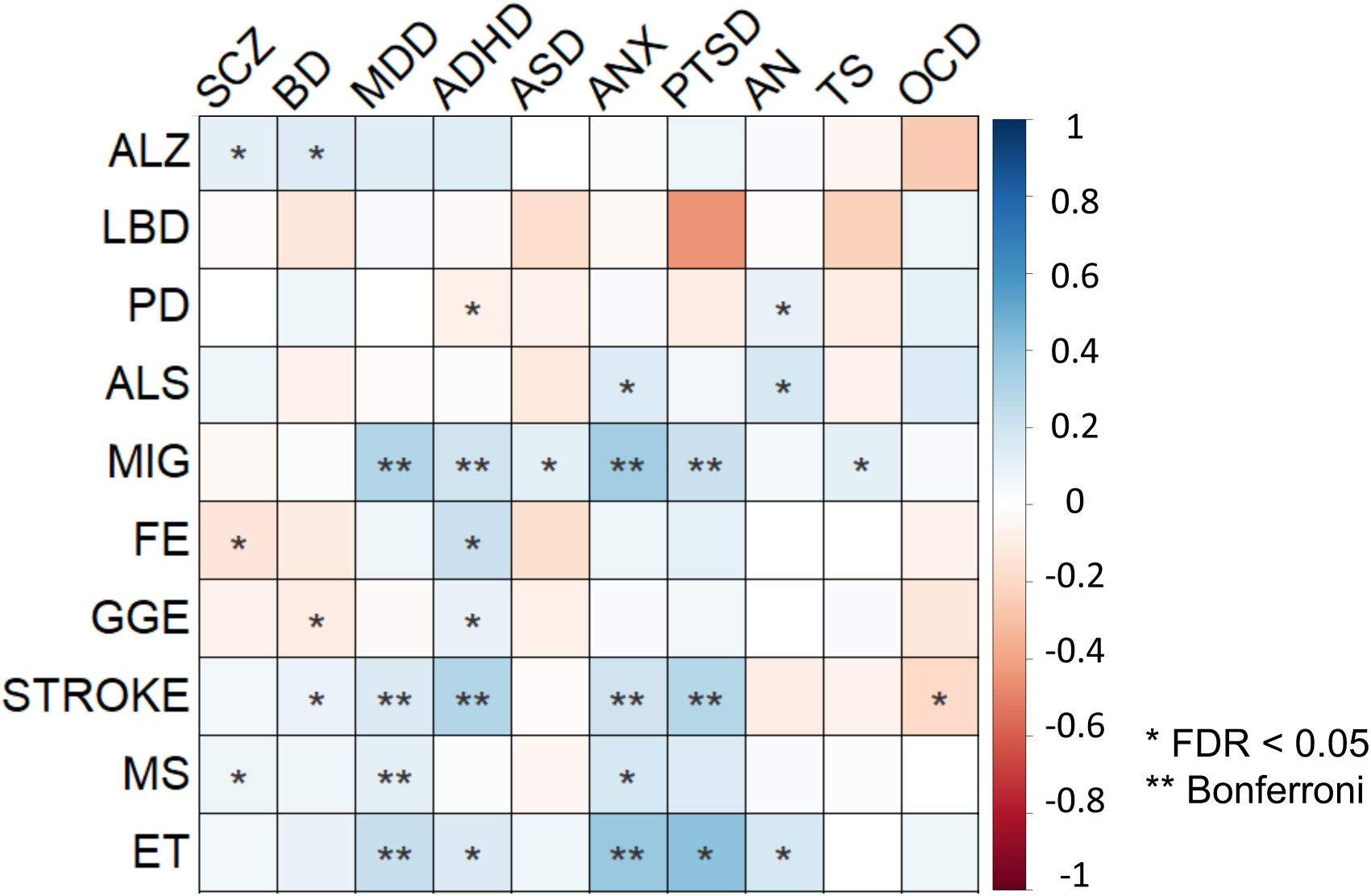
Genetic correlations. Global pairwise genetic correlations across neurological and psychiatric disorders estimated using linkage disequilibrium score regression^24^. One asterisk denotes statistical significance at FDR < 0.05, two asterisks denote statistical significance after Bonferroni correction. The color denotes the magnitude and direction of correlation.

### Genetic covariance structure

Applying genomic SEM^75^, we modeled the genetic covariance structure of neurological and psychiatric disorders. The model did not successfully converge for neurological diseases, likely due to insufficient correlation structure. We then leveraged a recently established factor model for psychiatric disorders^28^, which specified four latent factors that fit the data well (Supplementary Table 5). The first factor consisted of compulsive disorders (AN, OCD and TS), the second factor of psychotic disorders (SCZ and BD), the third factor was characterized by neurodevelopmental disorders (ASD and ADHD) as well as PTSD, MDD and TS, while the fourth factor consisted of internalizing disorders (MDD, ANX and PTSD). We then conducted confirmatory factor analyses (CFAs) and estimated whether any of the neurological diseases correlated with the psychiatric factors. After Bonferroni-correction (0.05/40 = 1.25×10^-3^), four neurological diseases (MIG, stroke, MS and ET) were found to significantly correlate with a psychiatric factor, indicating shared genetic covariance structure with psychiatric disorders. Specifically, MIG was positively correlated with the neurodevelopmental (*r*_g_=0.23, p=7.24×10^-11^) and internalizing (*r*_g_=0.31, p=2.98×10^-31^) factors, stroke was negatively correlated with the compulsive factor (*r*_g_=-0.20, p=6.95×10^-4^), but positively correlated with the neurodevelopmental (*r*_g_=0.28, p=5.66×10^-11^) and internalizing (*r*_g_=0.16, p=2.43×10^-5^) factors, while both MS (*r*_g_=0.15, p=2.12×10^-6^) and ET (*r*_g_=0.30, p=1.62×10^-7^) were positively correlated with the internalizing factor. No neurologic disease was significantly correlated with the psychotic factor.

### Genetic overlap beyond global genetic correlations

Using bivariate MiXeR^33^, we estimated the unique and overlapping genetic architectures between pairs of phenotypes (Supplementary Table 6). Unlike LDSC^24^, MiXeR can detect genetic overlap regardless of the global genetic correlations^33^. Corroborating recent work^35^, we found extensive genetic overlap across all psychiatric disorders, with a minor proportion of disorder-specific variants (Supplementary Fig. 3). MiXeR indicated varying degrees of genetic overlap between neurological diseases, with smaller proportions of shared risk compared to that observed among psychiatric disorders, suggesting that neurological diseases are more genetically distinct from each other. Despite disparate polygenicity estimates, we observed widespread genetic overlap between neurological and psychiatric disorders. This constituted a larger proportion of the genetic architectures of neurological diseases given their smaller polygenicity estimates relative to psychiatric disorders. As an example, MiXeR estimated pronounced genetic overlap between SCZ and neurological diseases PD, GGE and MIG, despite absent genetic correlations, indicative of a balanced mix of concordant and discordant effects among the shared variants (Fig. 4). Almost all genetic variants linked to PD and GGE and 70% of those linked to MIG were estimated to also influence risk of SCZ, while the overlap represented less than 30% of the SCZ variants.

**Fig. 4.**
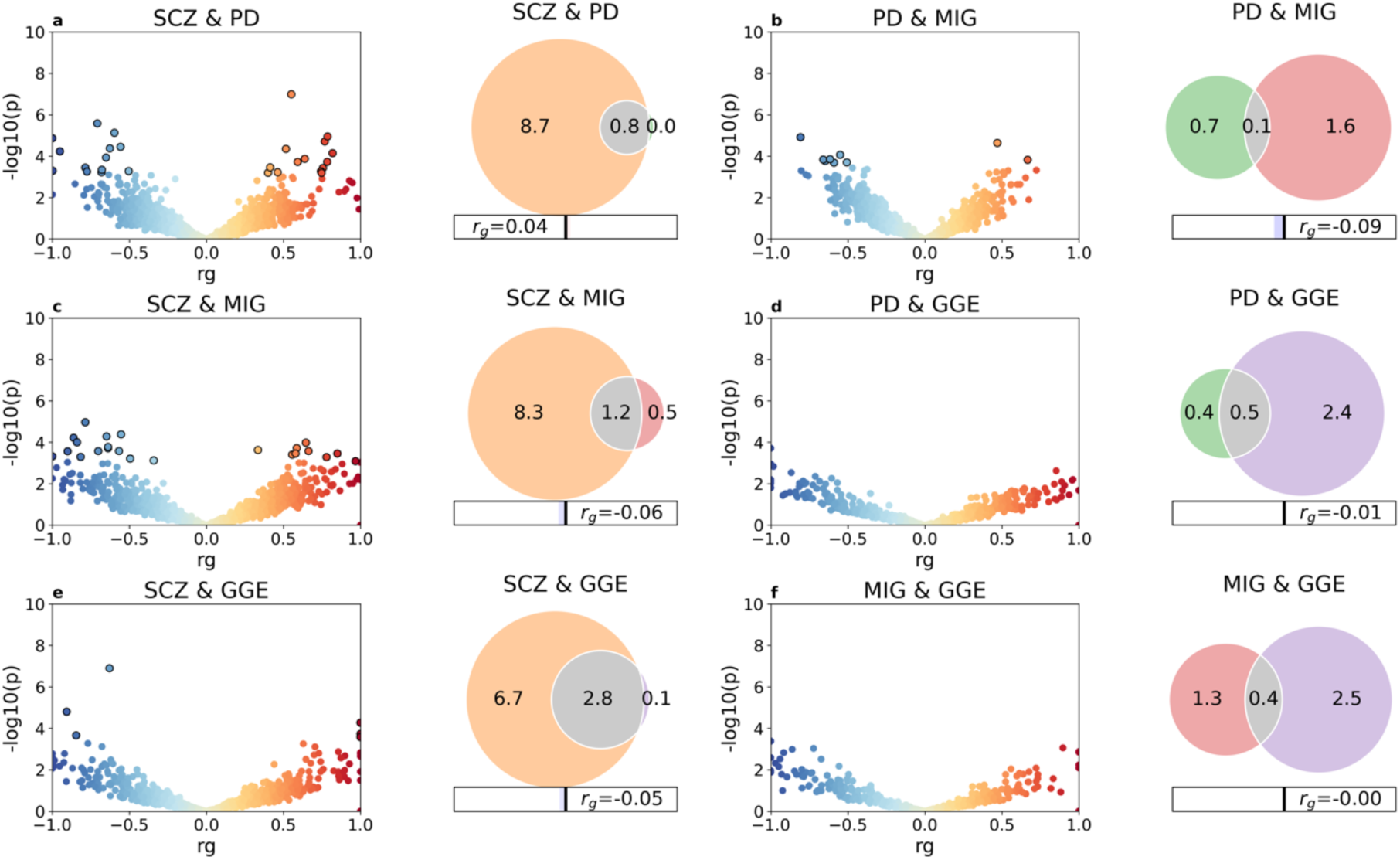
Genetic overlap beyond global genetic correlations. LAVA local correlations and MiXeR-modeled genome-wide genetic overlap for selected disorders schizophrenia (SCZ), Parkinson’s disease (PD), migraine (MIG) and genetic generalized epilepsy (GGE). To the left, volcano plots of local genetic correlation coefficients (rho) against -log_10_ p-values for each pairwise analysis per locus estimated using LAVA^34^ (See Supplementary Table 7 for full results). Dots encircled in black represent significantly correlated loci after false discovery rate correction. To the right, Venn diagrams showing the number (in thousands) of shared and disorder-specific variants and the global genetic correlation (*r*_g_) estimated using MiXeR^33^ (See Supplementary Fig. 3 and Supplementary Table 6 for full results). The total polygenicity for each disorder represents the estimated number of variants required to explain 90% the SNP-based heritability.

Applying LAVA^34^, we calculated the local genetic correlations across 2,495 genomic regions between all pairs of phenotypes. We performed local genetic correlation tests at loci where both phenotypes had heritability estimates significantly different from zero, and corrected for multiple testing using FDR. Corroborating the MiXeR findings, LAVA estimated multiple significantly correlated loci across most pairs of phenotypes, including between neurological and psychiatric disorders (Supplementary Table 7). As observed for locus and gene pleiotropy at the genome-wide significance level (Supplementary Tables 2 and 3), the number of LAVA local correlations largely reflected GWAS power. Consistent with the MiXeR findings, LAVA estimated correlated loci between SCZ and PD (14 positively correlated and 13 negatively correlated loci), GGE (six positively correlated and six negatively correlated loci), and MIG (10 positively correlated and 15 negatively correlated loci), adding further support for a shared genetic basis (Fig. 4).

### Tissue, cell-type and gene-set enrichment analyses

Finally, we compared GWAS enrichment with specific tissues, cell types and gene sets (Table 4), leveraging RNA sequencing data from the Genotype-Tissue Expression (GTEx) project^76^, single-cell RNA sequencing datasets from the developing and adult human brain^77^, and predefined Gene Ontology gene sets implemented in FUMA^72^. We performed Bonferroni correction for the number of tested items in each analysis.

The analyses revealed a range of neural and somatic biological associations associated with neurological diseases. As previously shown^52,57,77^, ALZ and MS were both significantly associated with immune-enriched tissues, microglia and immunological pathways, implying a key role of the immune system. Additionally, ALZ was associated with amyloid-beta related processes. Risk genes for PD were significantly associated with various neurobiological processes, particularly concerning synaptic vesicles, and were specifically upregulated in the substantia nigra^58^, central to PD pathogenesis. Risk genes for GGE were significantly associated with both GABAergic and excitatory neurons^62^, in line with hyperexcitability being the main pathophysiological feature of epilepsy, but were not associated with any tissue or gene set. Stroke was significantly associated with one gene set only, ‘fibrinogen’, an established stroke risk factor involved in clot formation^78^. Risk genes for LBD were linked to lipid metabolism.

Corroborating previous work^26,28,32,42,46,47,50^, GWAS on psychiatric disorders consistently implicated neuronal biology. Risk genes for ADHD, MDD and SCZ were all upregulated in brain tissue, implicated neurobiological processes and neuronal cell types. MDD was also associated with oligodendrocyte progenitor cells. BD risk genes were significantly associated with both GABAergic and excitatory neurons, but not with any tissue or gene set. ANX was significantly associated with several neurobiological pathways, while risk genes for AN were significantly downregulated in specific brain tissues.

Apart from COG, no comparator was significantly associated with neurons. COG and CRT-TH were the only comparators whose genes were significantly upregulated in brain tissue. Further results are described in the Supplementary Note. Full results are provided in Supplementary Tables 8 and 9 and Supplementary Figs. 4 and 5.

## Discussion

In the present study, we elucidate the shared genetic architecture of neurological and psychiatric disorders, indicating that they partly share a genetic basis. Overall, the results represent a major advance in our understanding of the common genetic variation underlying brain-related disorders, suggesting that a large set of genetic variants influence a range of pathogenic processes, wherein disorder specificity is determined by the distribution of effect sizes. While the shared genomic components suggest that neurological and psychiatric disorders partially share molecular genetic mechanisms, a more central role of neuronal biology was implicated in psychiatric disorders, while neurological diseases implicated a larger variety of biological processes. Altogether, the findings are consistent with accumulating evidence indicating that neurological and psychiatric disorders share key etiological aspects, contrasting their clinical distinction.

To compare the genetic basis of neurological and psychiatric disorders, we analyzed GWAS summary data from 20 major disorders, representing the largest cross-disorder analysis on this subject to date (Table 1). Moreover, the application of statistical methods with different modelling assumptions and different techniques for measuring genetic overlap allowed us to interrogate their genetic relationship in a more comprehensive manner than previous work^25^. In the univariate analysis, psychiatric disorders were more polygenic than neurological diseases. Polygenicity indicates the number of additive genetic effects that may combine to yield increased trait susceptibility, providing a measure of genetic architecture complexity and possibly heterogeneity^22,79^. While both neurological and psychiatric disorders are multifactorial and clinically heterogenous, the higher levels of polygenicity of psychiatric disorders is consistent with a hypothesis that multiple causal pathways may converge on the same mental illness, while fewer causal pathways may underlie neurological diseases. Despite similar twin-based heritability estimates across neurological and psychiatric disorders^21^, the SNP-based heritability estimates appeared to negatively correlate with typical onset of illness, regardless of disease category. This contrasts the theoretical expectation that common genetic variants might explain more variance in late-onset disorders, given their weaker impact on reproductive fitness, thereby reducing selective pressure^80^. However, current methodology may inappropriately account for the effect of age and large-effect variants such as *APOE* variants, warranting cautious interpretation of SNP-heritability estimates for late-onset disorders^81^.

Expanding upon previous work based on less powerful GWAS^25,31,36–40^, we demonstrate widespread genetic correlations between neurological and psychiatric disorders, most of which were positive (Fig. 3). The results indicate that neurological and psychiatric disorders partly exist on genetic continua, providing new insights into their genetic relationship. Importantly, the shared genetic components may map onto overlapping biological aspects that could be targeted therapeutically. The pattern of correlations was not uniform across disorders, with clusters of disorders being more correlated with each other. Notably, both MIG and ET were positively correlated with several psychiatric disorders, in particular the internalizing disorders ANX, MDD and PTSD, consistent with their extensive psychiatric comorbidities^16,17^. On a cautious note, however, the GWAS on both MIG and ET were largely based on self-reports^54,56^. Although self-reported and clinically ascertained cases are shown to strongly correlate^47,56,58^, we cannot exclude the possibility that some self-reports were based on underlying mental illness with somatoform symptomatology. The findings nevertheless emphasize the interconnected nature of these disorders, and may motivate further trialing of psychotherapy or antidepressants, which show beneficial effects for MIG prevention^82^.

Beyond genetic correlations, we observed a more pervasive degree of genetic overlap across neurological and psychiatric disorders, involving a mixture of concordant and discordant effect sizes (Supplementary Fig. 3). As an example, MiXeR^33^ indicated that a pronounced fraction of the genetic risk underlying PD, GGE and MIG overlaps with SCZ, despite absence of global genetic correlations (Fig. 4). The findings align with the discovery of multiple correlated genomic regions between these disorders using LAVA^34^ (Fig. 4), and shared loci detected below the genome-wide significance level^36,37,40^. The emerging results indicate a substantial genetic basis shared across neurological and psychiatric disorders, in which multiple common genetic variants impact risk of several disorders, but with divergent effect sizes. Accordingly, a given genetic variant may influence numerous biological pathways, each of which may be differently involved in the pathogenesis underlying distinct brain disorders. This is consistent with recent findings of highly distributed genetic effects across brain morphological, cognitive and personality traits^83–86^, indicating that multiple genetic variants with small effects affect the fine-tuning and dynamic interplay across a range of neural and behavioral systems. From a clinical perspective, the findings are highly relevant to the potential implementation of genomic precision medicine in psychiatry and neurology. Integrating genomic data across multiple disorders in a multivariate prediction framework may aid in identifying individuals who are more likely to experience comorbid symptoms, either endogenously or due to adverse treatment effects, which is currently an unmet clinical need.

The study has some limitations. The analysis was restricted to individuals of European ancestry, given the lack of well-powered GWAS on other ancestries. Trans-ancestral follow-up studies are required to assess the generalizability of these results. The present analysis was based on common genetic variants, but rare variants likely impact the comorbidity between neurological and psychiatric disorders as well. For example, rare variants are jointly associated with epilepsy, SCZ and ASD^32^. Our study was limited by bias inherent to the original GWAS, including population stratification and ascertainment procedures. As noted above, misdiagnosis could affect the results, in particular with more common disorders like anxiety or depression. However, prior extensive simulations did not find that misdiagnosis could explain the magnitude of correlated risk across psychiatric disorders^25,26^. Comorbid illness may also bias the assessment of genetic overlap, warranting more deeply phenotyped cohorts to assess differential genetic overlap among clinical subtypes. The results may be affected by LD, whereby a causal variant may be correlated with multiple nearby variants, leading to spurious pleiotropy. To address this, statistical fine-mapping follow-up studies are needed. Finally, there was uneven power among the included GWAS, which limit the value of cross-disorder comparison at the present stage. This particularly affects the biological interpretation of the mapped genes, which only represent a minor fraction of the genetic risk architectures underlying these disorders. As GWAS samples get larger, cross-trait analyses based on more diverse datasets, additional disorders, and specific subtypes, should be conducted.

In conclusion, by leveraging recent large-scale GWAS datasets and novel statistical tools, we demonstrate that neurological and psychiatric disorders partly share genetic etiology and biological associations related to the brain. Incorporating these complex and interconnected illnesses into a more unified framework may help accelerate progress in these fields, lead to a more coherent and productive clinical approach^3–8^, and promote precision medicine implementation.

## Acknowledgments

We thank the research participants, employees and researchers of the UK Biobank, 23andMe, Inc., MVP, iPSYCH and the many consortia for making this research possible. This research has been conducted using the UK Biobank Resource under Application Number 27412. Moreover, the research has been conducted using and data from MVP, under dbGap accession number phs001672. This work was partly performed on the TSD (Services for Sensitive Data) facilities, owned by the University of Oslo, operated and developed by the TSD service group at the University of Oslo, IT-Department (USIT). Computations were also performed on resources provided by UNINETT Sigma2—the National Infrastructure for High Performance Computing and Data Storage in Norway. We gratefully acknowledge support from the American National Institutes of Health (NS057198, EB000790, 1R01MH124839, R01MH120219, RF1AG073593), the Research Council of Norway (RCN) (229129, 213837, 324252, 300309, 273291, 223273, 248980, 326813), the South-East Norway Regional Health Authority (2019-108, 2022-073), KG Jebsen Stiftelsen (SKGJ-MED-021), EAA grant (#EEA-RO-NO-2018-0573). This project has received funding from the European Union’s Horizon 2020 research and innovation programme under grant agreement No 847776 and 964874 and 801133 (Marie Sklodowska-Curie grant agreement).

## Author contributions

O.B.S. conceived the study. O.B.S., G.K., S.B., V.F., N.P., G.H., O.F., A.S., and O.A.A. designed the study. G.K., N.P., O.F., and A.S. performed quality control on the GWAS summary data. G.K., S.B., D.M., A.S. and O.F. conducted the formal analysis. O.B.S., A.S., A.D.G., O.F., A.M.D. and O.A.A. provided supervision. O.B.S. wrote the paper. O.B.S. and O.A.A. administered the project. A.M.D. and O.A.A. provided resources and acquired funding. All authors provided valuable discussions and critical input to the data. The funders had no role in the conceptualization, design, data collection, analysis, decision to publish, or preparation of manuscript.

## Additional information

Supplementary information is available for this paper. Correspondence should be directed to O.B.S.

## Methods

### GWAS summary statistics

We collated large-scale GWAS summary statistics based on available sample sizes and the quality of the phenotyping procedures (Table 1; See Supplementary Note for description of each GWAS dataset). All individuals included in the analysis were of European ancestry. Informed consent was obtained from all participants in the respective GWAS. The Regional Committee for Medical Research Ethics – Southeast Norway evaluated the current protocol and found that no additional institutional review board approval was necessary as no individual data were used. All GWAS datasets were derived from existing GWAS except the two datasets on total cortical surface area and average cortical thickness, which were generated from the UK Biobank under accession number 27412, after excluding all individuals with neurological and psychiatric disorders (Supplementary Note). For epilepsy, we chose to include its two main subtypes, focal epilepsy and genetic generalized epilepsy (GGE), rather than including the phenotype ‘all epilepsies combined’, due to the substantial differences in the genetic risk architectures underlying these two subtypes^62^, as emphasized by their differences in estimated SNP-heritability (2.9% vs 23.8%, respectively; Table 1). Before commencing analysis, all GWAS summary statistics underwent uniform quality control and were harmonized and preprocessed into a consistent file structure with a common reference for positions, rsIDs and effect alleles using the v1.6.0 cleansumstats pipeline^87^.

### Genome-wide significant loci

For each GWAS, we defined independently associated genomic loci using FUMA^72^. First, we identified independent significant SNPs with a genome-wide significant p-value (5×10^-8^) that were independent from each other at *r*^2^<0.60. LD *r*^2^ values were obtained from the 1000 Genomes Project European-ancestry haplotype reference panel^88^. The borders of the loci were defined by identifying all candidate SNPs in LD (*r*^2^≥0.6) with one of the independent significant SNPs in the locus. All loci less than 250kb apart were merged.

To evaluate locus pleiotropy, we used the procedure previously applied by Watanabe et al. (2019)^22^. After identifying genome-wide significant loci for each phenotype, we grouped any physically overlapping loci across all phenotypes. A grouped locus could therefore contain more than one independent locus for a given phenotype if several loci were combined (i.e., loci *A* and *C* could both overlap with locus *B* but not with each other, but they would be grouped into one locus resulting in a continuous genomic region). Each grouped locus was then assigned to their specific phenotypes and the following categories: psychiatric disorders, neurological diseases, COG, cortical MRI measures (CRT-SA and CRT-TH), somatic diseases and height. We then determined the number and fraction of grouped loci shared across categories and between all pairs of phenotypes. The extended MHC-region (chr6: 25–37 Mb) was excluded from this analysis due to its complex LD structure.

### MAGMA gene, gene-property and gene-set analysis

For each GWAS dataset, we identified significantly associated protein-coding genes and gene-sets using MAGMA (v1.08)^73^ as implemented in FUMA^72^ with default settings, using the SNP-wise mean model and the European 1000 Genomes reference cohort phase 3 as reference panel. The input SNPs were mapped to 20,260 protein-coding genes, excluding the extended MHC-region (chr6: 25–37 Mb). Gene boundaries were expanded to 35 kb upstream and 10 kb downstream to include probable regulatory regions outside the transcribed region^89^. Genes were considered significant if the p-value was less than 0.05 after Bonferroni correction for the number of tested genes (0.05/20,260 = 2.47×10^-6^). MAGMA calculates an association p-value for each gene based on the aggregate of all SNPs mapped to each gene, accounting for gene-size, number of SNPs in a gene and LD between markers. We then carried out competitive gene-set analysis based on the identified genes in each phenotype. Specifically, we focused on the Gene Ontology gene set terms: biological processes (7,350 gene sets), cellular components (1,001 gene sets) and molecular functions (1,645 gene sets) obtained from MsigDB version 7.0^90^. Gene sets were considered significant if the p-value was <0.05 after Bonferroni correction for the number of tested gene sets in each category (0.05/7,350 = 6,80×10^-6^, 0.05/1,001 = 5,00×10^-5^, 0.05/1,645 = 3.04×10^-5^, respectively).

Based on the gene-based results above, we carried out tissue specific expression analysis in 54 adult tissue types based on RNA sequencing data GTEx v.8^76^ implemented in FUMA^72^. Tissues were considered significant if the *P* value was less than 0.05 after Bonferroni correction for 54 tissues. For cell type specificity analysis, we tested for enrichment in 24 single-cell RNA sequencing data sets from the developing and adult human brain available in FUMA using MAGMA gene-property analysis^77^. The specific datasets were: Allen_Human_LGN_level1^91^, Allen_Human_LGN_level2^91^, Allen_Human_MTG_level1^91^, Allen_Human_MTG_level2^91^, DroNc_Human_Hippocampus^92^, GSE104276_Human_Prefrontal_cortex_all_ages^93^, GSE104276_Human_Prefrontal_cortex_per_ages^93^, GSE67835_Human_Cortex^94^, Linnarsson_GSE101601_Human_Temporal_cortex^95^, Linnarsson_GSE76381_Human_Midbrain^96^, PsychENCODE_Developmental^97^ PsychENCODE_Adult^97^, and GSE168408_Human_Prefrontal_Cortex datasets from level 1 to 2, spanning six developmental stages: fetal, neonatal, infancy, childhood, adolescence and adult^98^. In the cell type specific analysis, systematic stepwise conditional analysis was performed within datasets to ensure that complex batch effects did not lead to false positives, as well as Bonferroni correction for multiple testing of 379 cell types (0.05/379 = 1.30×10^-4^).

All statistical tests conducted using MAGMA were one sided. We did not perform additional correction for multiple testing across the 28 phenotypes, since the aim of analysis was not to determine which of the phenotypes a specific gene, gene-set, tissue or cell type was associated with, but to explore group level patterns of shared associations across the phenotypes.

### SNP-heritability and global genetic correlations

Using LDSC^70^, we estimated the SNP-based heritability in the liability scale for each disorder, using reported population prevalence estimates (Table 1), and the SNP-based heritability on the observed scale for the continuous traits. LDSC distinguishes confounding from polygenicity by regressing the association statistics of SNPs on their LD scores^70^. All analyses were based on HapMap 3 SNPs only, with the MHC region (chr6: 25–34 Mb) excluded. Precalculated LD scores from the European 1000 Genomes reference cohort were used (https://data.broadinstitute.org/alkesgroup/LDSCORE/eur_w_ld_chr.tar.bz2). Additionally, we used the bivariate extension of LDSC^24^ to estimate the global genetic correlations, i.e. the covariance in the SNP-heritability, between all pairs of phenotypes. Adjusting for the number of traits tested, we applied both the FDR method of Benjamini-Yekutieli^99^ given the dependence between the tests and Bonferroni-correction.

### Genomic SEM

Genomic SEM^75^ models the multivariate genetic architecture across traits and may uncover broad latent factors underlying genetic correlations, reveal clusters of correlated traits, and determine how latent factors correlate with each other. Genomic SEM analysis typically involves two steps, exploratory factor analysis (EFA) and CFA. While EFA identifies the most appropriate number of latent factors, assigning factors to specific traits given sufficiently high loadings to those factors, the resulting model is validated using CFA. In the present study, we first aimed to conduct a joint genomic SEM model of psychiatric and neurological disorders. However, the model did not successfully converge for neurological diseases, likely due to insufficient correlation structure across the neurological diseases. Instead, we applied a recently established factor model for psychiatric disorders^28^, and conducted separate CFAs for each individual neurological disease and the group of psychiatric disorders and estimated whether any of the neurological diseases significantly correlated with the psychiatric factors. We performed Bonferroni correction for the number of tested items (N=10*4, testing loadings for 10 neurological diseases onto 4 psychiatric factors). The CFA was conducted using all available variants, without partitioning genetic variants into even and odd chromosomes, as the analysis only included the CFA stage using a set of predefined models. The goodness of fit was evaluated through standard metric (AIC, CFI, SRMR, presented in Supplementary Table 5), showing appropriate model fit.

### Univariate and bivariate MiXeR analysis

We first applied univariate MiXeR^71^ analysis to each GWAS summary dataset to estimate the proportion of causally associated genetic variants from a reference panel (the polygenicity) and the variance of effect size per causal variant (the discoverability) using maximum likelihood estimation, and the GWAS sample size necessary to discover genetic variants that explain 90% of SNP-heritability of each phenotype. We applied a threshold of 90% SNP-heritability to avoid extrapolating model parameters into variants with infinitesimally small effects. MiXeR is based on a Gaussian mixture model, assuming that a given GWAS summary dataset can be modeled as a “mixture” of pre-defined components with causal and non-causal variants, each with its own Gaussian (normal) distribution. MiXeR incorporates the effects of LD structure, minor allele frequency, GWAS sample size, genomic inflation due to cryptic relatedness, and sample overlap (in the bivariate extension). Before analysis, the MHC region was excluded from all GWAS, while the chromosome 19 was in addition excluded from ALZ due to the strong effects of the *APOE* region^52^ and complicated LD that biases the estimates of polygenicity.

Informed by the model parameters from univariate MiXeR for each phenotype, MiXeR constructs a bivariate mixture model for pairs of phenotypes, in which a mixture of four bivariate Gaussian components is modeled: variants influencing one phenotype only, variants influencing both phenotypes, and variants that are not associated with either phenotype. Bivariate MiXeR estimates the polygenicity of the shared component irrespective of effect directions and correlation of effect sizes. Additionally, MiXeR estimates the genetic correlation of shared variants, and the global genetic correlation. Model fit is evaluated by calculating the difference between the Akaike information criterion (AIC) for best-fitting MiXeR estimates and reference models. Positive AIC differences are interpreted as evidence that the best-fitting MiXeR estimates are distinguishable from the reference model. For univariate MiXeR, an “infinitesimal model” in which all variants are assumed to be ‘causal’ is used as the reference. For bivariate MiXeR, AIC differences are calculated by comparing the best-fitting model to minimum possible overlap, constrained by *r*_g_, and maximum possible overlap, constrained by the polygenicity of the least polygenic trait. We provide conditional Q-Q plots and log-likelihood plots to visualize the stability of the fitness procedure.

### Estimating local genetic correlations using LAVA

For all pairs of phenotypes, we applied LAVA (v1.3.8) to estimate local genetic correlations across 2,495 semi-independent genetic loci of approximately equal size (∼1 Mb). LAVA accounts for potential sample overlap using LDSC^70^. After computing local SNP-heritability estimates for each phenotype, we conducted pairwise local genetic correlation analysis for all loci with local SNP-heritability significantly different from zero. We applied FDR correction to account for multiple comparisons. The statistical tests conducted were all two sided.

## Code availability

Cleansumstats pipeline (https://github.com/BioPsyk/cleansumstats)

FUMA (https://fuma.ctglab.nl/)

Genomic SEM (https://github.com/MichelNivard/GenomicSEM)

LAVA (https://github.com/josefin-werme/LAVA)

LDSC (https://github.com/bulik/ldsc)

MAGMA (https://ctg.cncr.nl/software/magma)

MiXeR (https://github.com/precimed/mixer)

PLINK (https://www.cog-genomics.org/plink/2.0/)

Regenie (https://rgcgithub.github.io/regenie)

## Data availability

All data are publicly available or available on request.

