## Supplementary Information for "The shared genetic risk architecture of neurological and psychiatric disorders: a genome-wide analysis"

**Affiliations**

### 1 Supplementary Methods

#### 1.1 Descriptions of GWAS summary data

GWAS summary statistics for 28 phenotypes were included in this cross-trait analysis. A summary of the samples contributing to each GWAS, including the phenotype definition, is provided below. Detailed information about cohort design, genotyping, quality control and imputation can be found in the original publications (Table 1).

##### 1.1.1 Neurological diseases

**Alzheimer's disease (ALZ).** We retrieved GWAS summary statistics based on 12 cohorts contributing to the Wightman *et al.* (2021) GWAS on ALZ[1], yielding a total sample of 86,531 cases and 676,386 controls, all of European ancestry. Depending on the cohort, cases had been clinically diagnosed with ALZ according to ICD-9 or ICD-10 criteria, were found to have definite, probable or possible ALZ at a memory clinic, or were identified in medical registries as having been prescribed Donepezil (Aricept). Additionally, a proxy phenotype for ALZ case-control status was generated from a self-report questionnaire from the UK Biobank[2], in which participants reported whether their biological mother or father ever suffered from ALZ/dementia, and they reported each parent's current age (or age at death, if applicable).

**Amyotrophic lateral sclerosis (ALS).** 117 cohorts were meta-analyzed, yielding a total sample of 27,205 cases and 110,881 controls. All cases were diagnosed and ascertained through specialized motor neuron disease clinics where they were diagnosed with ALS according to the (revised) El Escorial Criteria[3] by neurologists specialized in motor neuron diseases. All individuals were of European ancestry.

**Essential tremor (ET).** Two clinical cohorts, comprising of samples from multiple centers, were meta-analyzed together with data from two population-based cohorts from the UK Biobank and 23andMe, yielding a total sample of 7,177 cases and 475,877 controls[4]. In the clinical cohorts, cases were diagnosed according to consensus criteria from the Movement Disorder Society (MDS), the Tremor Research Investigational Group (TRIG), other clinical or research criteria, or review of longitudinal movement disorder clinical notes. In the population-based cohorts, cases self-reported as having ET. All individuals were of European ancestry.

**Focal epilepsy (FE) and genetic generalized epilepsy (GGE).** The multi-ancestry GWAS[5] on the broad epilepsy subtypes FE and GGE by the International league against epilepsy (ILAE) included 14,939 cases with FE, 6,952 cases with GGE and 42,436 controls of European ancestry that were included in the present analysis. All cases were classified by neurologists.

**Lewy body dementia (LBD).** The GWAS on LBD[6] comprised of multiple sites/consortia from Europe and North America, of which GWAS summary data on 2,591 cases and 4,027 controls of European ancestry were included in the present analysis. Cases were diagnosed with pathologically definite or clinically probable disease according to consensus criteria.

**Migraine (MIG).** We retrieved GWAS summary data based on four study collections contributing to the GWAS on MIG by the International Headache Genetics Consortium (IHGC) [7], totaling 48,975 cases and 540,381 controls. Most migraine diagnoses were self-reported, while a subset of cases was phenotyped in headache specialist centers. All individuals were of European ancestry.

**Multiple sclerosis (MS).** We retrieved GWAS summary data on 14,802 cases and 26,703 controls contributing to the GWAS on MS by the International Multiple Sclerosis Genetics Consortium (IMSGC)[8]. Cases were determined according to standard diagnostic criteria. All individuals were of European ancestry.

**Parkinson's disease (PD).** The GWAS summary data was based on a meta-analysis from three different GWAS[9-11], yielding a total sample of 53,858 cases and 846,380 controls. A subset of cases was ascertained by clinicians[10], a subset in the 23andMe cohort self-reported as having PD in a web-based survey[11], while another subset was determined as proxy-cases using UK Biobank data (individuals who do not have Parkinson's disease but have a first degree relative that does)[9]. All individuals were of European ancestry.

**Stroke.** We included GWAS summary statistics on any stroke (comprising ischemic stroke, intracerebral hemorrhage, and stroke of unknown or undetermined type) contributing to a recent GWAS on stroke[12]. Cases were determined according to various clinical criteria depending on the cohort. All individuals were of of European ancestry.

##### 1.1.2 Psychiatric disorders

**Anorexia nervosa (AN).** Thirty-three datasets were meta-analyzed, yielding 16,992 cases and 55,525 controls[13]. Cases met criteria for lifetime diagnosis of AN via hospital or register records, structured clinical interviews, or online questionnaires based on standardized criteria (DSM-III-R, DSM-IV, ICD-8, ICD-9 or ICD-10), while in one contributing cohort, the UK Biobank, cases self-reported diagnosis of AN. All individuals were of European ancestry.

**Anxiety disorders (ANX).** Three cohorts were meta-analyzed[14], yielding a total sample of 31,977 cases and 82,114 controls. In the first cohort from the UK Biobank, cases self-reported a lifetime professional diagnosis for at least one of the core five ANX (generalized anxiety disorder, social phobia, panic disorder, agoraphobia or specific phobia). Additionally, some cases met criteria for a likely lifetime diagnosis of DSM-IV generalized anxiety disorder based on anxiety questions from the Composite International Diagnostic Interview Short-form questionnaire. In the second cohort from the ANGST study[15], cases met DSM criteria for at least one of the five core ANX. In the third cohort from iPSYCH[16], diagnosis of one of the five ANX was made by a psychiatrist. All individuals were of European ancestry.

**Attention deficit hyperactive disorder (ADHD).** Twelve cohorts, including data from iPSYCH, deCODE genetics and ten cohorts collected by the PGC, were meta-analyzed, yielding a sample of 38,691 cases and 186,843 controls[17]. In the iPSYCH cohort, cases were diagnosed by psychiatrists according to ICD-10 criteria identified using the Danish Psychiatric Central Research Register and the Danish National Patient register. In the deCODE cohort, cases had been diagnosed according to ICD-10 criteria, or had been prescribed medication specific for ADHD symptoms. Depending on the PGC cohorts, cases were recruited from clinics, hospitals or through medical registries and diagnosed using research-based assessments administered by clinicians or trained staff. All individuals were of European ancestry.

**Autism spectrum disorder (ASD).** Five family-based cohorts and a population-based case-control sample from iPSYCH were meta-analyzed[18], yielding a sample of 18,381 cases and 27,969 controls. In the family-based cohorts, diagnosis was confirmed for all affected individuals using standard research tools and expert clinical consensus diagnosis. In the population-based sample, cases were identified using the Danish Psychiatric Central Research Register and were diagnosed with ASD before 2014 by a psychiatrist according to ICD-10. All individuals were of European ancestry.

**Bipolar disorder (BD).** Fifty-seven cohorts collected in Europe, North America and Australia were meta-analyzed, yielding a total sample of 41,917 cases and 371,549 controls[19]. For 52 cohorts, cases met international consensus criteria (DSM-IV, ICD-9 or ICD-10) for lifetime diagnosis of BD, established using structured diagnostic interviews, clinician-administered checklists or medical record review. For five external cohorts (iPSYCH, deCODE genetics, Estonian Biobank, Trøndelag Health Study (HUNT) and UK Biobank), cases were ascertained using ICD codes or self-report during a nurse interview. All individuals were of European ancestry.

**Major depressive disorder (MDD).** We meta-analyzed separate GWAS data on MDD, from the PGC[20], 23andMe[21], UK Biobank[22], Million Veteran Program[23], and FinnGen[24], yielding a total sample of 412,305 cases and 1,588,397 controls. In the PGC cohort[20], lifetime diagnosis of MDD was ascertained using structured clinical interviews (DSM-V, ICD-9, ICD-10), clinician-administered checklists, or review of medical records, while some met broader criteria. In the 23andMe cohort[21], cases were defined according to a self-reported clinical diagnosis of depression. In the UK Biobank cohort[22], case and control status of broad depression was defined by the participants' response to the questions 'Have you ever seen a general practitioner for nerves, anxiety, tension or depression?' or 'Have you ever seen a psychiatrist for nerves, anxiety, tension or depression?', while participants who either identified with bipolar disorder, schizophrenia, or personality disorder, or were prescribed antipsychotic medications, were excluded. In both the Million Veteran Program and FinnGen cohorts, case definitions were derived from electronic health record data based on ICD-9 and -ICD-10 codes for MDD. All individuals were of European ancestry.

**Obsessive compulsive disorder (OCD).** Two cohorts were meta-analyzed yielding 2,688 cases and 7,037 controls[25]. All cases met DSM-IV criteria for OCD. All individuals were of European ancestry.

**Post-traumatic stress disorder (PTSD).** We included only individuals of European ancestry of an ancestrally diverse meta-analysis of 60 different PTSD-cohorts[26], yielding 20,329 cases and 124,440 controls. Cases were determined either by lifetime or current PTSD. Diagnosis was established using various instruments and different versions of the DSM (DSM-III-R, DSM-IV, DSM-5).

**Schizophrenia (SCZ).** We excluding all individuals of non-European ancestry from the recent GWAS on SCZ from the PGC[27], yielding a sample of 53,386 cases and 77,258 controls across 75 cohorts. Depending on the cohort, cases were defined as individuals diagnosed with schizophrenia, schizoaffective disorder, or schizophrenia spectrum disorders, determined by research diagnostic interview, expert clinical consensus diagnosis, or review of medical records or hospital registers.

**Tourette syndrome (TS).** The GWAS consisted of three case-control cohorts and one family-based cohort including 4,819 cases and 9,488 controls[28]. All cases met DSM-IV-TR or DSM-5 criteria for TS, while 12 cases met DSM-5 criteria for chronic motor or vocal tic disorder. Cases were recruited through TS specialty clinics or were identified by email or online recruitment combined with validated, web-based phenotypic assessments. All individuals were of European ancestry.

###### 1.1.3 Comparators

**Average cortical thickness (CRT-TH) and total cortical surface area (CRT-SA).** To generate the neuroimaging data, we used the Freesurfer v5.3 automatical cortical pipeline[29] to process T1-weighted MRI data from all white Europeans participating in the UK Biobank, under accession number 27412. The final neuroimaging sample included 32,877 individuals, after excluding individuals with poor scan quality, and all individuals with a neurological, psychiatric or brain-related disorder, specifically the ICD10 diagnoses 'Malignant neoplasms of eye, brain and other parts of central nervous system' (C69-C72), 'Mental, Behavioral and Neurodevelopmental disorders' (F00-F99), 'Diseases of the nervous system' (G00-G99), 'Cerebrovascular diseases' (I60-I69), and 'Congenital malformations of the nervous system' (Q00-07), to avoid bias when estimating genetic overlap with neurological and psychiatric disorders. Age, sex, scanner location, scan quality and the first twenty genetic principal components were included as covariates and a rank-based inverse normal transformation was applied to the phenotype when conducting association analysis. We further made use of the UK Biobank v3 imputed data. The GWAS on brain morphology measures were carried out using the standard additive model of linear association in regenie v3.2.5.[30]

**General cognitive ability (COG).** 14 epidemiological cohorts contributed to the GWAS on COG[31], including a total of 269,867 participants of European ancestry. Various neurocognitive tests, primarily assessing fluid domains of cognitive functioning, were conducted across the cohorts.

**Chronic kidney disease (CKD).** We retrieved GWAS summary data on 41,395 cases and 439,303 controls of European ancestry contributing to a GWAS on CKD[32]. CKD was defined as an eGFR below 60 ml $min^{-1}$ per 1.73 $m^2$.

**Coronary artery disease (CAD).** We included GWAS summary statistics on 71,602 cases and 260,875 controls of European ancestry contributing to GWAS on CAD[33-35]. Cases were defined by an inclusive CAD phenotype, for example fatal or nonfatal myocardial infarction, acute coronary syndrome, percutaneous transluminal coronary angioplasty, coronary artery bypass grafting, chronic ischemic heart disease, angina and coronary stenosis of >50%.

**Inflammatory bowel disease (IBD).** A total of 25,042 cases and 34,915 controls contributed to the GWAS on IBD, all of European ancestry[36]. Diagnosis of IBD was based on accepted radiological, endoscopic and histopathological criteria.

**Type 2 Diabetes (T2D).** The GWAS summary statistics on T2D was based on a meta-analysis of 32 GWAS, including 74,124 cases and 824,006 controls, all of European ancestry[37]. Cases were defined according to standard diagnostic criteria.

**Height.** We included GWAS summary statistics on 4,080,687 individuals of European ancestry contributing to the recent GWAS on height[38].

#### 2 Supplementary Results

##### 2.1 Global genetic correlations

Using linkage disequilibrium score regression (LDSC)[39] (Fig. 3; Supplementary Table 4), we estimated significant genetic correlations between ET and five psychiatric disorders at FDR < 0.05. Specifically, ET was positively correlated with PTSD ($r_g$=0.40, p=4.00x10$^{-4}$), ANX ($r_g$=0.37, p=4.63x10$^{-6}$), MDD ($r_g$=0.22, p=6.08x10$^{-7}$), AN ($r_g$=0.17, p=2.80x10$^{-3}$) and ADHD ($r_g$=0.15, p=7.40x10$^{-3}$), indicating shared genetic effects. MIG, which was positively correlated with ET ($r_g$=0.17, p=3.90 x10$^{-3}$), showed a similar pattern of genetic correlations with psychiatric disorders as ET. Specifically, MIG was significantly correlated with ANX ($r_g$=0.35, p=8.59x10$^{-16}$), MDD ($r_g$=0.29, p=1.71x10$^{-45}$), PTSD ($r_g$=0.22, p=1.00x10$^{-4}$), ADHD ($r_g$=0.20, p=6.83x10$^{-10}$), ASD ($r_g$=0.12, p=3.30x10$^{-3}$) and TS ($r_g$=0.12, p=1.39x10$^{-2}$) at FDR < 0.05. Stroke was also significantly correlated with multiple psychiatric disorders at FDR < 0.05. Stroke was positively correlated with ADHD ($r_g$=0.29, p=6.70x10$^{-13}$), PTSD ($r_g$=0.28, p=1.00x10$^{-4}$), ANX ($r_g$=0.20, p=5.09x10$^{-5}$) MDD ($r_g$=0.16, p=1.22x10$^{-7}$) and BD ($r_g$=0.09, p=9.30x10$^{-3}$), while it was negatively correlated with OCD ($r_g$=-0.19, p=5.90x10$^{-3}$).

Except for negative correlations between CRT-TH and the psychiatric disorders ADHD ($r_g$=-0.19, p=1.67x10$^{-7}$), ANX ($r_g$=-0.14, p=1.60x10$^{-3}$) and MDD ($r_g$=-0.08, p=1.40x10$^{-3}$), neither of the two neuroimaging measures were significantly correlated with neurological or psychiatric disorders at FDR < 0.05. CRT-TH and CRT-SA were negatively correlated with each other ($r_g$=-0.36, p=3.43x10$^{-17}$), while CRT-TH was positively correlated with COG ($r_g$=0.23, p=4.75x10$^{-17}$) and height ($r_g$=0.20, p=6.86x10$^{-16}$), and negatively correlated with CAD ($r_g$=-0.12, p=5.00x10$^{-4}$). Cognitive impairment is a clinical hallmark shared between many psychiatric and neurological conditions[40-43]. As expected[31], COG was negatively correlated with several neurological and psychiatric disorders, but positively correlated with ASD ($r_g$=0.23, p=5.60x10$^{-14}$) and AN ($r_g$=0.09, p=3.60x10$^{-3}$). Notably, COG was also significantly correlated with somatic disorders CAD ($r_g$=-0.19, p=2.14x10$^{-18}$), T2D ($r_g$=-0.05, p=1.22x10$^{-2}$), IBD ($r_g$=0.10, p=2.00x10$^{-4}$) as well as height ($r_g$=0.10, p=2.65x10$^{-16}$), indicating that genetic variants linked to COG influence a wide spectrum of health-related traits. CAD, which was genetically correlated with stroke ($r_g$=0.52, p=1.65x10$^{-33}$), showed a similar pattern of genetic correlations as stroke. Specifically, CAD was significantly correlated with four psychiatric disorders (MDD ($r_g$=0.22, p=4.15x10$^{-24}$), ADHD ($r_g$=0.31, p=5.06x10$^{-26}$), ANX ($r_g$=0.25, p=1.85x10$^{-10}$) and PTSD ($r_g$=0.27, p=9.66x10$^{-6}$) as well as FE ($r_g$=0.20, p=5.60x10$^{-3}$). T2D, a cardiovascular disease risk factor, was also significantly correlated with MDD ($r_g$=0.11, p=2.86x10$^{-10}$), ADHD ($r_g$=0.18, p=4.82x10$^{-11}$) and ANX ($r_g$=0.11, p=2.20x10$^{-3}$), but negatively correlated with AN ($r_g$=-0.10, p=1.40x10$^{-3}$) and OCD ($r_g$=-0.14, p=5.20x10$^{-3}$). Apart from psychiatric disorders, T2D was significantly correlated with stroke ($r_g$=0.27, p=2.92x10$^{-13}$), MS ($r_g$=0.08, p=1.26x10$^{-2}$), and CAD ($r_g$=0.33, p=4.18x10$^{-27}$). IBD was significantly positively correlated with five psychiatric disorders SCZ ($r_g$=0.14, p=9.83x10$^{-9}$), BD ($r_g$=0.09, p=8.00x10$^{-4}$), MDD ($r_g$=0.11, p=6.11x10$^{-7}$), ANX ($r_g$=0.11, p=3.50x10$^{-3}$) and OCD ($r_g$=0.15, p=1.03x10$^{-2}$), but only with MS ($r_g$=0.25, p=4.45x10$^{-9}$) among the neurological disorders. CKD was negatively correlated with four psychiatric disorders SCZ ($r_g$=-0.14, p=4.67x10$^{-5}$), BD ($r_g$=-0.11, p=2.80x10$^{-3}$), PTSD ($r_g$=-0.36, p=1.00x10$^{-4}$), and AN ($r_g$=-0.17, p=3.60x10$^{-3}$), but not with any neurological disorder. Height showed multiple significant genetic correlations at FDR $<$ 0.05. Specifically, height was negatively correlated with MDD ($r_g$=-0.04, p=2.00x10$^{-4}$), ANX ($r_g$=-0.07, p=9.00x10$^{-4}$), PTSD ($r_g$=-0.11, p=6.10x10$^{-3}$), ALZ ($r_g$=-0.09, p=1.80x10$^{-2}$), and CAD ($r_g$=-0.13, p=2.55x10$^{-12}$), and positively correlated with CRT-TH ($r_g$=0.20, p=6.86x10$^{-16}$), COG ($r_g$=-0.10, p=2.65x10$^{-16}$) and CKD ($r_g$=0.10, p=3.00x10$^{-4}$), demonstrating multiple health-related correlations with this anthropometric measure. As observed for other measures of genetic overlap in this study, the presence of statistically significant genetic correlations appeared to be driven by statistical power. The GWAS with low power (e.g., TS and OCD) showed fewer significant correlations than other psychiatric disorders, while this is likely to change as their respective GWAS increase in sample size.

#### 2.2 Differential expression of MAGMA genes in specific tissues

Using MAGMA, we evaluated whether the implicated genes for each GWAS were differentially expressed in 54 specific human tissue types based on Genotype-Tissue Expression v.8 data[44], as implemented in FUMA[45] (Supplementary Fig. 3) Risk genes for ALZ were significantly upregulated in the spleen, the terminal ileum of the small intestine and in whole blood, all linked to the immune system. Risk genes for ADHD were significantly upregulated in the cerebral cortex only. Risk genes for AN were significantly downregulated in the putamen and the hippocampus, and differentially expressed in the putamen, the hippocampus and the amygdala (considering both sides). Genes implicated for CRT-TH were significantly upregulated in the cerebellar hemisphere only. Genes implicated for COG were significantly upregulated in several brain tissues, specifically the anterior cingulate cortex, frontal cortex, cerebral cortex, nucleus accumbens, the cerebellar hemisphere and the cerebellum, and downregulated in the renal cortex. Moreover, COG genes were differentially expressed in the following brain tissues, the putamen, anterior cingulate cortex, hypothalamus, amygdala, hippocampus, frontal cortex, cerebral cortex, nucleus accumbens and the substantia nigra. Height genes were significantly upregulated, downregulated or differentially expressed in 37 out of all 54 tissue types assessed, indicating body-wide expression of height genes. Risk genes for IBD were significantly upregulated in the spleen and whole blood, and downregulated in several brain tissues, specifically the putamen, amygdala, hippocampus, C-1 of the spinal cord, the substantia nigra, hypothalamus and frontal cortex. Risk genes for MDD were significantly upregulated in several brain tissues, specifically the hypothalamus, nucleus accumbens, putamen, hippocampus, the frontal cortex, nucleus caudate, substantia nigra, amygdala, anterior cingulate cortex, cerebral cortex, cerebellum, and the cerebellar hemisphere. Moreover, risk genes for MDD were differentially expressed in the hypothalamus, nucleus accumbens, putamen, hippocampus, and the frontal cortex. Risk genes for MS were significantly upregulated in EBV-transformed lymphocytes, the spleen, whole blood and in the terminal ileum of the small intestine. PD risk genes were significantly upregulated in the substantia nigra only. Risk genes for SCZ were significantly upregulated in several brain tissues, specifically the hippocampus, frontal cortex, cerebellum, the anterior cingulate cortex, the cerebral cortex, the cerebellar hemisphere, the amygdala, and the nucleus accumbens. Furthermore, SCZ risk genes were differentially expressed in the hippocampus, frontal cortex, cerebellum. and the anterior cingulate cortex. Finally, T2D risk genes were significantly downregulated in the substantia nigra, amygdala, hippocampus, C-1 of the spinal cord, putamen, caudate nucleus, nucleus accumbens and the hypothalamus.

### 3 Supplementary Figures

#### 3.1 Supplementary Fig. 1 | GWAS power plots

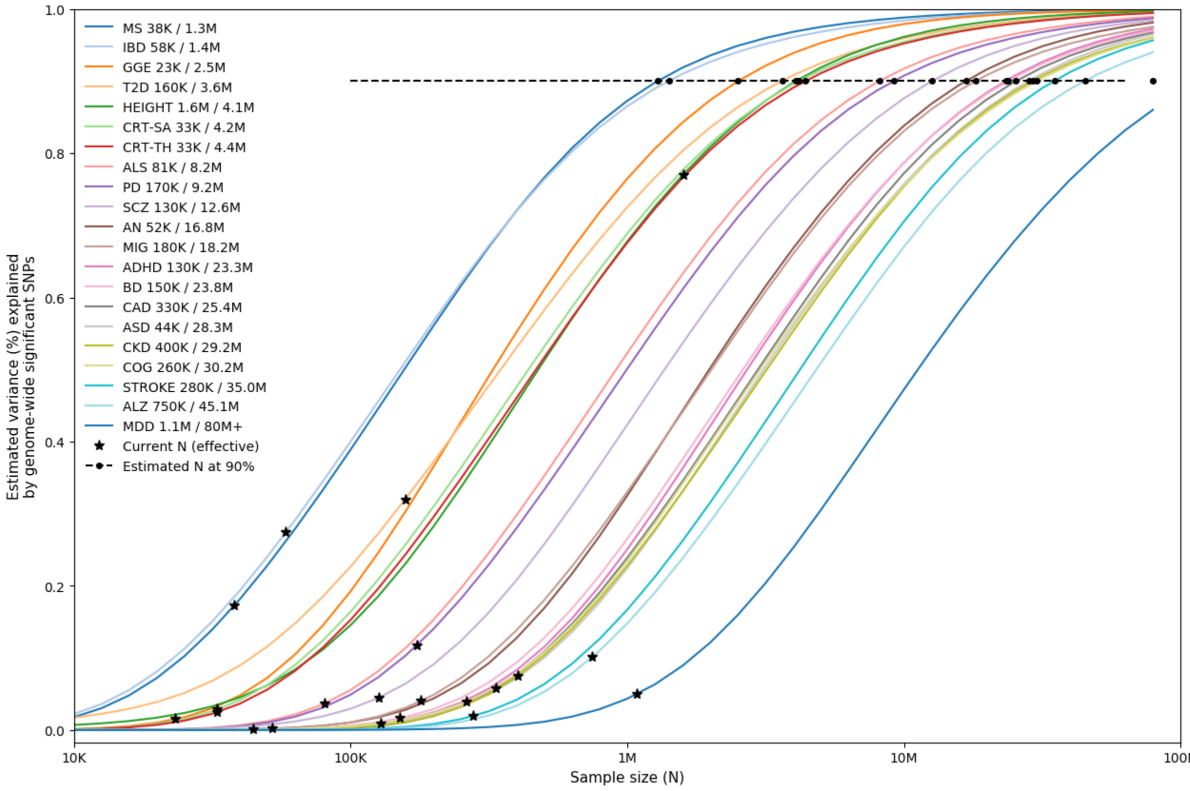

**Supplementary Fig. 1 | GWAS power plots**. Power plots of current and future GWAS estimated using MiXeR[46], displaying the proportion of SNP- heritability explained by genome-wide significant SNPs as a function of sample size, N. Asterisks indicate the current effective sample sizes of the GWAS, and circles indicate the estimated sample sizes needed to capture 90% of the genetic variance for that phenotype.

#### 3.2 Supplementary Fig. 2 | Genetic correlations

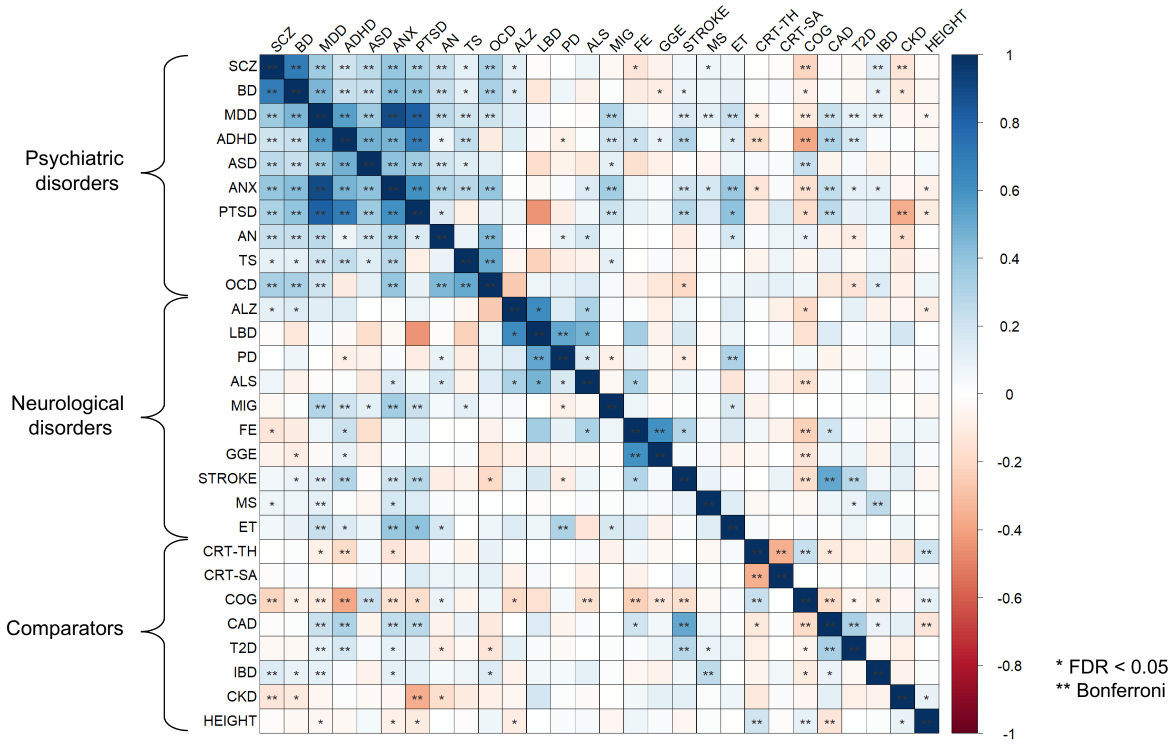

**Supplementary Fig. 2 | Genetic correlations.** Global pairwise genetic correlations across all phenotypes estimated using linkage disequilibrium score regression[39]. One asterisk denotes statistical significance at FDR < 0.05, two asterisks denote statistical significance after Bonferroni correction. The color denotes the magnitude and direction of correlation.

#### 3.3 Supplementary Fig. 3 | Bivariate MiXeR estimates of genetic overlap

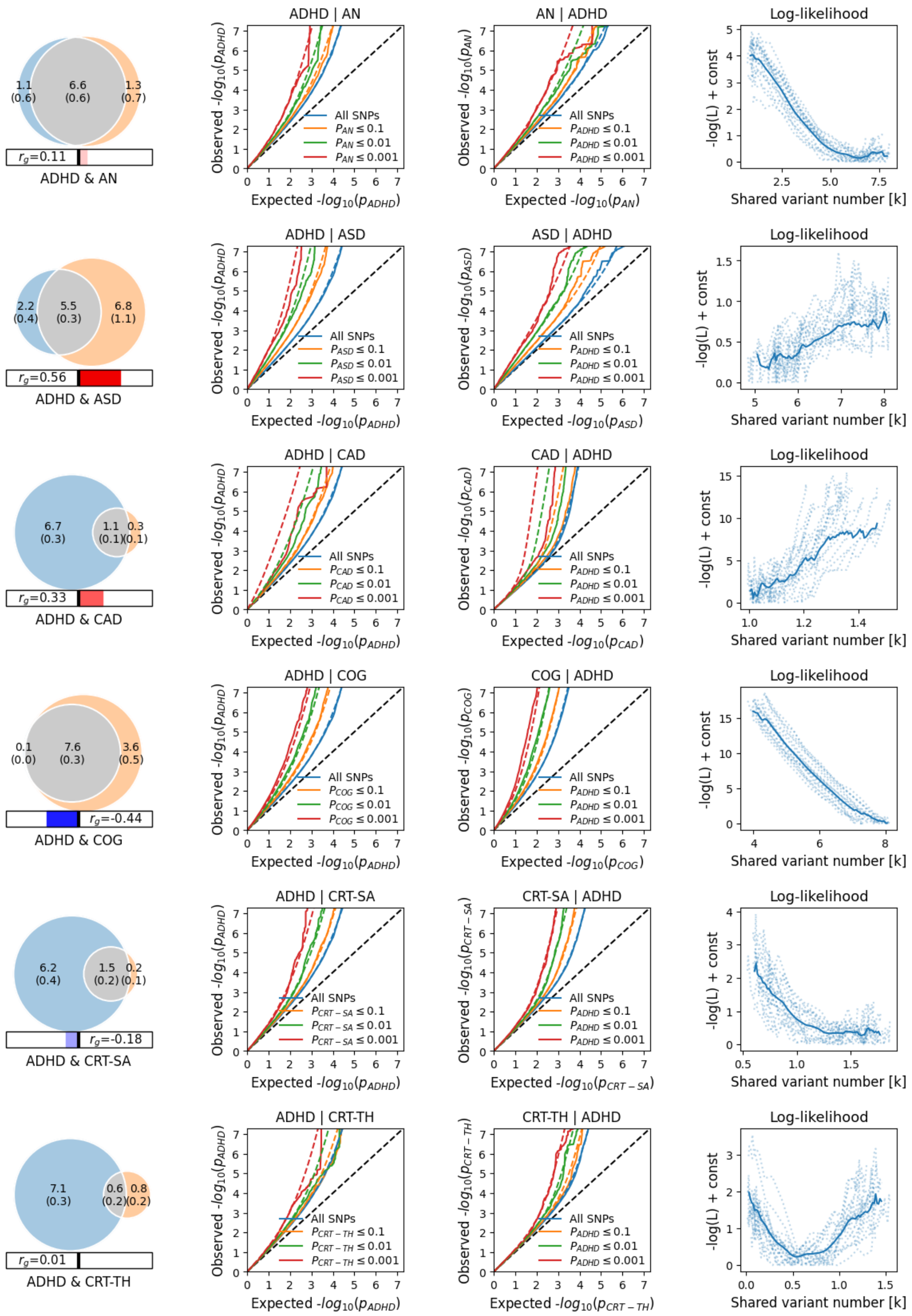

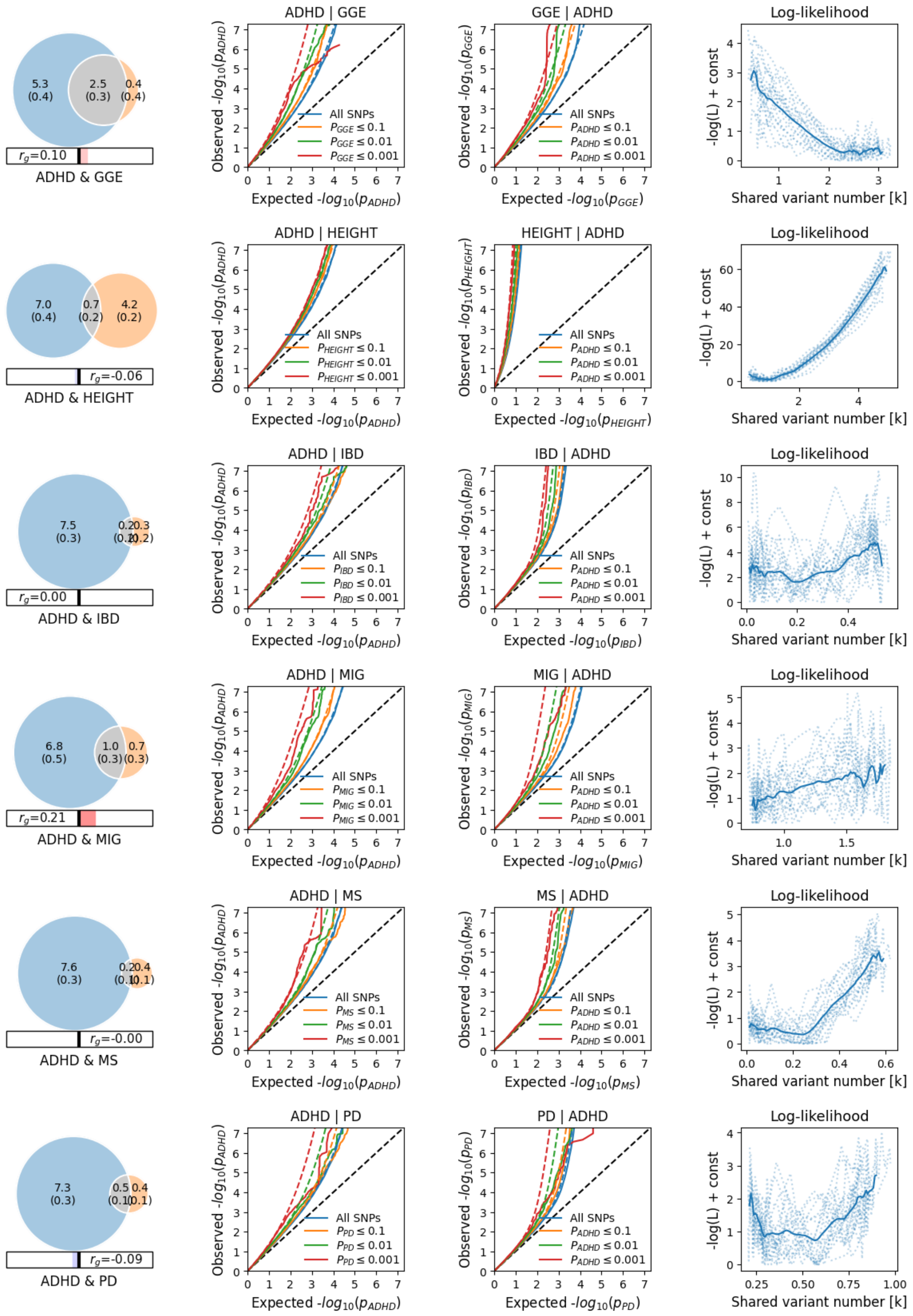
ADHD & GGE
$r_g$=0.10
ADHD | GGE
GGE | ADHD
Log-likelihood
ADHD & HEIGHT
$r_g$=-0.06
ADHD | HEIGHT
HEIGHT | ADHD
ADHD & IBD
$r_g$=0.00
ADHD | IBD
IBD | ADHD
ADHD & MIG
$r_g$=0.21
ADHD | MIG
MIG | ADHD
ADHD & MS
$r_g$=-0.00
ADHD | MS
MS | ADHD
ADHD & PD
$r_g$=-0.09
ADHD | PD
PD | ADHD
All SNPs
Expected $-log_{10}(p_{ADHD})$
Observed $-log_{10}(p_{ADHD})$
-log(L) + const
Shared variant number [k]

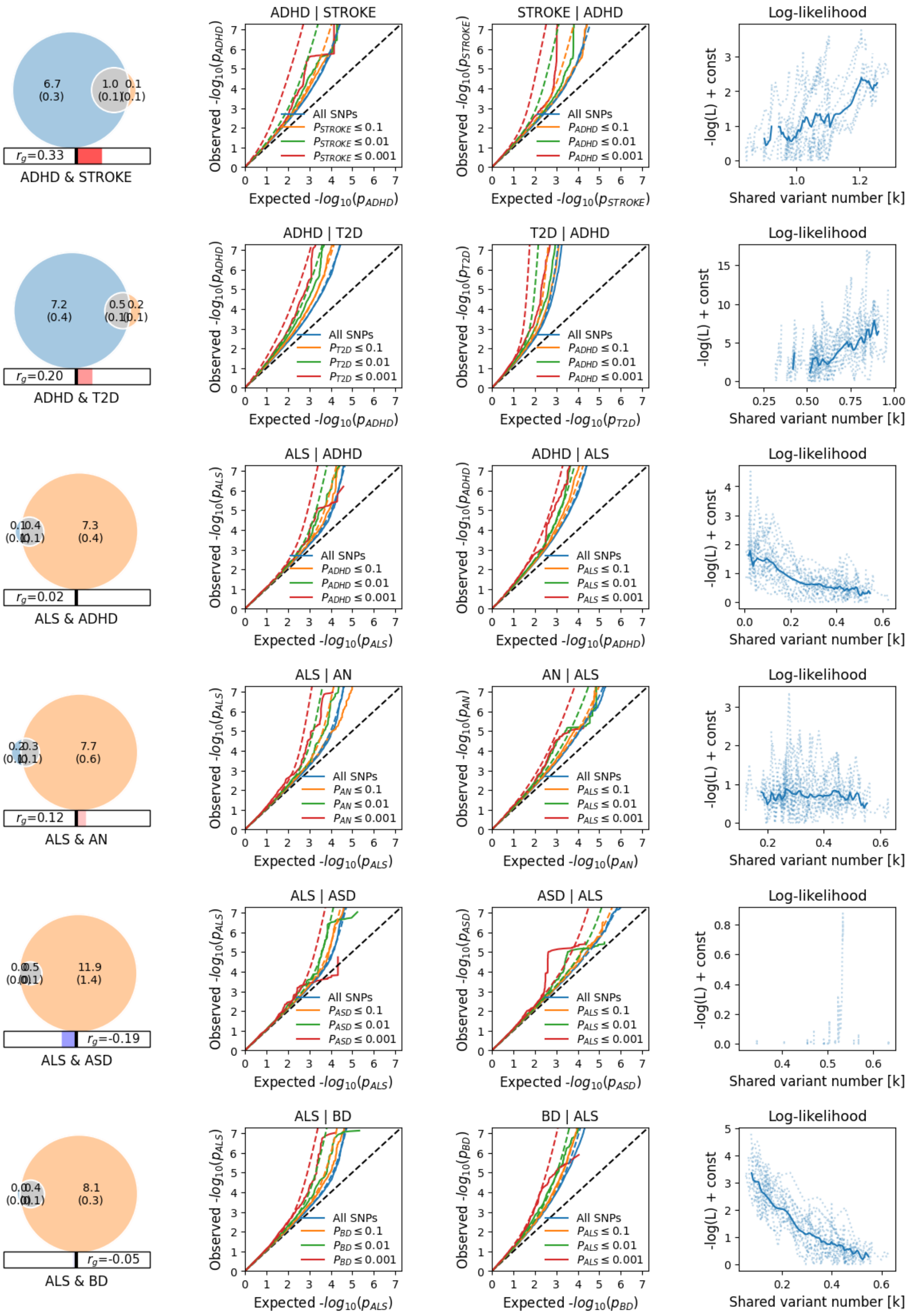
ADHD | STROKE
STROKE | ADHD
Log-likelihood
6.7
(0.3)
1.0
(0.1)
0.1
(0.1)
$r_g$=0.33
ADHD & STROKE
Observed -$log_{10}(p_{ADHD})$
Expected -$log_{10}(p_{ADHD})$
Observed -$log_{10}(p_{STROKE})$
Expected -$log_{10}(p_{STROKE})$
All SNPs
$P_{STROKE} \leq 0.1$
$P_{STROKE} \leq 0.01$
$P_{STROKE} \leq 0.001$
$P_{ADHD} \leq 0.1$
$P_{ADHD} \leq 0.01$
$P_{ADHD} \leq 0.001$
-log(L) + const
Shared variant number [k]
ADHD | T2D
T2D | ADHD
7.2
(0.4)
0.5
(0.1)
0.2
(0.1)
$r_g$=0.20
ADHD & T2D
Observed -$log_{10}(p_{T2D})$
Expected -$log_{10}(p_{T2D})$
$P_{T2D} \leq 0.1$
$P_{T2D} \leq 0.01$
$P_{T2D} \leq 0.001$
ALS | ADHD
ADHD | ALS
0.1
(0.1)
0.4
(0.1)
7.3
(0.4)
$r_g$=0.02
ALS & ADHD
Observed -$log_{10}(p_{ALS})$
Expected -$log_{10}(p_{ALS})$
$P_{ALS} \leq 0.1$
$P_{ALS} \leq 0.01$
$P_{ALS} \leq 0.001$
ALS | AN
AN | ALS
0.2
(0.1)
0.3
(0.1)
7.7
(0.6)
$r_g$=0.12
ALS & AN
Observed -$log_{10}(p_{AN})$
Expected -$log_{10}(p_{AN})$
$P_{AN} \leq 0.1$
$P_{AN} \leq 0.01$
$P_{AN} \leq 0.001$
ALS | ASD
ASD | ALS
0.0
(0.0)
0.5
(0.1)
11.9
(1.4)
$r_g$=-0.19
ALS & ASD
Observed -$log_{10}(p_{ASD})$
Expected -$log_{10}(p_{ASD})$
$P_{ASD} \leq 0.1$
$P_{ASD} \leq 0.01$
$P_{ASD} \leq 0.001$
ALS | BD
BD | ALS
0.0
(0.0)
0.4
(0.1)
8.1
(0.3)
$r_g$=-0.05
ALS & BD
Observed -$log_{10}(p_{BD})$
Expected -$log_{10}(p_{BD})$
$P_{BD} \leq 0.1$
$P_{BD} \leq 0.01$
$P_{BD} \leq 0.001$

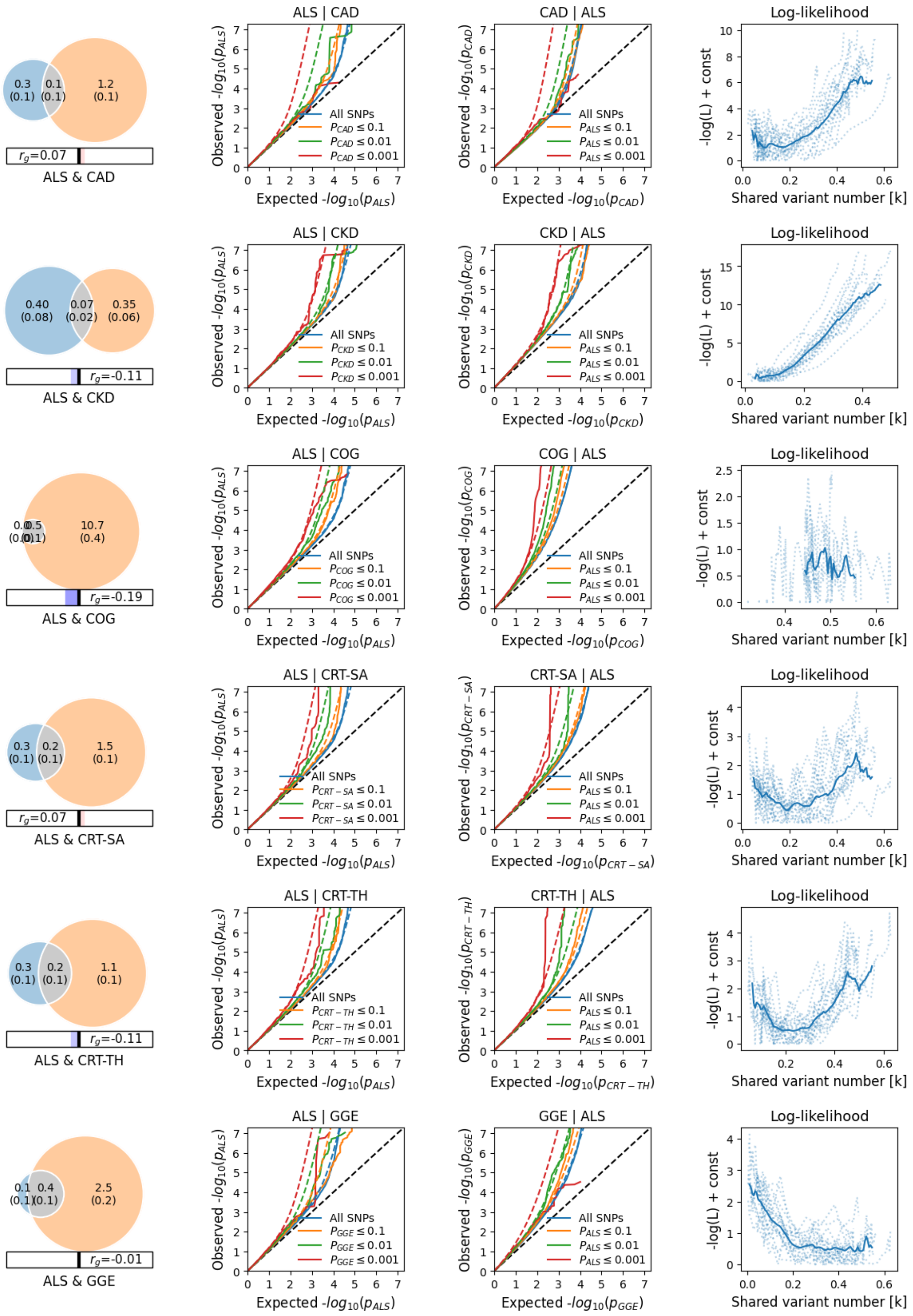

ALS & CAD
$r_g$=0.07
ALS | CAD
CAD | ALS
Log-likelihood
ALS & CKD
$r_g$=-0.11
ALS | CKD
CKD | ALS
ALS & COG
$r_g$=-0.19
ALS | COG
COG | ALS
ALS & CRT-SA
$r_g$=0.07
ALS | CRT-SA
CRT-SA | ALS
ALS & CRT-TH
$r_g$=-0.11
ALS | CRT-TH
CRT-TH | ALS
ALS & GGE
$r_g$=-0.01
ALS | GGE
GGE | ALS
All SNPs
Observed $-log_{10}(p_{ALS})$
Expected $-log_{10}(p_{ALS})$
-log(L) + const
Shared variant number [k]

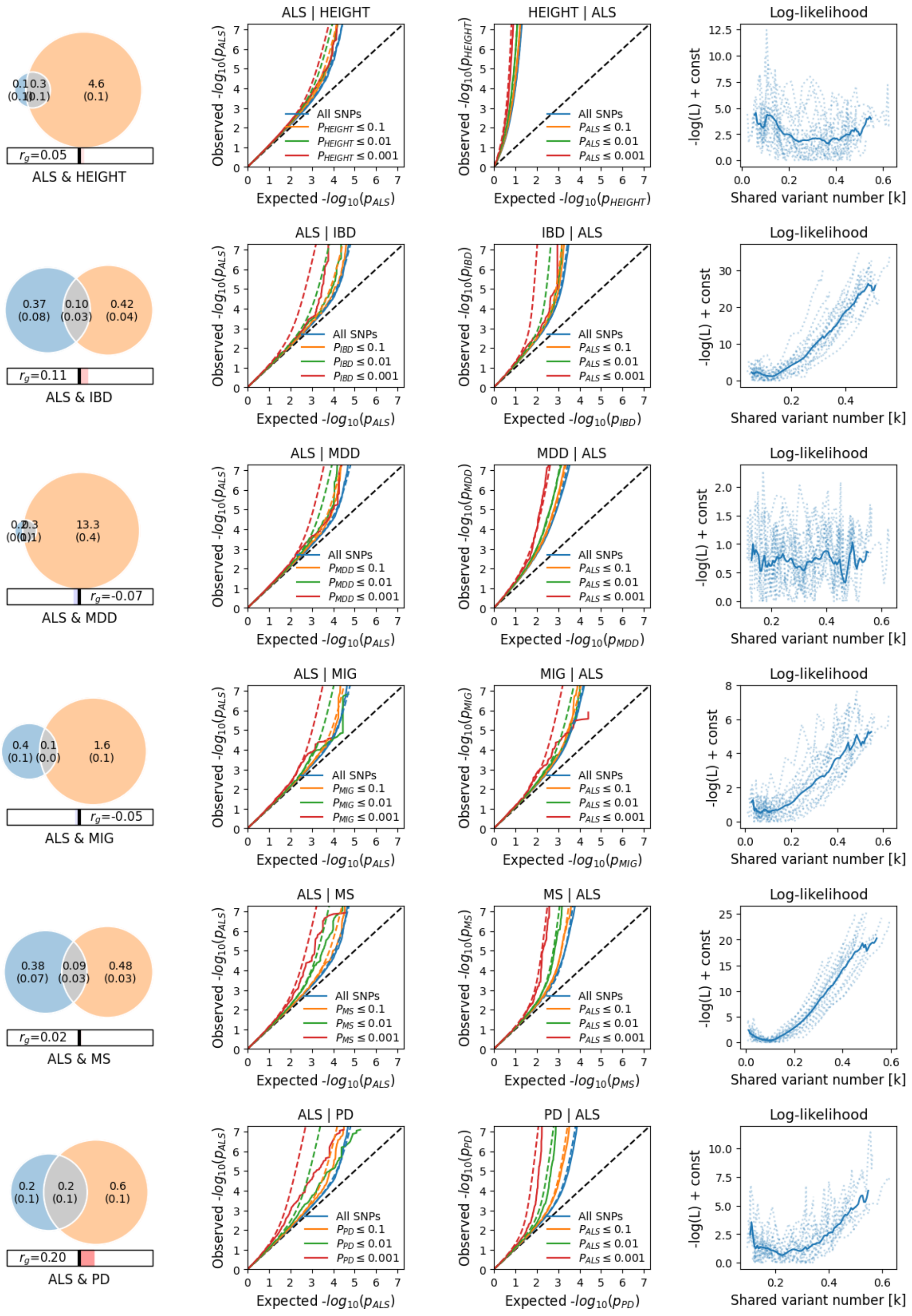

ALS & HEIGHT
$r_g$=0.05
ALS | HEIGHT
HEIGHT | ALS
Log-likelihood
ALS & IBD
$r_g$=0.11
ALS | IBD
IBD | ALS
ALS & MDD
$r_g$=-0.07
ALS | MDD
MDD | ALS
ALS & MIG
$r_g$=-0.05
ALS | MIG
MIG | ALS
ALS & MS
$r_g$=0.02
ALS | MS
MS | ALS
ALS & PD
$r_g$=0.20
ALS | PD
PD | ALS
All SNPs
Expected -$log_{10}(p_{ALS})$
Shared variant number [k]
-log(L) + const

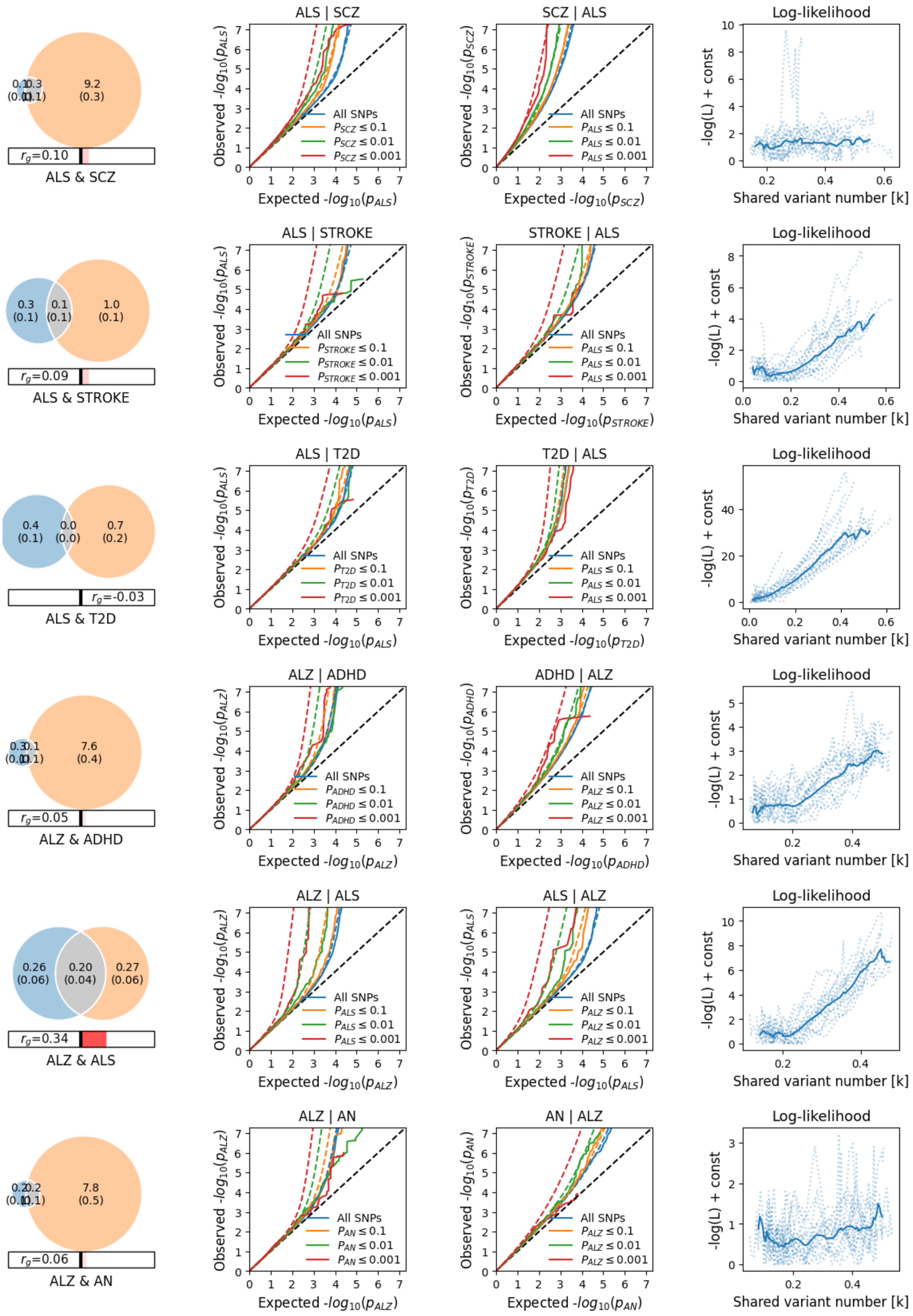
9.2
(0.3)
$r_g$=0.10
ALS & SCZ
ALS | SCZ
SCZ | ALS
Log-likelihood
0.3
(0.1)
0.1
(0.1)
1.0
(0.1)
$r_g$=0.09
ALS & STROKE
ALS | STROKE
STROKE | ALS
0.4
(0.1)
0.0
(0.0)
0.7
(0.2)
$r_g$=-0.03
ALS & T2D
ALS | T2D
T2D | ALS
7.6
(0.4)
$r_g$=0.05
ALZ & ADHD
ALZ | ADHD
ADHD | ALZ
0.26
(0.06)
0.20
(0.04)
0.27
(0.06)
$r_g$=0.34
ALZ & ALS
ALZ | ALS
ALS | ALZ
7.8
(0.5)
$r_g$=0.06
ALZ & AN
ALZ | AN
AN | ALZ
Observed -$log_{10}(p)$
Expected -$log_{10}(p)$
All SNPs
$\le 0.1$
$\le 0.01$
$\le 0.001$
-log(L) + const
Shared variant number [k]

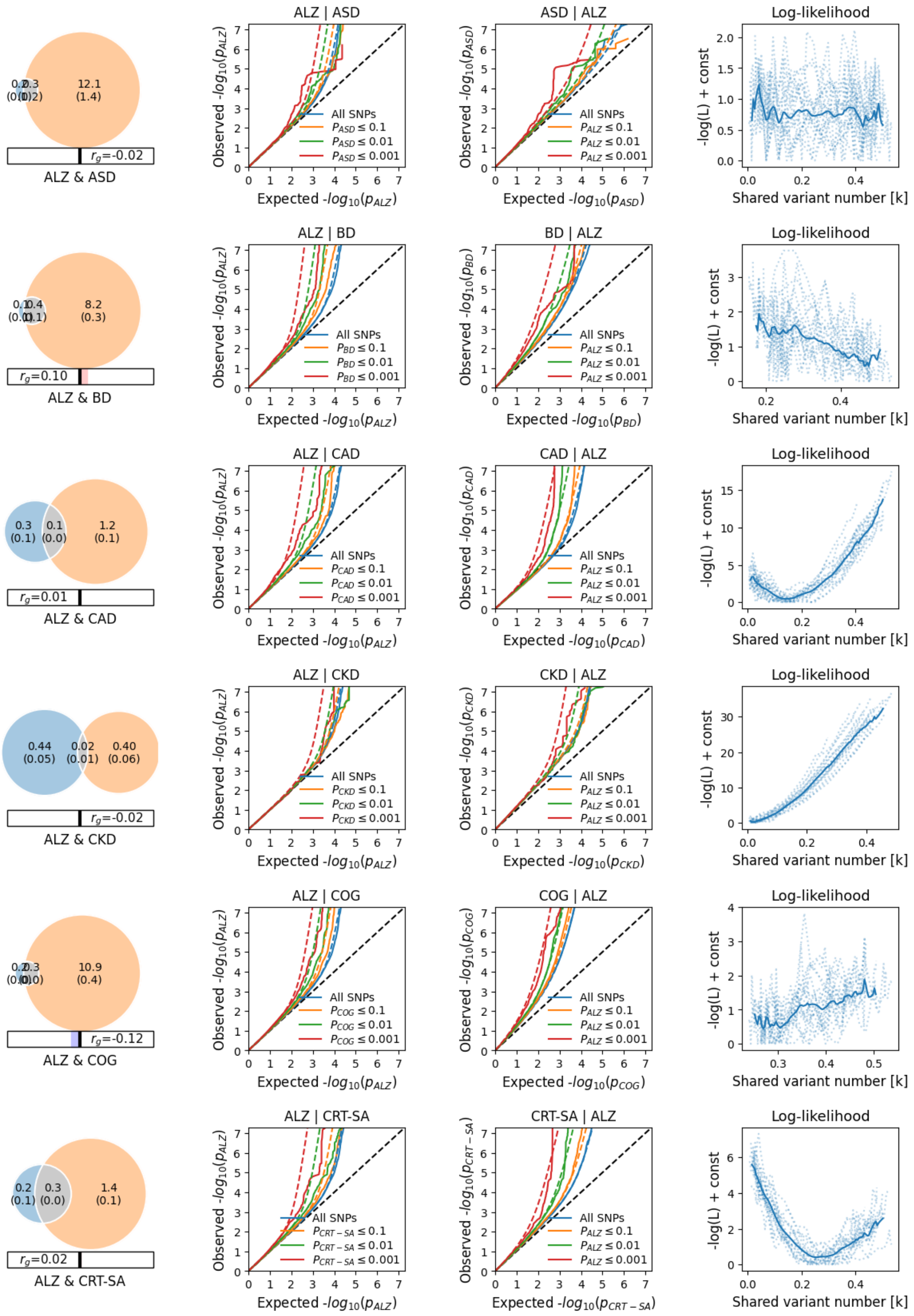
12.1
(1.4)
$r_g$=-0.02
ALZ & ASD
ALZ | ASD
ASD | ALZ
Log-likelihood
8.2
(0.3)
$r_g$=0.10
ALZ & BD
ALZ | BD
BD | ALZ
0.3
(0.1)
0.1
(0.0)
1.2
(0.1)
$r_g$=0.01
ALZ & CAD
ALZ | CAD
CAD | ALZ
0.44
(0.05)
0.02
(0.01)
0.40
(0.06)
$r_g$=-0.02
ALZ & CKD
ALZ | CKD
CKD | ALZ
10.9
(0.4)
$r_g$=-0.12
ALZ & COG
ALZ | COG
COG | ALZ
0.2
(0.1)
0.3
(0.0)
1.4
(0.1)
$r_g$=0.02
ALZ & CRT-SA
ALZ | CRT-SA
CRT-SA | ALZ
All SNPs
Observed $-log_{10}(p_{ALZ})$
Expected $-log_{10}(p_{ALZ})$
-log(L) + const
Shared variant number [k]

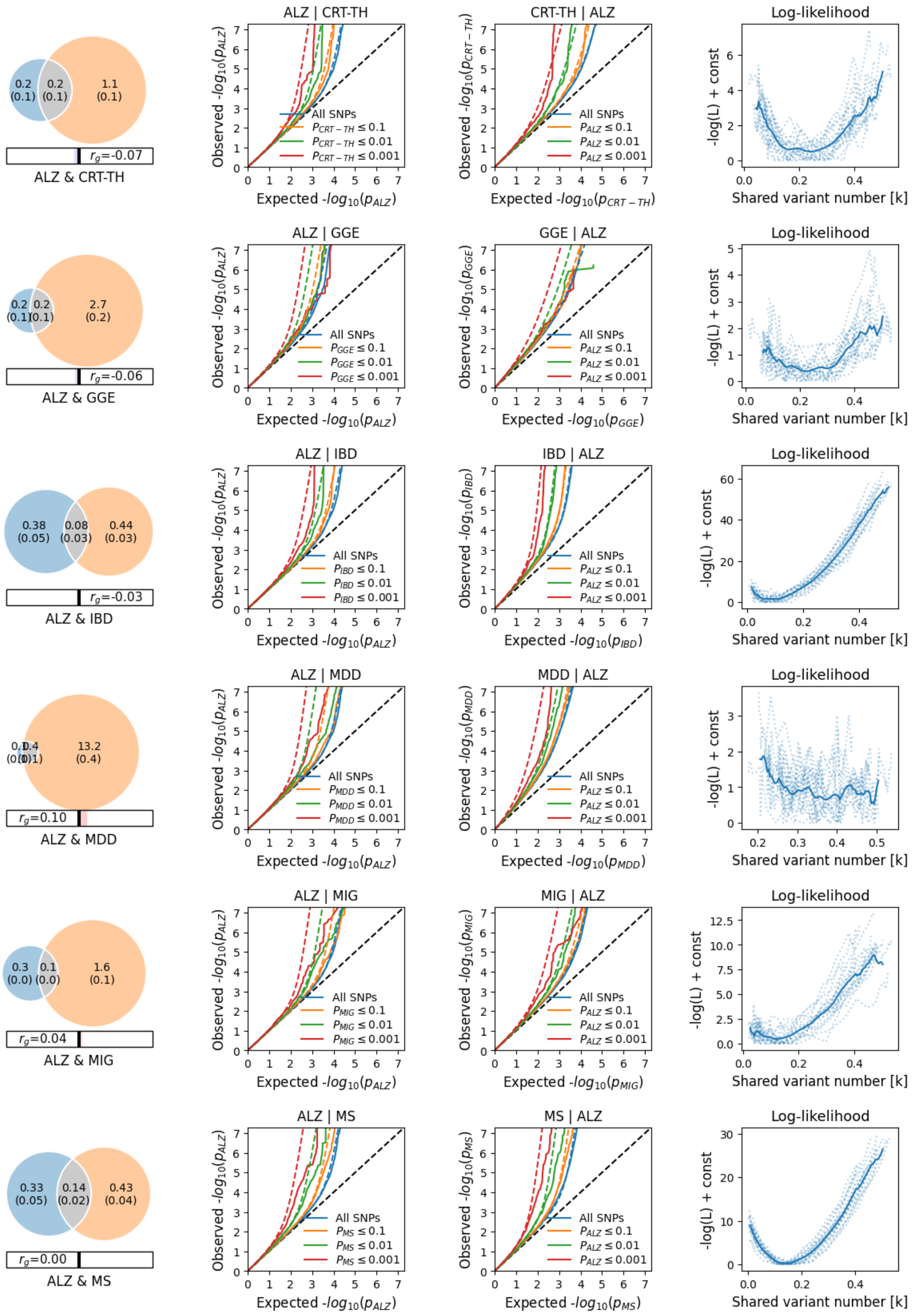
ALZ | CRT-TH
CRT-TH | ALZ
Log-likelihood
ALZ & CRT-TH
$r_g$=-0.07
ALZ | GGE
GGE | ALZ
ALZ & GGE
$r_g$=-0.06
ALZ | IBD
IBD | ALZ
ALZ & IBD
$r_g$=-0.03
ALZ | MDD
MDD | ALZ
ALZ & MDD
$r_g$=0.10
ALZ | MIG
MIG | ALZ
ALZ & MIG
$r_g$=0.04
ALZ | MS
MS | ALZ
ALZ & MS
$r_g$=0.00
All SNPs
Observed -$log_{10}(p_{ALZ})$
Expected -$log_{10}(p_{ALZ})$
-log(L) + const
Shared variant number [k]

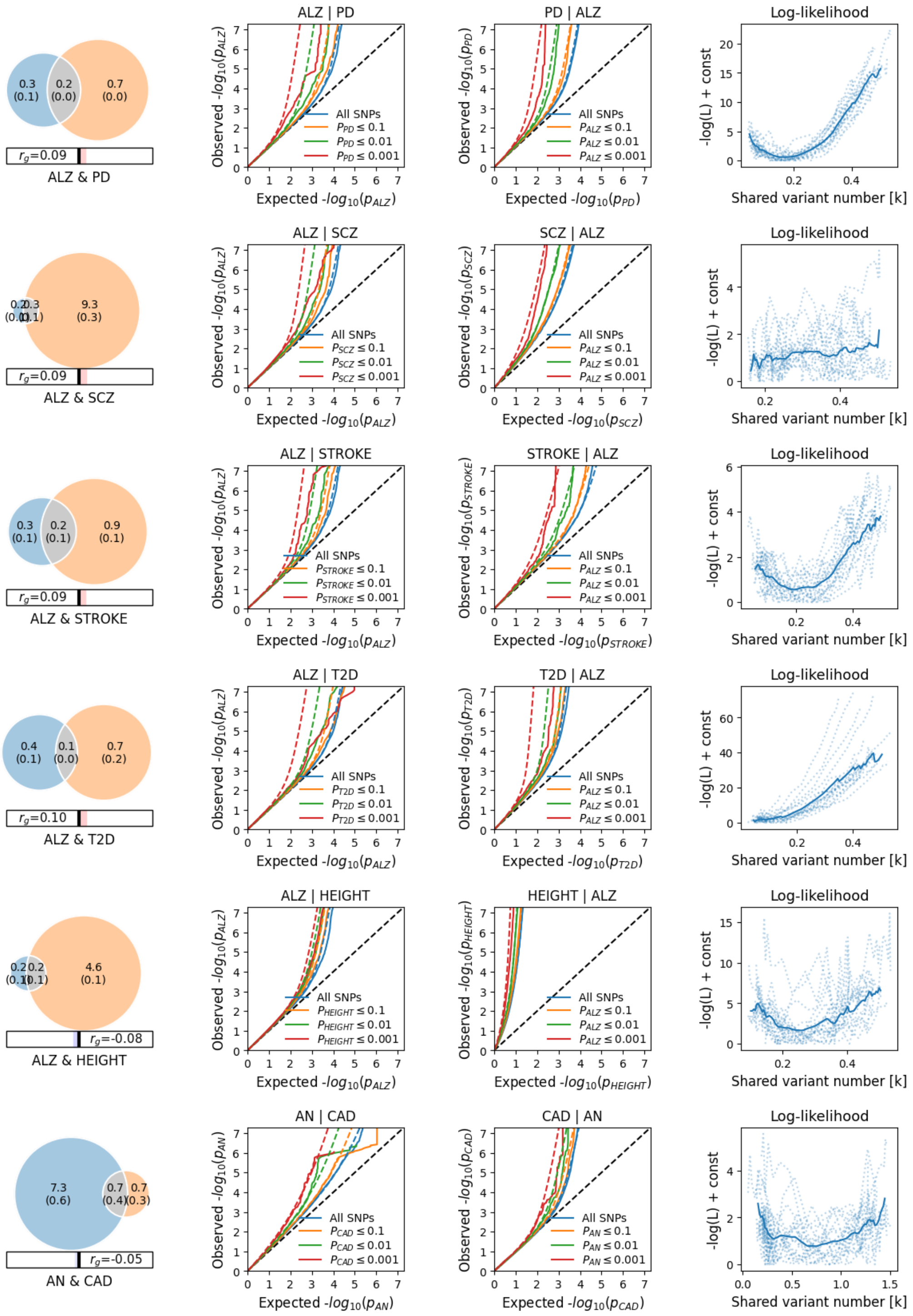

0.3 (0.1)
0.2 (0.0)
0.7 (0.0)
$r_g$=0.09
ALZ & PD
ALZ | PD
Observed -$log_{10}(p_{ALZ})$
Expected -$log_{10}(p_{ALZ})$
All SNPs
$P_{PD} \leq 0.1$
$P_{PD} \leq 0.01$
$P_{PD} \leq 0.001$
PD | ALZ
Observed -$log_{10}(p_{PD})$
Expected -$log_{10}(p_{PD})$
All SNPs
$P_{ALZ} \leq 0.1$
$P_{ALZ} \leq 0.01$
$P_{ALZ} \leq 0.001$
Log-likelihood
-log(L) + const
Shared variant number [k]
0.2 0.3
(0.1) (0.1)
9.3 (0.3)
$r_g$=0.09
ALZ & SCZ
ALZ | SCZ
$P_{SCZ} \leq 0.1$
$P_{SCZ} \leq 0.01$
$P_{SCZ} \leq 0.001$
SCZ | ALZ
Observed -$log_{10}(p_{SCZ})$
Expected -$log_{10}(p_{SCZ})$
0.3 (0.1)
0.2 (0.1)
0.9 (0.1)
$r_g$=0.09
ALZ & STROKE
ALZ | STROKE
$P_{STROKE} \leq 0.1$
$P_{STROKE} \leq 0.01$
$P_{STROKE} \leq 0.001$
STROKE | ALZ
Observed -$log_{10}(p_{STROKE})$
Expected -$log_{10}(p_{STROKE})$
0.4 (0.1)
0.1 (0.0)
0.7 (0.2)
$r_g$=0.10
ALZ & T2D
ALZ | T2D
$P_{T2D} \leq 0.1$
$P_{T2D} \leq 0.01$
$P_{T2D} \leq 0.001$
T2D | ALZ
Observed -$log_{10}(p_{T2D})$
Expected -$log_{10}(p_{T2D})$
0.2 0.2
(0.1) (0.1)
4.6 (0.1)
$r_g$=-0.08
ALZ & HEIGHT
ALZ | HEIGHT
$P_{HEIGHT} \leq 0.1$
$P_{HEIGHT} \leq 0.01$
$P_{HEIGHT} \leq 0.001$
HEIGHT | ALZ
Observed -$log_{10}(p_{HEIGHT})$
Expected -$log_{10}(p_{HEIGHT})$
7.3 (0.6)
0.7 (0.4)
0.7 (0.3)
$r_g$=-0.05
AN & CAD
AN | CAD
Observed -$log_{10}(p_{AN})$
Expected -$log_{10}(p_{AN})$
$P_{CAD} \leq 0.1$
$P_{CAD} \leq 0.01$
$P_{CAD} \leq 0.001$
CAD | AN
Observed -$log_{10}(p_{CAD})$
Expected -$log_{10}(p_{CAD})$
$P_{AN} \leq 0.1$
$P_{AN} \leq 0.01$
$P_{AN} \leq 0.001$

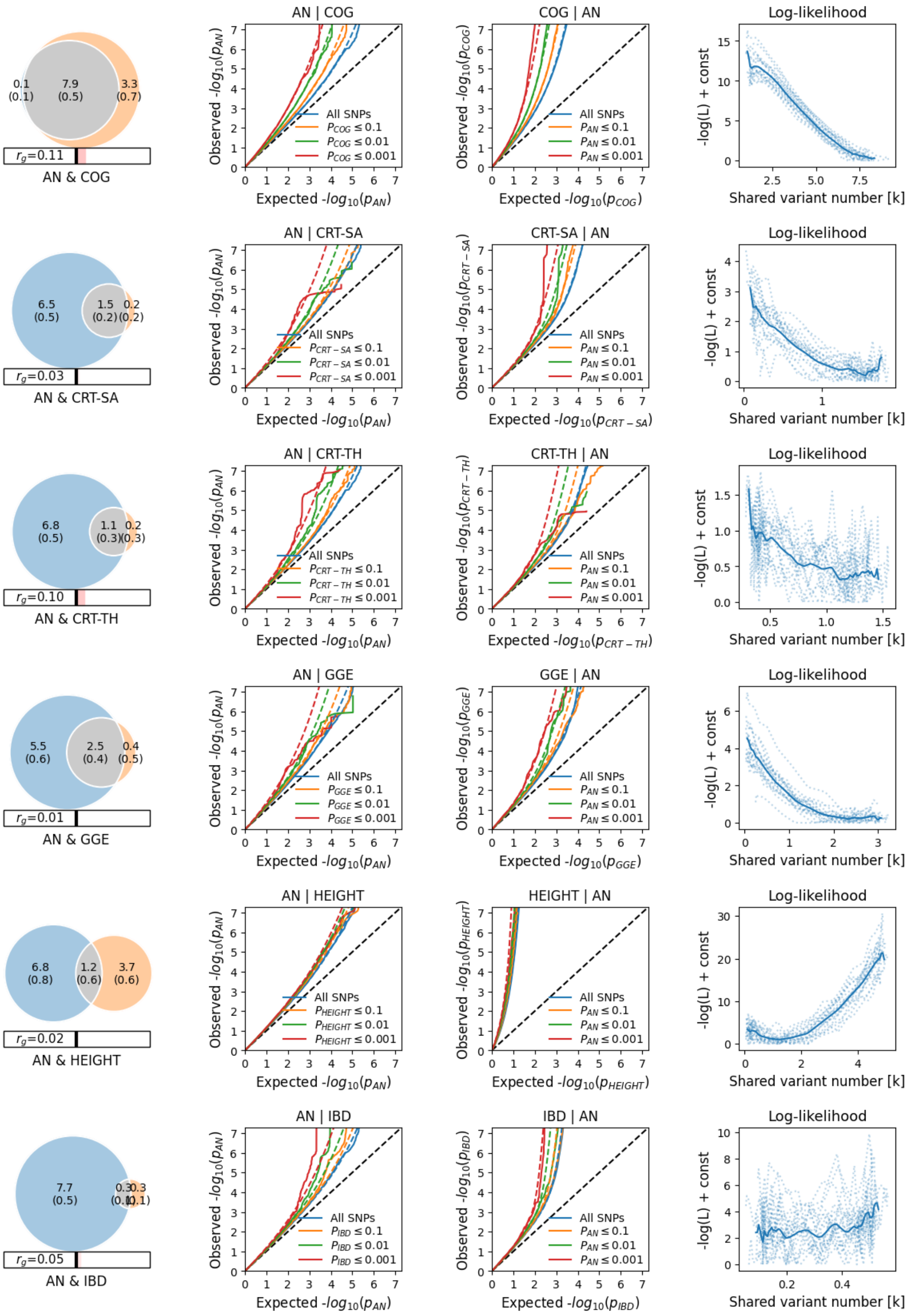
0.1 (0.1)
7.9 (0.5)
3.3 (0.7)
$r_g$=0.11
AN & COG
AN | COG
Observed -$log_{10}(p_{AN})$
Expected -$log_{10}(p_{AN})$
All SNPs
$P_{COG} \leq 0.1$
$P_{COG} \leq 0.01$
$P_{COG} \leq 0.001$
COG | AN
Observed -$log_{10}(p_{COG})$
Expected -$log_{10}(p_{COG})$
$P_{AN} \leq 0.1$
$P_{AN} \leq 0.01$
$P_{AN} \leq 0.001$
Log-likelihood
-log(L) + const
Shared variant number [k]
6.5 (0.5)
1.5 (0.2)
0.2 (0.2)
$r_g$=0.03
AN & CRT-SA
AN | CRT-SA
$P_{CRT-SA} \leq 0.1$
$P_{CRT-SA} \leq 0.01$
$P_{CRT-SA} \leq 0.001$
CRT-SA | AN
Observed -$log_{10}(p_{CRT-SA})$
Expected -$log_{10}(p_{CRT-SA})$
6.8 (0.5)
1.1 (0.3)
0.2 (0.3)
$r_g$=0.10
AN & CRT-TH
AN | CRT-TH
$P_{CRT-TH} \leq 0.1$
$P_{CRT-TH} \leq 0.01$
$P_{CRT-TH} \leq 0.001$
CRT-TH | AN
Observed -$log_{10}(p_{CRT-TH})$
Expected -$log_{10}(p_{CRT-TH})$
5.5 (0.6)
2.5 (0.4)
0.4 (0.5)
$r_g$=0.01
AN & GGE
AN | GGE
$P_{GGE} \leq 0.1$
$P_{GGE} \leq 0.01$
$P_{GGE} \leq 0.001$
GGE | AN
Observed -$log_{10}(p_{GGE})$
Expected -$log_{10}(p_{GGE})$
6.8 (0.8)
1.2 (0.6)
3.7 (0.6)
$r_g$=0.02
AN & HEIGHT
AN | HEIGHT
$P_{HEIGHT} \leq 0.1$
$P_{HEIGHT} \leq 0.01$
$P_{HEIGHT} \leq 0.001$
HEIGHT | AN
Observed -$log_{10}(p_{HEIGHT})$
Expected -$log_{10}(p_{HEIGHT})$
7.7 (0.5)
0.3 (0.1)
0.3 (0.1)
$r_g$=0.05
AN & IBD
AN | IBD
$P_{IBD} \leq 0.1$
$P_{IBD} \leq 0.01$
$P_{IBD} \leq 0.001$
IBD | AN
Observed -$log_{10}(p_{IBD})$
Expected -$log_{10}(p_{IBD})$

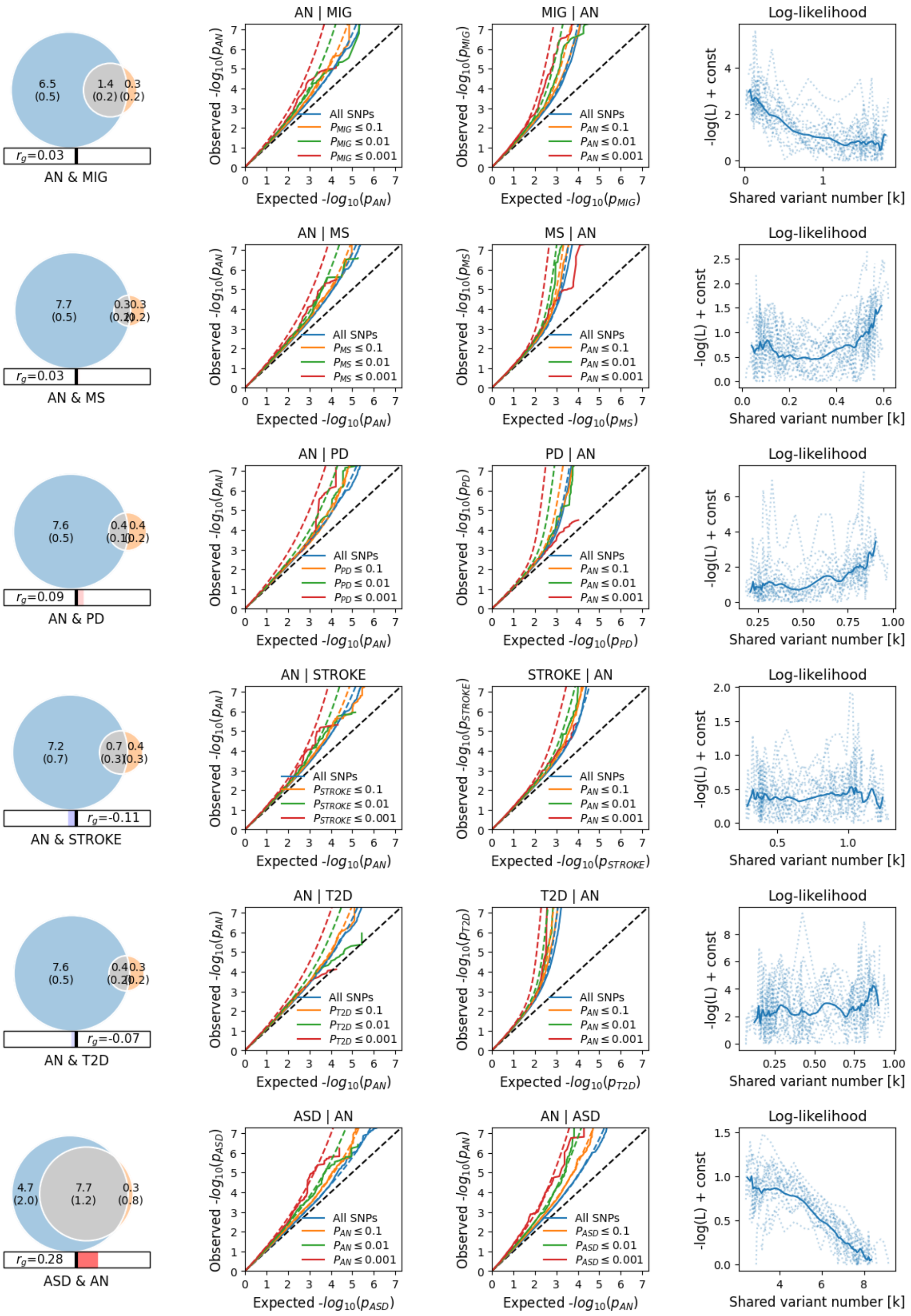
6.5 (0.5)
1.4 (0.2)
0.3 (0.2)
$r_g$=0.03
AN & MIG
AN | MIG
MIG | AN
Log-likelihood
Observed -$log_{10}(p_{AN})$
Expected -$log_{10}(p_{AN})$
Observed -$log_{10}(p_{MIG})$
Expected -$log_{10}(p_{MIG})$
All SNPs
$P_{MIG} \leq 0.1$
$P_{MIG} \leq 0.01$
$P_{MIG} \leq 0.001$
$P_{AN} \leq 0.1$
$P_{AN} \leq 0.01$
$P_{AN} \leq 0.001$
-log(L) + const
Shared variant number [k]
7.7 (0.5)
$r_g$=0.03
AN & MS
AN | MS
MS | AN
Observed -$log_{10}(p_{MS})$
Expected -$log_{10}(p_{MS})$
$P_{MS} \leq 0.1$
$P_{MS} \leq 0.01$
$P_{MS} \leq 0.001$
7.6 (0.5)
0.4 (0.1)
0.4 (0.2)
$r_g$=0.09
AN & PD
AN | PD
PD | AN
Observed -$log_{10}(p_{PD})$
Expected -$log_{10}(p_{PD})$
$P_{PD} \leq 0.1$
$P_{PD} \leq 0.01$
$P_{PD} \leq 0.001$
7.2 (0.7)
0.7 (0.3)
0.4 (0.3)
$r_g$=-0.11
AN & STROKE
AN | STROKE
STROKE | AN
Observed -$log_{10}(p_{STROKE})$
Expected -$log_{10}(p_{STROKE})$
$P_{STROKE} \leq 0.1$
$P_{STROKE} \leq 0.01$
$P_{STROKE} \leq 0.001$
7.6 (0.5)
$r_g$=-0.07
AN & T2D
AN | T2D
T2D | AN
Observed -$log_{10}(p_{T2D})$
Expected -$log_{10}(p_{T2D})$
$P_{T2D} \leq 0.1$
$P_{T2D} \leq 0.01$
$P_{T2D} \leq 0.001$
4.7 (2.0)
7.7 (1.2)
0.3 (0.8)
$r_g$=0.28
ASD & AN
ASD | AN
AN | ASD
Observed -$log_{10}(p_{ASD})$
Expected -$log_{10}(p_{ASD})$
$P_{ASD} \leq 0.1$
$P_{ASD} \leq 0.01$
$P_{ASD} \leq 0.001$

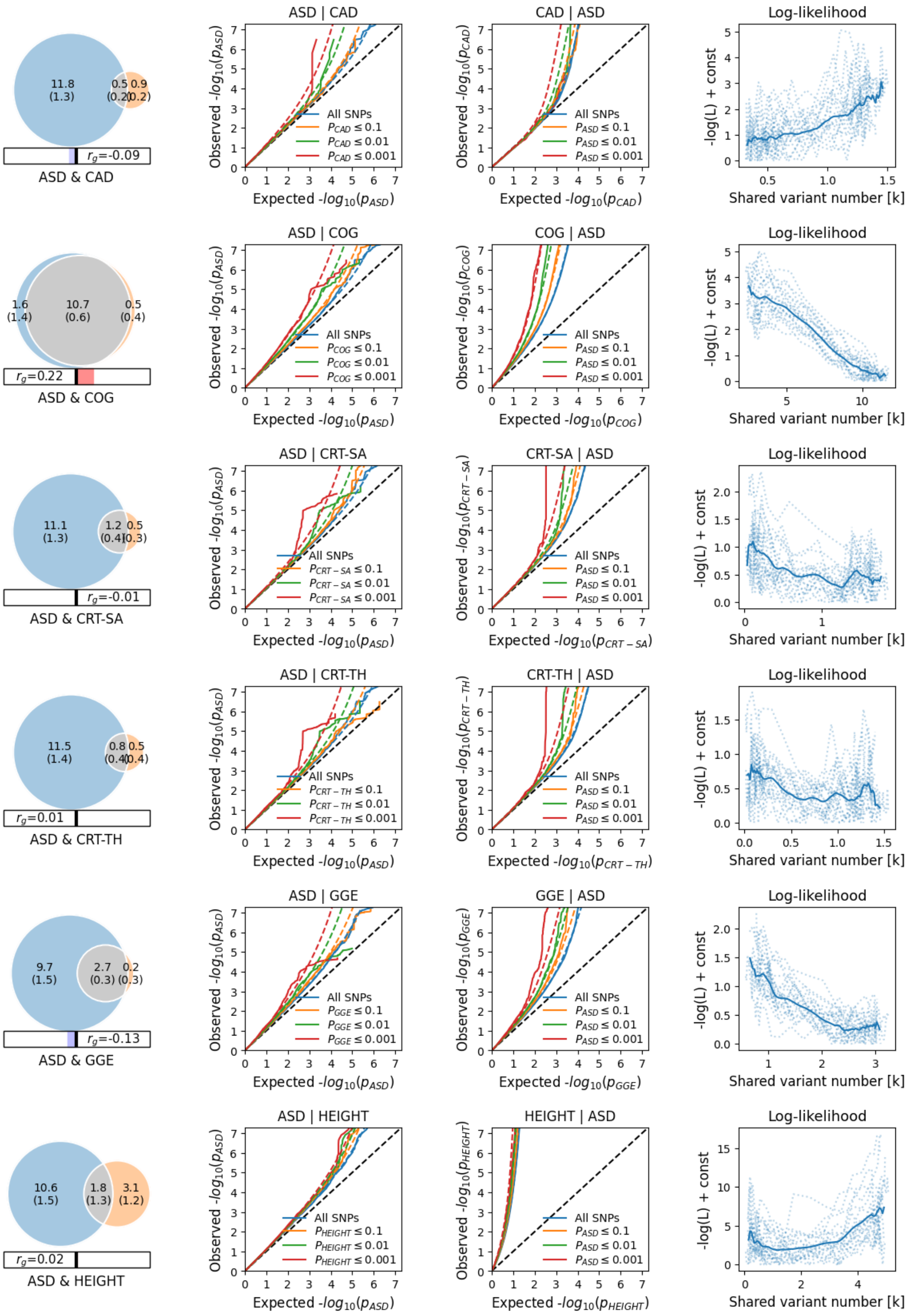
ASD & CAD
$r_g$=-0.09
ASD | CAD
CAD | ASD
Log-likelihood
ASD & COG
$r_g$=0.22
ASD | COG
COG | ASD
ASD & CRT-SA
$r_g$=-0.01
ASD | CRT-SA
CRT-SA | ASD
ASD & CRT-TH
$r_g$=0.01
ASD | CRT-TH
CRT-TH | ASD
ASD & GGE
$r_g$=-0.13
ASD | GGE
GGE | ASD
ASD & HEIGHT
$r_g$=0.02
ASD | HEIGHT
HEIGHT | ASD
All SNPs
Expected $-log_{10}(p_{ASD})$
Observed $-log_{10}(p_{ASD})$
-log(L) + const
Shared variant number [k]

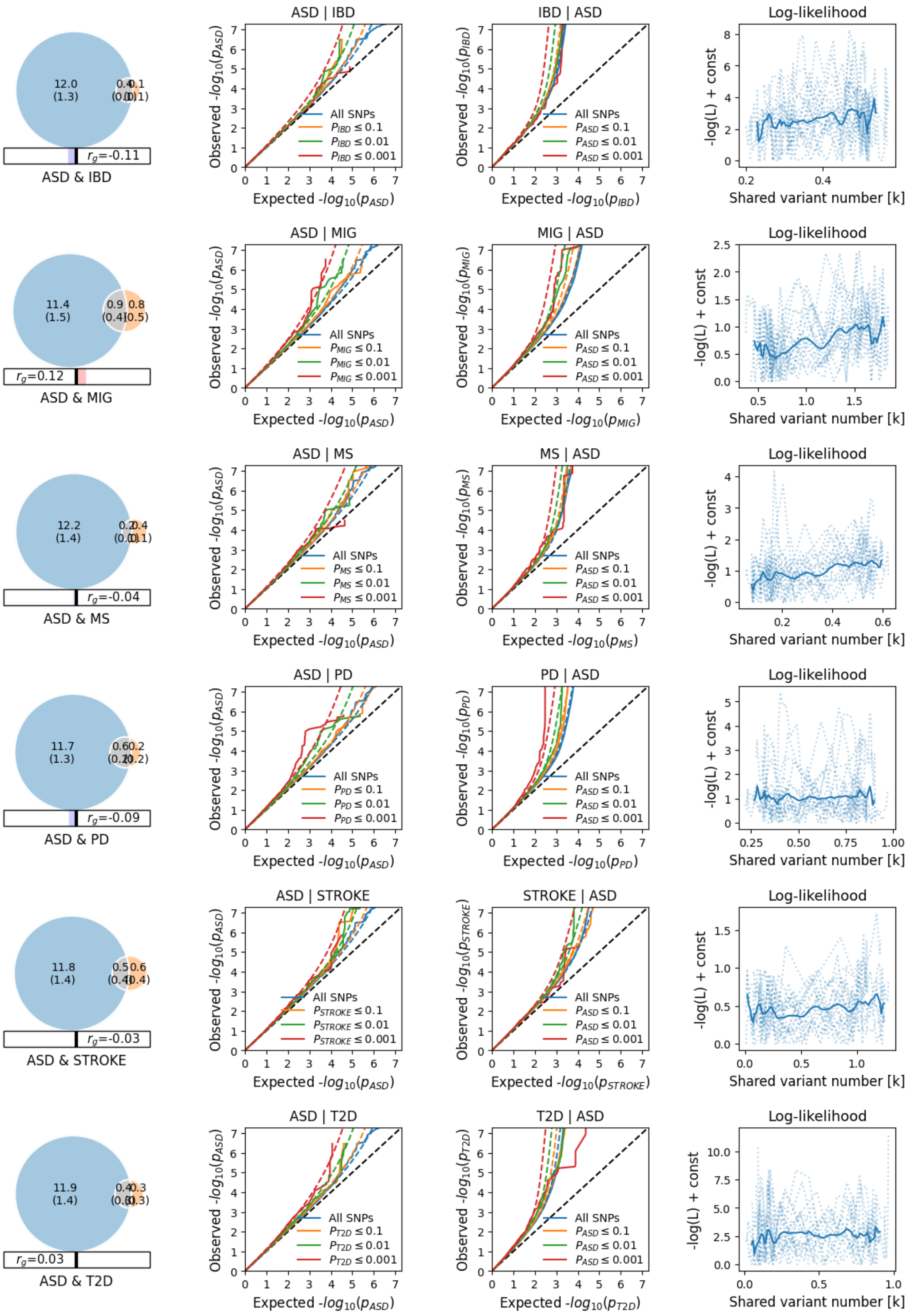

12.0 (1.3)
0.4 (0.0)
0.1 (0.1)
$r_g$=-0.11
ASD & IBD
ASD | IBD
IBD | ASD
Log-likelihood
Observed -$log_{10}(p_{ASD})$
Expected -$log_{10}(p_{ASD})$
Observed -$log_{10}(p_{IBD})$
Expected -$log_{10}(p_{IBD})$
All SNPs
$P_{IBD} \leq 0.1$
$P_{IBD} \leq 0.01$
$P_{IBD} \leq 0.001$
$P_{ASD} \leq 0.1$
$P_{ASD} \leq 0.01$
$P_{ASD} \leq 0.001$
-log(L) + const
Shared variant number [k]
11.4 (1.5)
0.9 (0.4)
0.8 (0.5)
$r_g$=0.12
ASD & MIG
ASD | MIG
MIG | ASD
Observed -$log_{10}(p_{MIG})$
Expected -$log_{10}(p_{MIG})$
$P_{MIG} \leq 0.1$
$P_{MIG} \leq 0.01$
$P_{MIG} \leq 0.001$
12.2 (1.4)
0.2 (0.0)
0.4 (0.1)
$r_g$=-0.04
ASD & MS
ASD | MS
MS | ASD
Observed -$log_{10}(p_{MS})$
Expected -$log_{10}(p_{MS})$
$P_{MS} \leq 0.1$
$P_{MS} \leq 0.01$
$P_{MS} \leq 0.001$
11.7 (1.3)
0.6 (0.2)
0.2 (0.2)
$r_g$=-0.09
ASD & PD
ASD | PD
PD | ASD
Observed -$log_{10}(p_{PD})$
Expected -$log_{10}(p_{PD})$
$P_{PD} \leq 0.1$
$P_{PD} \leq 0.01$
$P_{PD} \leq 0.001$
11.8 (1.4)
0.5 (0.4)
0.6 (0.4)
$r_g$=-0.03
ASD & STROKE
ASD | STROKE
STROKE | ASD
Observed -$log_{10}(p_{STROKE})$
Expected -$log_{10}(p_{STROKE})$
$P_{STROKE} \leq 0.1$
$P_{STROKE} \leq 0.01$
$P_{STROKE} \leq 0.001$
11.9 (1.4)
0.4 (0.3)
0.3 (0.3)
$r_g$=0.03
ASD & T2D
ASD | T2D
T2D | ASD
Observed -$log_{10}(p_{T2D})$
Expected -$log_{10}(p_{T2D})$
$P_{T2D} \leq 0.1$
$P_{T2D} \leq 0.01$
$P_{T2D} \leq 0.001$

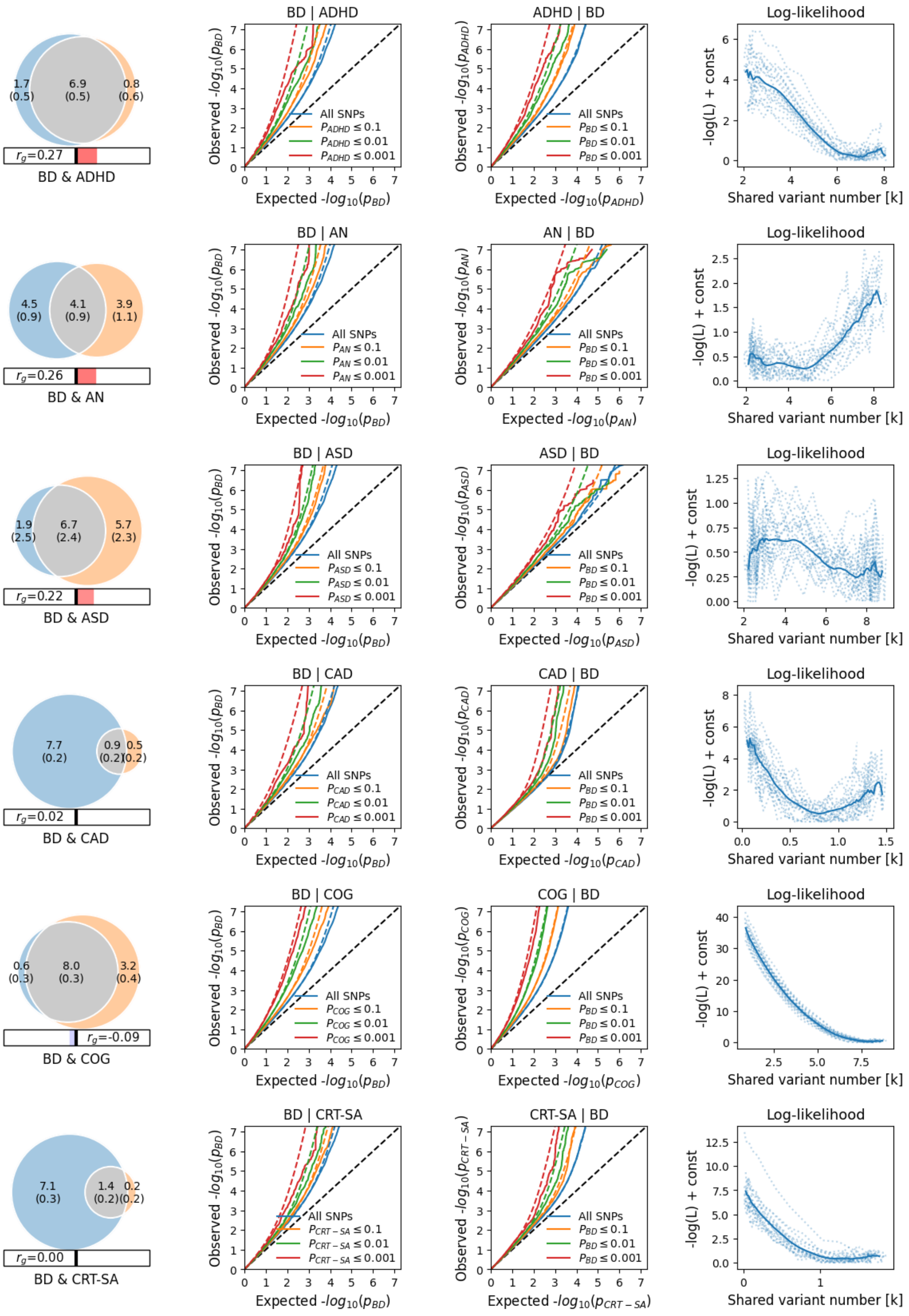
1.7 (0.5)
6.9 (0.5)
0.8 (0.6)
$r_g$=0.27
BD & ADHD
BD | ADHD
ADHD | BD
Log-likelihood
4.5 (0.9)
4.1 (0.9)
3.9 (1.1)
$r_g$=0.26
BD & AN
BD | AN
AN | BD
1.9 (2.5)
6.7 (2.4)
5.7 (2.3)
$r_g$=0.22
BD & ASD
BD | ASD
ASD | BD
7.7 (0.2)
0.9 (0.2)
0.5 (0.2)
$r_g$=0.02
BD & CAD
BD | CAD
CAD | BD
0.6 (0.3)
8.0 (0.3)
3.2 (0.4)
$r_g$=-0.09
BD & COG
BD | COG
COG | BD
7.1 (0.3)
1.4 (0.2)
0.2 (0.2)
$r_g$=0.00
BD & CRT-SA
BD | CRT-SA
CRT-SA | BD
All SNPs
$P_{BD} \leq 0.1$
$P_{BD} \leq 0.01$
$P_{BD} \leq 0.001$
Observed $-log_{10}(p_{BD})$
Expected $-log_{10}(p_{BD})$
-log(L) + const
Shared variant number [k]

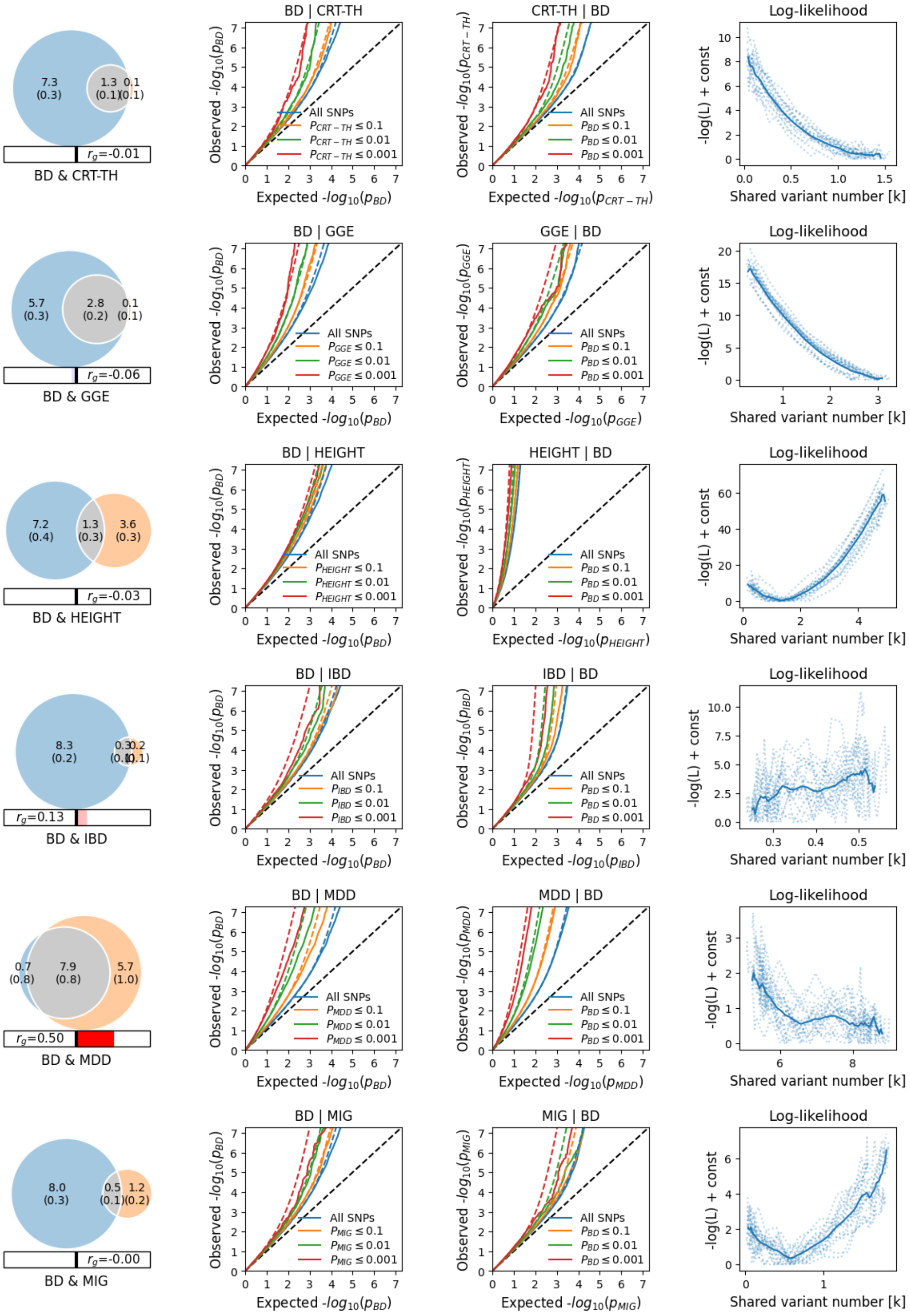
BD | CRT-TH
CRT-TH | BD
Log-likelihood
7.3 (0.3)
1.3 (0.1)
0.1 (0.1)
$r_g$=-0.01
BD & CRT-TH
All SNPs
$P_{CRT-TH} \leq 0.1$
$P_{CRT-TH} \leq 0.01$
$P_{CRT-TH} \leq 0.001$
$P_{BD} \leq 0.1$
$P_{BD} \leq 0.01$
$P_{BD} \leq 0.001$
Observed $-log_{10}(p_{BD})$
Expected $-log_{10}(p_{BD})$
Observed $-log_{10}(p_{CRT-TH})$
Expected $-log_{10}(p_{CRT-TH})$
-log(L) + const
Shared variant number [k]
BD | GGE
GGE | BD
5.7 (0.3)
2.8 (0.2)
0.1 (0.1)
$r_g$=-0.06
BD & GGE
$P_{GGE} \leq 0.1$
$P_{GGE} \leq 0.01$
$P_{GGE} \leq 0.001$
Observed $-log_{10}(p_{GGE})$
Expected $-log_{10}(p_{GGE})$
BD | HEIGHT
HEIGHT | BD
7.2 (0.4)
1.3 (0.3)
3.6 (0.3)
$r_g$=-0.03
BD & HEIGHT
$P_{HEIGHT} \leq 0.1$
$P_{HEIGHT} \leq 0.01$
$P_{HEIGHT} \leq 0.001$
Observed $-log_{10}(p_{HEIGHT})$
Expected $-log_{10}(p_{HEIGHT})$
BD | IBD
IBD | BD
8.3 (0.2)
0.3 (0.1)
0.2 (0.1)
$r_g$=0.13
BD & IBD
$P_{IBD} \leq 0.1$
$P_{IBD} \leq 0.01$
$P_{IBD} \leq 0.001$
Observed $-log_{10}(p_{IBD})$
Expected $-log_{10}(p_{IBD})$
BD | MDD
MDD | BD
0.7 (0.8)
7.9 (0.8)
5.7 (1.0)
$r_g$=0.50
BD & MDD
$P_{MDD} \leq 0.1$
$P_{MDD} \leq 0.01$
$P_{MDD} \leq 0.001$
Observed $-log_{10}(p_{MDD})$
Expected $-log_{10}(p_{MDD})$
BD | MIG
MIG | BD
8.0 (0.3)
0.5 (0.1)
1.2 (0.2)
$r_g$=-0.00
BD & MIG
$P_{MIG} \leq 0.1$
$P_{MIG} \leq 0.01$
$P_{MIG} \leq 0.001$
Observed $-log_{10}(p_{MIG})$
Expected $-log_{10}(p_{MIG})$

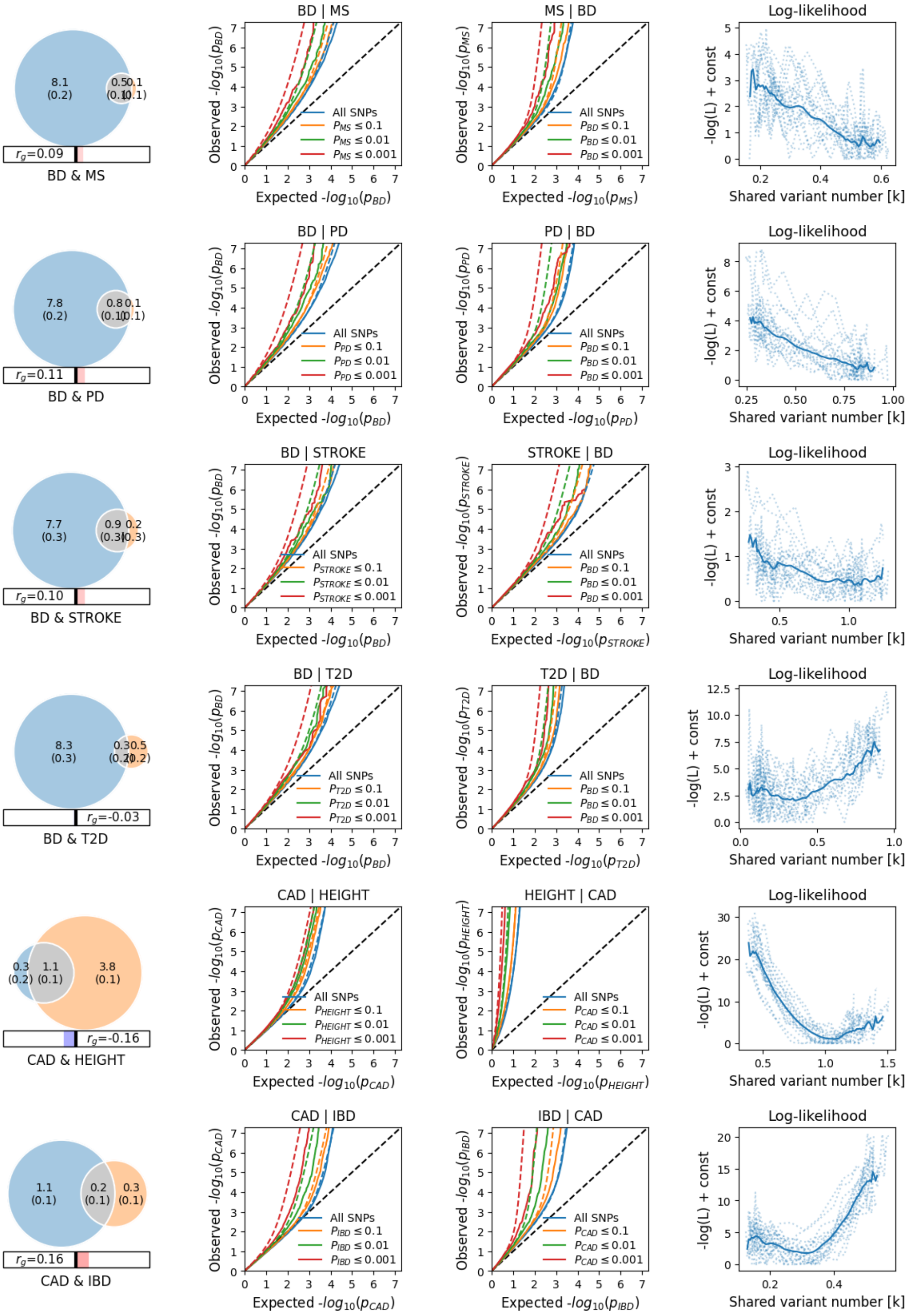

8.1
(0.2)
0.5
(0.1)
0.1
(0.1)
$r_g$=0.09
BD & MS
BD | MS
Observed -$log_{10}(p_{BD})$
Expected -$log_{10}(p_{BD})$
All SNPs
$P_{MS} \leq 0.1$
$P_{MS} \leq 0.01$
$P_{MS} \leq 0.001$
MS | BD
Observed -$log_{10}(p_{MS})$
Expected -$log_{10}(p_{MS})$
$P_{BD} \leq 0.1$
$P_{BD} \leq 0.01$
$P_{BD} \leq 0.001$
Log-likelihood
-log(L) + const
Shared variant number [k]
7.8
(0.2)
0.8
(0.1)
0.1
(0.1)
$r_g$=0.11
BD & PD
BD | PD
$P_{PD} \leq 0.1$
$P_{PD} \leq 0.01$
$P_{PD} \leq 0.001$
PD | BD
Observed -$log_{10}(p_{PD})$
Expected -$log_{10}(p_{PD})$
7.7
(0.3)
0.9
(0.3)
0.2
(0.3)
$r_g$=0.10
BD & STROKE
BD | STROKE
$P_{STROKE} \leq 0.1$
$P_{STROKE} \leq 0.01$
$P_{STROKE} \leq 0.001$
STROKE | BD
Observed -$log_{10}(p_{STROKE})$
Expected -$log_{10}(p_{STROKE})$
8.3
(0.3)
0.3
(0.2)
0.5
(0.2)
$r_g$=-0.03
BD & T2D
BD | T2D
$P_{T2D} \leq 0.1$
$P_{T2D} \leq 0.01$
$P_{T2D} \leq 0.001$
T2D | BD
Observed -$log_{10}(p_{T2D})$
Expected -$log_{10}(p_{T2D})$
0.3
(0.2)
1.1
(0.1)
3.8
(0.1)
$r_g$=-0.16
CAD & HEIGHT
CAD | HEIGHT
Observed -$log_{10}(p_{CAD})$
Expected -$log_{10}(p_{CAD})$
$P_{HEIGHT} \leq 0.1$
$P_{HEIGHT} \leq 0.01$
$P_{HEIGHT} \leq 0.001$
HEIGHT | CAD
Observed -$log_{10}(p_{HEIGHT})$
Expected -$log_{10}(p_{HEIGHT})$
$P_{CAD} \leq 0.1$
$P_{CAD} \leq 0.01$
$P_{CAD} \leq 0.001$
1.1
(0.1)
0.2
(0.1)
0.3
(0.1)
$r_g$=0.16
CAD & IBD
CAD | IBD
$P_{IBD} \leq 0.1$
$P_{IBD} \leq 0.01$
$P_{IBD} \leq 0.001$
IBD | CAD
Observed -$log_{10}(p_{IBD})$
Expected -$log_{10}(p_{IBD})$

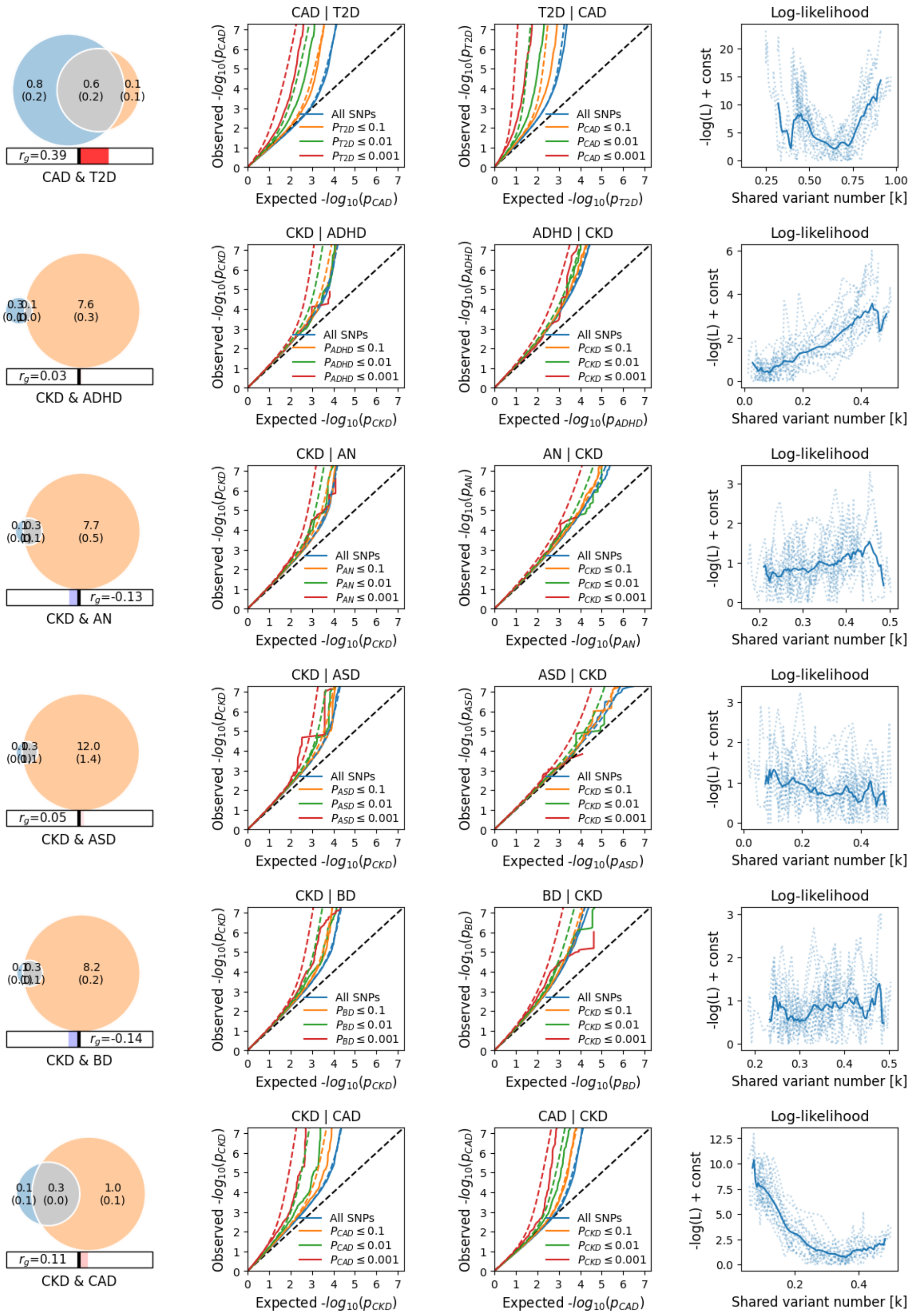

0.8 (0.2)
0.6 (0.2)
0.1 (0.1)
$r_g$=0.39
CAD & T2D
CAD | T2D
Observed -$log_{10}(p_{CAD})$
Expected -$log_{10}(p_{CAD})$
All SNPs
$P_{T2D} \leq 0.1$
$P_{T2D} \leq 0.01$
$P_{T2D} \leq 0.001$
T2D | CAD
Observed -$log_{10}(p_{T2D})$
Expected -$log_{10}(p_{T2D})$
$P_{CAD} \leq 0.1$
$P_{CAD} \leq 0.01$
$P_{CAD} \leq 0.001$
Log-likelihood
-log(L) + const
Shared variant number [k]
7.6 (0.3)
$r_g$=0.03
CKD & ADHD
CKD | ADHD
ADHD | CKD
7.7 (0.5)
$r_g$=-0.13
CKD & AN
CKD | AN
AN | CKD
12.0 (1.4)
$r_g$=0.05
CKD & ASD
CKD | ASD
ASD | CKD
8.2 (0.2)
$r_g$=-0.14
CKD & BD
CKD | BD
BD | CKD
0.1 (0.1)
0.3 (0.0)
1.0 (0.1)
$r_g$=0.11
CKD & CAD
CKD | CAD
CAD | CKD

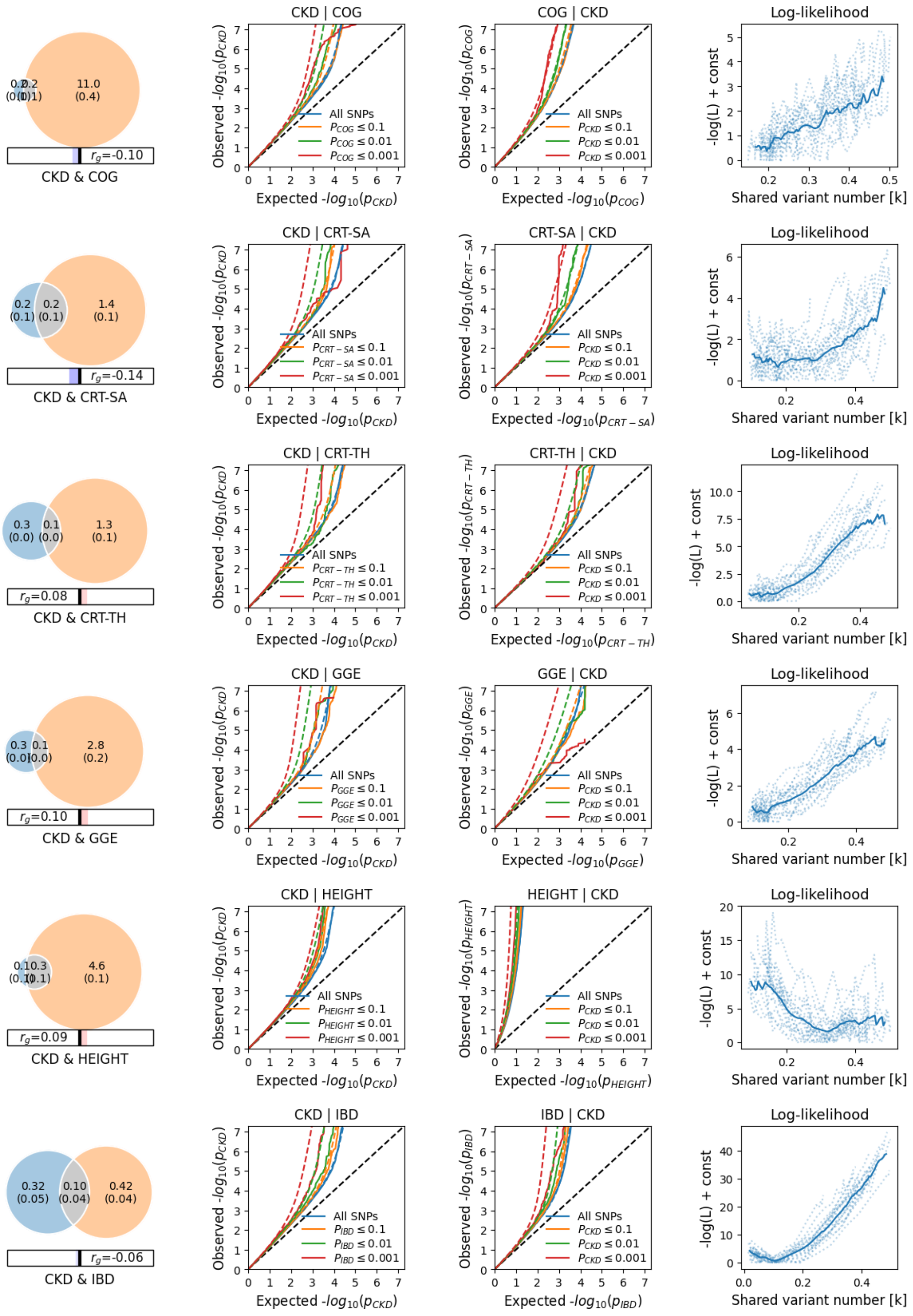

CKD & COG
$r_g$=-0.10
CKD | COG
COG | CKD
Log-likelihood
CKD & CRT-SA
$r_g$=-0.14
CKD | CRT-SA
CRT-SA | CKD
CKD & CRT-TH
$r_g$=0.08
CKD | CRT-TH
CRT-TH | CKD
CKD & GGE
$r_g$=0.10
CKD | GGE
GGE | CKD
CKD & HEIGHT
$r_g$=0.09
CKD | HEIGHT
HEIGHT | CKD
CKD & IBD
$r_g$=-0.06
CKD | IBD
IBD | CKD
All SNPs
Observed -$log_{10}(p_{CKD})$
Expected -$log_{10}(p_{CKD})$
-log(L) + const
Shared variant number [k]

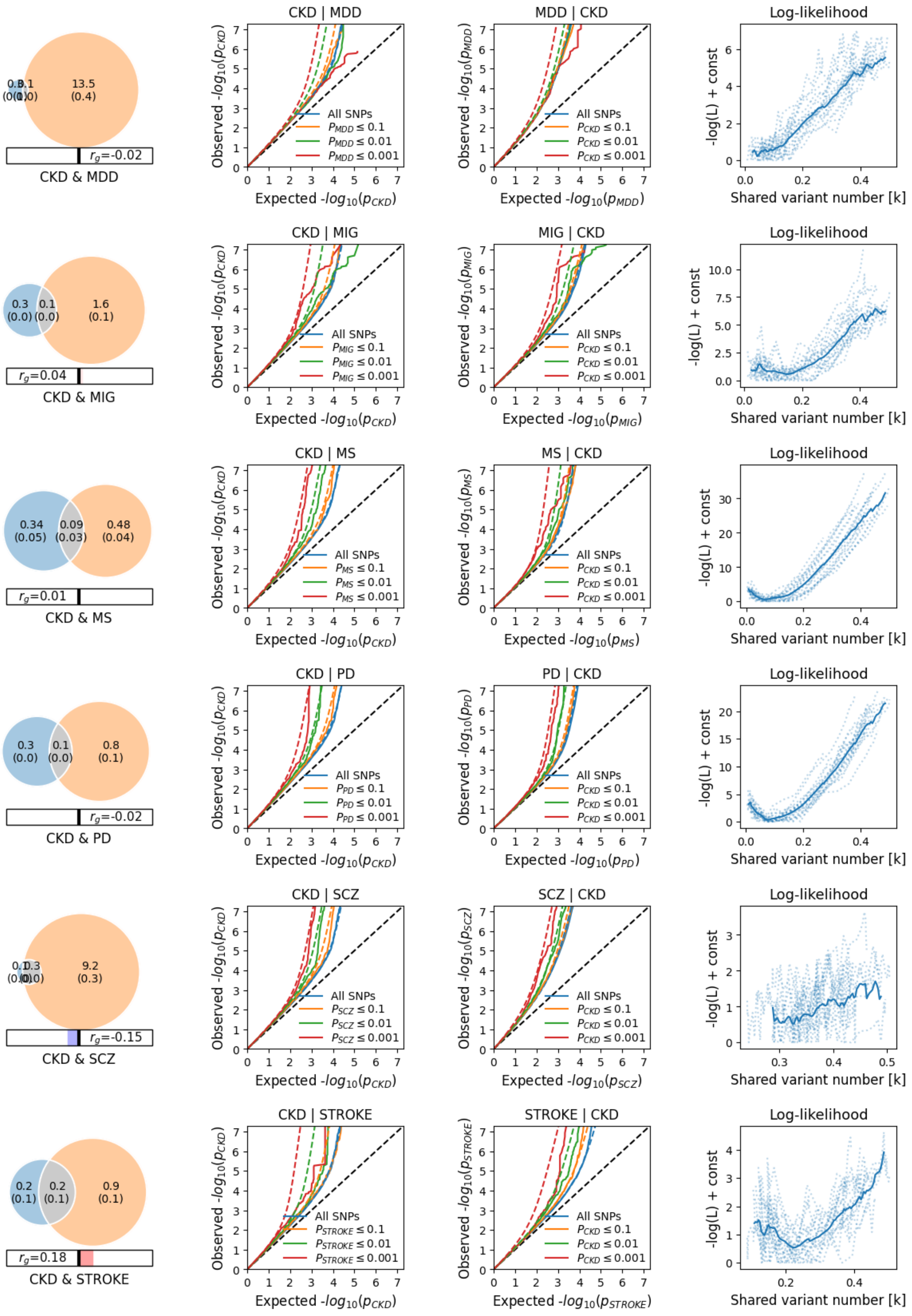

CKD & MDD
$r_g$=-0.02
13.5
(0.4)
CKD | MDD
MDD | CKD
Log-likelihood
All SNPs
$P_{MDD} \leq 0.1$
$P_{MDD} \leq 0.01$
$P_{MDD} \leq 0.001$
$P_{CKD} \leq 0.1$
$P_{CKD} \leq 0.01$
$P_{CKD} \leq 0.001$
Observed -$log_{10}(p_{CKD})$
Expected -$log_{10}(p_{CKD})$
Observed -$log_{10}(p_{MDD})$
Expected -$log_{10}(p_{MDD})$
-log(L) + const
Shared variant number [k]
CKD & MIG
0.3 (0.0) 0.1 (0.0) 1.6 (0.1)
$r_g$=0.04
CKD | MIG
MIG | CKD
$P_{MIG} \leq 0.1$
$P_{MIG} \leq 0.01$
$P_{MIG} \leq 0.001$
Observed -$log_{10}(p_{MIG})$
Expected -$log_{10}(p_{MIG})$
CKD & MS
0.34 (0.05) 0.09 (0.03) 0.48 (0.04)
$r_g$=0.01
CKD | MS
MS | CKD
$P_{MS} \leq 0.1$
$P_{MS} \leq 0.01$
$P_{MS} \leq 0.001$
Observed -$log_{10}(p_{MS})$
Expected -$log_{10}(p_{MS})$
CKD & PD
0.3 (0.0) 0.1 (0.0) 0.8 (0.1)
$r_g$=-0.02
CKD | PD
PD | CKD
$P_{PD} \leq 0.1$
$P_{PD} \leq 0.01$
$P_{PD} \leq 0.001$
Observed -$log_{10}(p_{PD})$
Expected -$log_{10}(p_{PD})$
CKD & SCZ
9.2 (0.3)
$r_g$=-0.15
CKD | SCZ
SCZ | CKD
$P_{SCZ} \leq 0.1$
$P_{SCZ} \leq 0.01$
$P_{SCZ} \leq 0.001$
Observed -$log_{10}(p_{SCZ})$
Expected -$log_{10}(p_{SCZ})$
CKD & STROKE
0.2 (0.1) 0.2 (0.1) 0.9 (0.1)
$r_g$=0.18
CKD | STROKE
STROKE | CKD
$P_{STROKE} \leq 0.1$
$P_{STROKE} \leq 0.01$
$P_{STROKE} \leq 0.001$
Observed -$log_{10}(p_{STROKE})$
Expected -$log_{10}(p_{STROKE})$

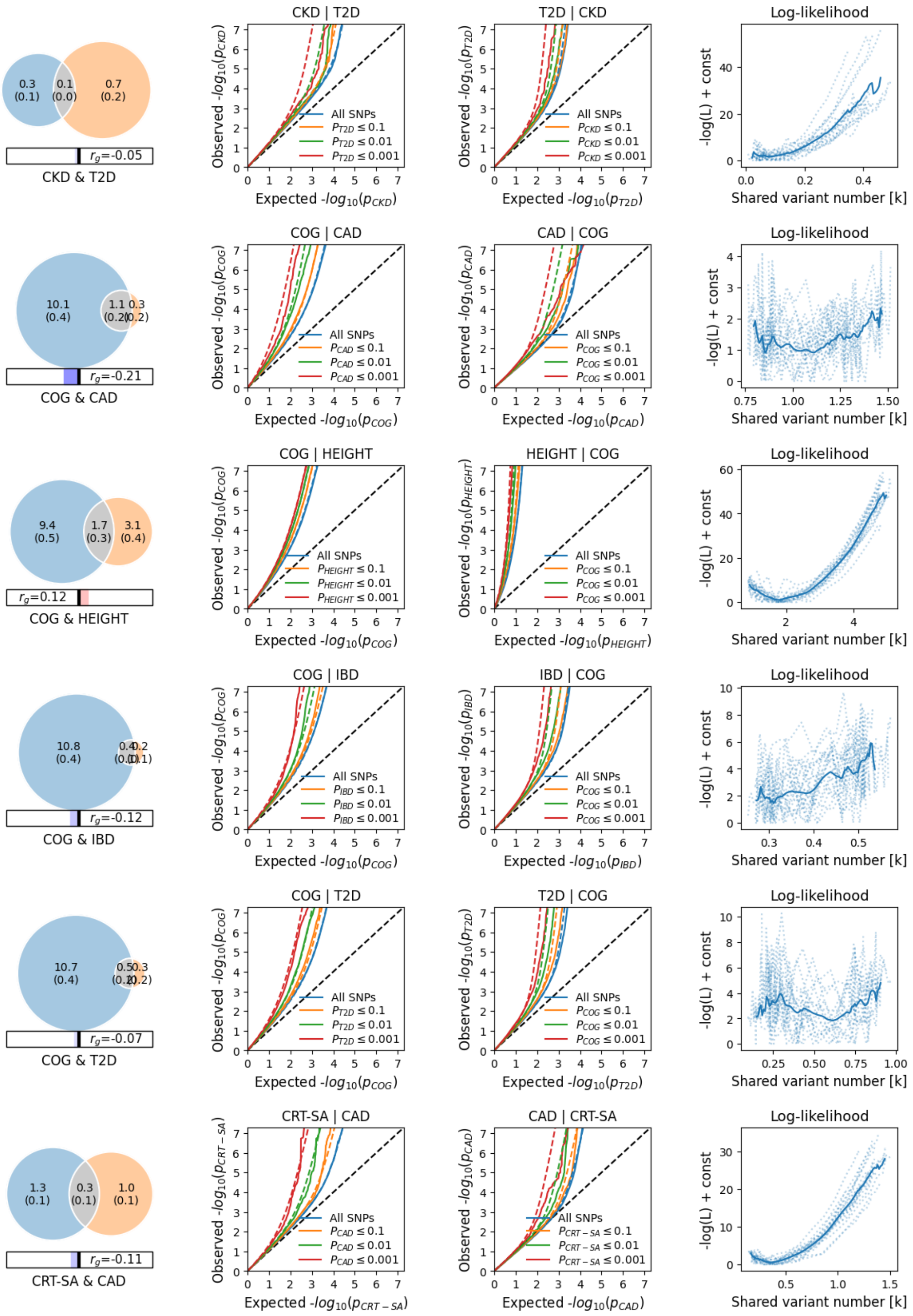
CKD | T2D
T2D | CKD
Log-likelihood
0.3 (0.1)
0.1 (0.0)
0.7 (0.2)
$r_g$=-0.05
CKD & T2D
Observed -$log_{10}(p_{CKD})$
Expected -$log_{10}(p_{CKD})$
Observed -$log_{10}(p_{T2D})$
Expected -$log_{10}(p_{T2D})$
All SNPs
$P_{T2D} \leq 0.1$
$P_{T2D} \leq 0.01$
$P_{T2D} \leq 0.001$
$P_{CKD} \leq 0.1$
$P_{CKD} \leq 0.01$
$P_{CKD} \leq 0.001$
-log(L) + const
Shared variant number [k]
COG | CAD
CAD | COG
10.1 (0.4)
1.1 (0.2)
0.3 (0.2)
$r_g$=-0.21
COG & CAD
Observed -$log_{10}(p_{COG})$
Expected -$log_{10}(p_{COG})$
Observed -$log_{10}(p_{CAD})$
Expected -$log_{10}(p_{CAD})$
$P_{CAD} \leq 0.1$
$P_{CAD} \leq 0.01$
$P_{CAD} \leq 0.001$
$P_{COG} \leq 0.1$
$P_{COG} \leq 0.01$
$P_{COG} \leq 0.001$
COG | HEIGHT
HEIGHT | COG
9.4 (0.5)
1.7 (0.3)
3.1 (0.4)
$r_g$=0.12
COG & HEIGHT
Observed -$log_{10}(p_{HEIGHT})$
Expected -$log_{10}(p_{HEIGHT})$
$P_{HEIGHT} \leq 0.1$
$P_{HEIGHT} \leq 0.01$
$P_{HEIGHT} \leq 0.001$
COG | IBD
IBD | COG
10.8 (0.4)
0.4 (0.0)
0.2 (0.1)
$r_g$=-0.12
COG & IBD
Observed -$log_{10}(p_{IBD})$
Expected -$log_{10}(p_{IBD})$
$P_{IBD} \leq 0.1$
$P_{IBD} \leq 0.01$
$P_{IBD} \leq 0.001$
COG | T2D
T2D | COG
10.7 (0.4)
0.5 (0.2)
0.3 (0.2)
$r_g$=-0.07
COG & T2D
CRT-SA | CAD
CAD | CRT-SA
1.3 (0.1)
0.3 (0.1)
1.0 (0.1)
$r_g$=-0.11
CRT-SA & CAD
Observed -$log_{10}(p_{CRT-SA})$
Expected -$log_{10}(p_{CRT-SA})$
$P_{CRT-SA} \leq 0.1$
$P_{CRT-SA} \leq 0.01$
$P_{CRT-SA} \leq 0.001$

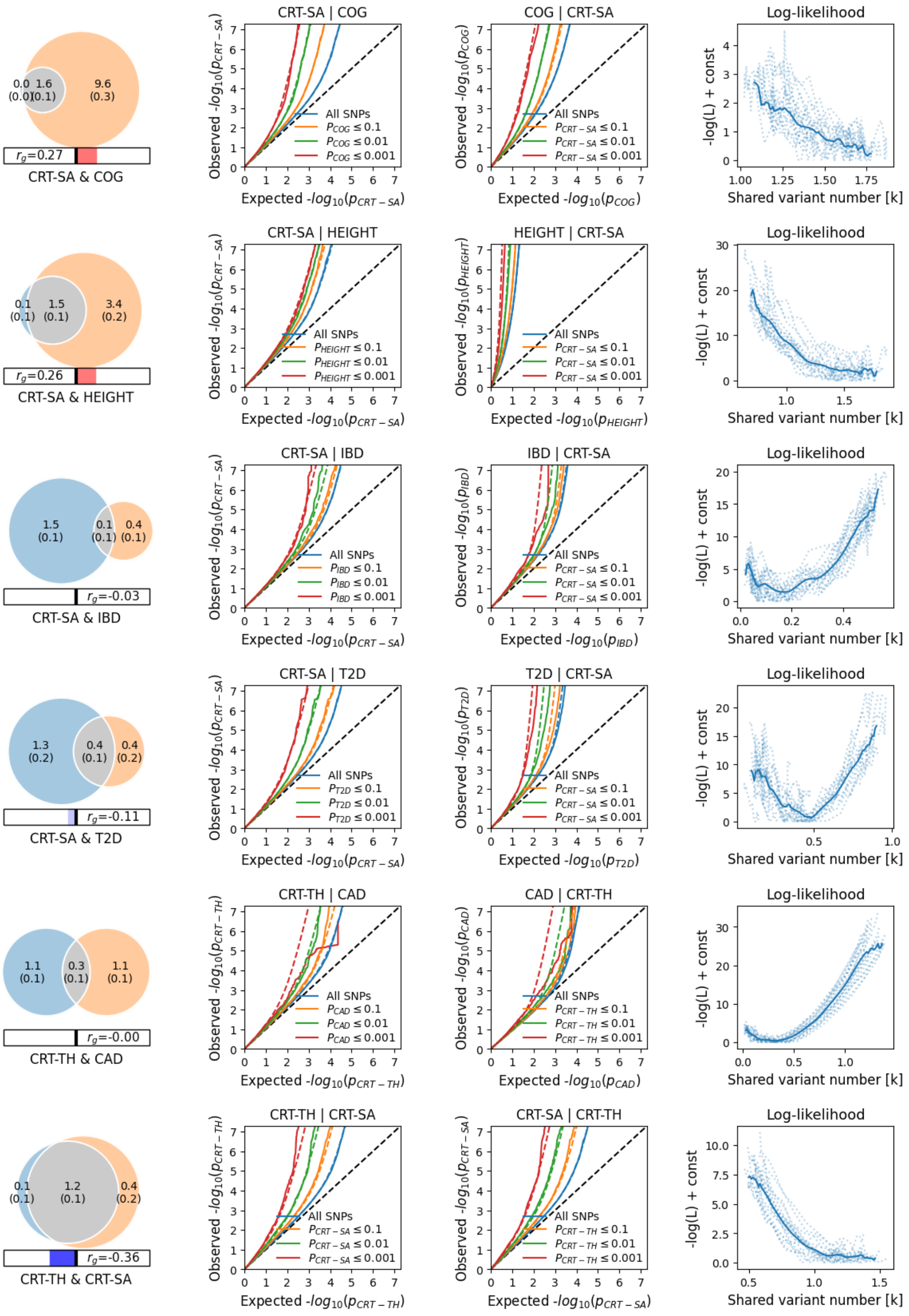
CRT-SA & COG
$r_g$=0.27
CRT-SA | COG
COG | CRT-SA
Log-likelihood
CRT-SA & HEIGHT
$r_g$=0.26
CRT-SA | HEIGHT
HEIGHT | CRT-SA
Log-likelihood
CRT-SA & IBD
$r_g$=-0.03
CRT-SA | IBD
IBD | CRT-SA
Log-likelihood
CRT-SA & T2D
$r_g$=-0.11
CRT-SA | T2D
T2D | CRT-SA
Log-likelihood
CRT-TH & CAD
$r_g$=-0.00
CRT-TH | CAD
CAD | CRT-TH
Log-likelihood
CRT-TH & CRT-SA
$r_g$=-0.36
CRT-TH | CRT-SA
CRT-SA | CRT-TH
Log-likelihood
All SNPs
Observed -$log_{10}(p)$
Expected -$log_{10}(p)$
-log(L) + const
Shared variant number [k]

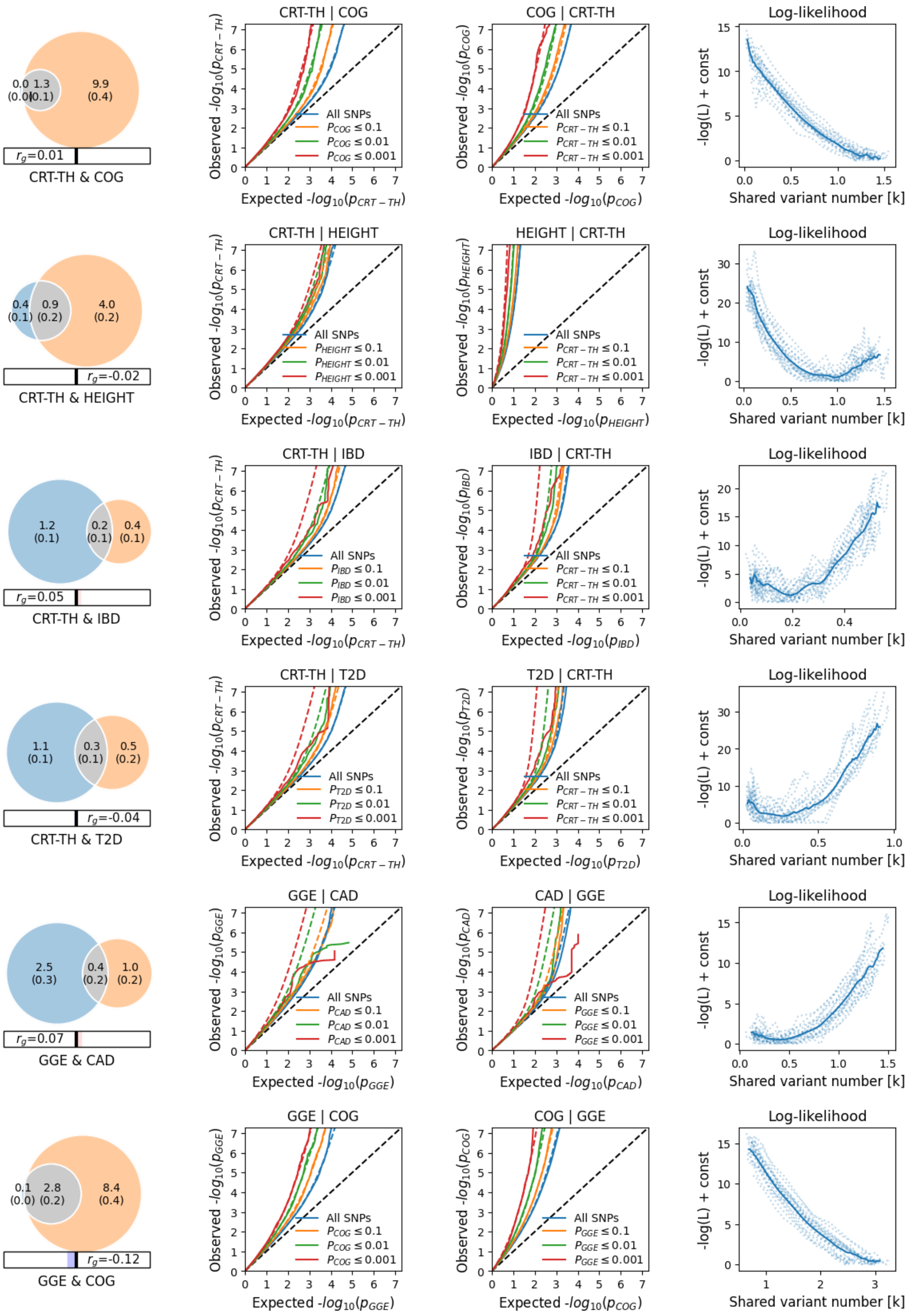

0.0 (0.0)
1.3 (0.1)
9.9 (0.4)
$r_g$=0.01
CRT-TH & COG
CRT-TH | COG
COG | CRT-TH
Log-likelihood
0.4 (0.1)
0.9 (0.2)
4.0 (0.2)
$r_g$=-0.02
CRT-TH & HEIGHT
CRT-TH | HEIGHT
HEIGHT | CRT-TH
Log-likelihood
1.2 (0.1)
0.2 (0.1)
0.4 (0.1)
$r_g$=0.05
CRT-TH & IBD
CRT-TH | IBD
IBD | CRT-TH
Log-likelihood
1.1 (0.1)
0.3 (0.1)
0.5 (0.2)
$r_g$=-0.04
CRT-TH & T2D
CRT-TH | T2D
T2D | CRT-TH
Log-likelihood
2.5 (0.3)
0.4 (0.2)
1.0 (0.2)
$r_g$=0.07
GGE & CAD
GGE | CAD
CAD | GGE
Log-likelihood
0.1 (0.0)
2.8 (0.2)
8.4 (0.4)
$r_g$=-0.12
GGE & COG
GGE | COG
COG | GGE
Log-likelihood
All SNPs
Observed -$log_{10}(p)$
Expected -$log_{10}(p)$
-log(L) + const
Shared variant number [k]

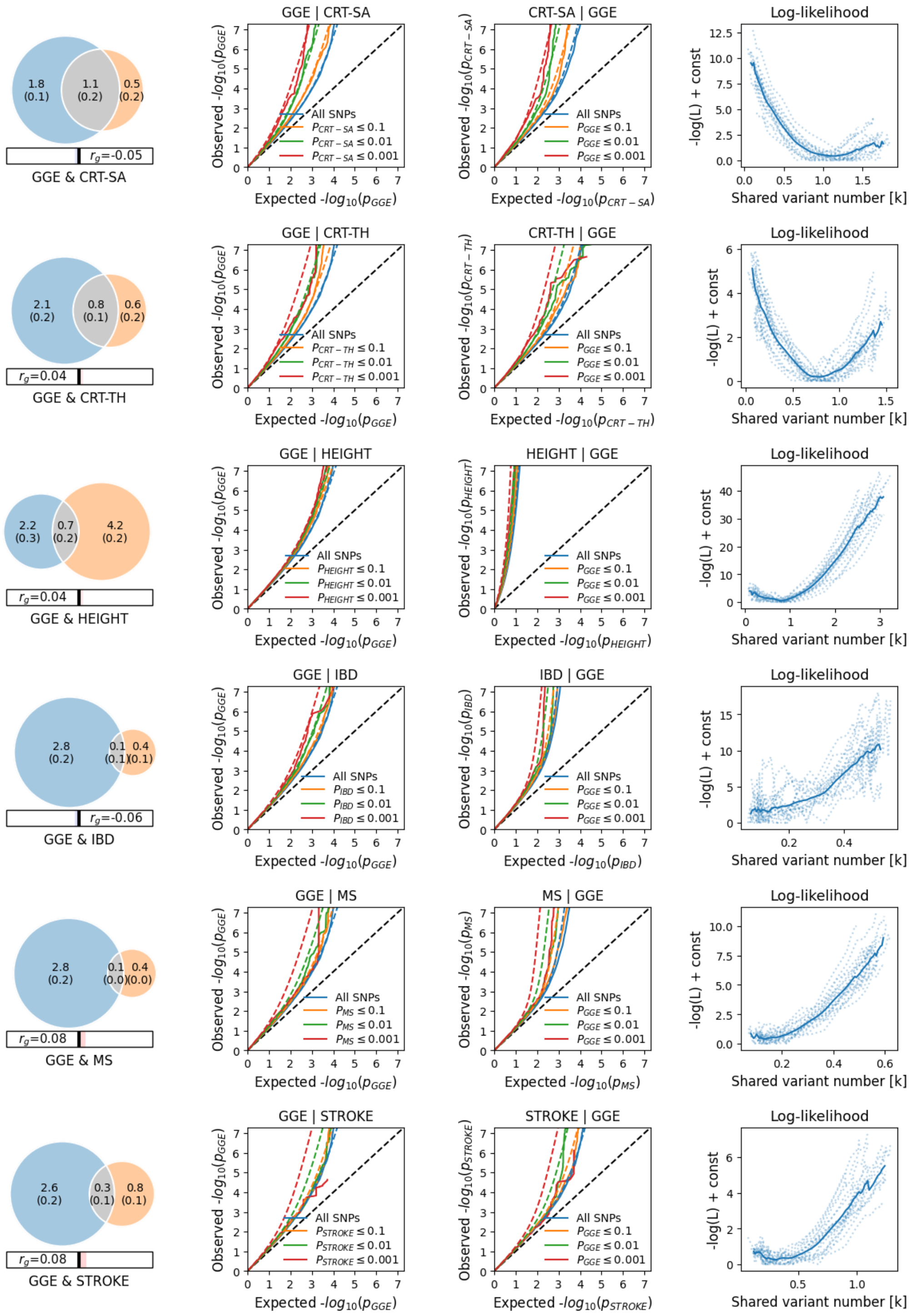

1.8 (0.1)
1.1 (0.2)
0.5 (0.2)
$r_g$=-0.05
GGE & CRT-SA
GGE | CRT-SA
CRT-SA | GGE
Log-likelihood
All SNPs
$P_{CRT-SA} \leq 0.1$
$P_{CRT-SA} \leq 0.01$
$P_{CRT-SA} \leq 0.001$
$P_{GGE} \leq 0.1$
$P_{GGE} \leq 0.01$
$P_{GGE} \leq 0.001$
Observed -$log_{10}(p_{GGE})$
Expected -$log_{10}(p_{GGE})$
Observed -$log_{10}(p_{CRT-SA})$
Expected -$log_{10}(p_{CRT-SA})$
-log(L) + const
Shared variant number [k]
2.1 (0.2)
0.8 (0.1)
0.6 (0.2)
$r_g$=0.04
GGE & CRT-TH
GGE | CRT-TH
CRT-TH | GGE
$P_{CRT-TH} \leq 0.1$
$P_{CRT-TH} \leq 0.01$
$P_{CRT-TH} \leq 0.001$
Observed -$log_{10}(p_{CRT-TH})$
Expected -$log_{10}(p_{CRT-TH})$
2.2 (0.3)
0.7 (0.2)
4.2 (0.2)
$r_g$=0.04
GGE & HEIGHT
GGE | HEIGHT
HEIGHT | GGE
$P_{HEIGHT} \leq 0.1$
$P_{HEIGHT} \leq 0.01$
$P_{HEIGHT} \leq 0.001$
Observed -$log_{10}(p_{HEIGHT})$
Expected -$log_{10}(p_{HEIGHT})$
2.8 (0.2)
0.1 (0.1)
0.4 (0.1)
$r_g$=-0.06
GGE & IBD
GGE | IBD
IBD | GGE
$P_{IBD} \leq 0.1$
$P_{IBD} \leq 0.01$
$P_{IBD} \leq 0.001$
Observed -$log_{10}(p_{IBD})$
Expected -$log_{10}(p_{IBD})$
2.8 (0.2)
0.1 (0.0)
0.4 (0.0)
$r_g$=0.08
GGE & MS
GGE | MS
MS | GGE
$P_{MS} \leq 0.1$
$P_{MS} \leq 0.01$
$P_{MS} \leq 0.001$
Observed -$log_{10}(p_{MS})$
Expected -$log_{10}(p_{MS})$
2.6 (0.2)
0.3 (0.1)
0.8 (0.1)
$r_g$=0.08
GGE & STROKE
GGE | STROKE
STROKE | GGE
$P_{STROKE} \leq 0.1$
$P_{STROKE} \leq 0.01$
$P_{STROKE} \leq 0.001$
Observed -$log_{10}(p_{STROKE})$
Expected -$log_{10}(p_{STROKE})$

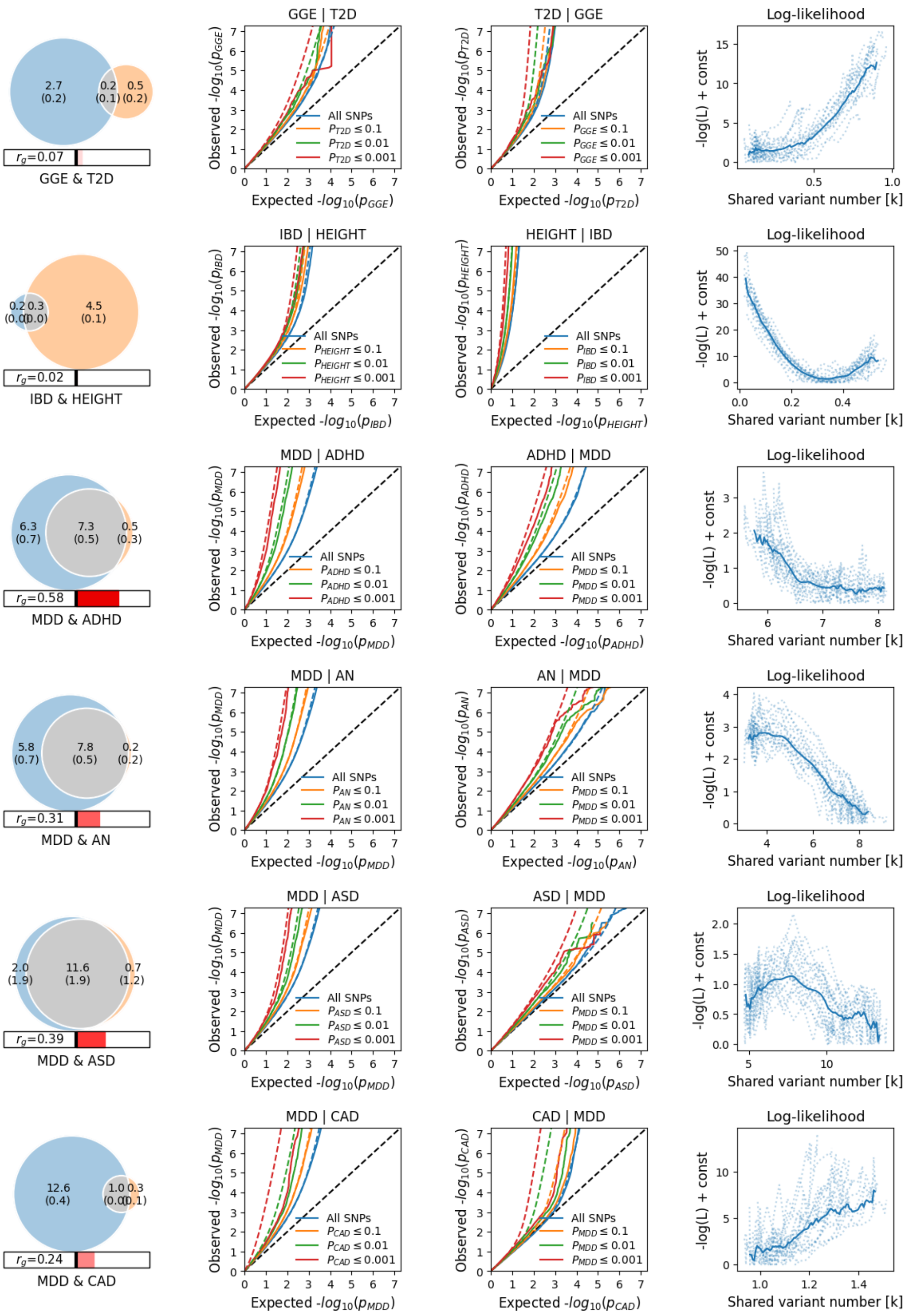

2.7 (0.2) 0.2 (0.1) 0.5 (0.2)
$r_g$=0.07
GGE & T2D
GGE | T2D
Observed -log10(p_GGE)
Expected -log10(p_GGE)
All SNPs
P_T2D ≤ 0.1
P_T2D ≤ 0.01
P_T2D ≤ 0.001
T2D | GGE
Observed -log10(p_T2D)
Expected -log10(p_T2D)
All SNPs
P_GGE ≤ 0.1
P_GGE ≤ 0.01
P_GGE ≤ 0.001
Log-likelihood
-log(L) + const
Shared variant number [k]
0.2 0.3 (0.0) (0.0) 4.5 (0.1)
$r_g$=0.02
IBD & HEIGHT
IBD | HEIGHT
Observed -log10(p_IBD)
Expected -log10(p_IBD)
All SNPs
P_HEIGHT ≤ 0.1
P_HEIGHT ≤ 0.01
P_HEIGHT ≤ 0.001
HEIGHT | IBD
Observed -log10(p_HEIGHT)
Expected -log10(p_HEIGHT)
All SNPs
P_IBD ≤ 0.1
P_IBD ≤ 0.01
P_IBD ≤ 0.001
Log-likelihood
-log(L) + const
Shared variant number [k]
6.3 (0.7) 7.3 (0.5) 0.5 (0.3)
$r_g$=0.58
MDD & ADHD
MDD | ADHD
Observed -log10(p_MDD)
Expected -log10(p_MDD)
All SNPs
P_ADHD ≤ 0.1
P_ADHD ≤ 0.01
P_ADHD ≤ 0.001
ADHD | MDD
Observed -log10(p_ADHD)
Expected -log10(p_ADHD)
All SNPs
P_MDD ≤ 0.1
P_MDD ≤ 0.01
P_MDD ≤ 0.001
Log-likelihood
-log(L) + const
Shared variant number [k]
5.8 (0.7) 7.8 (0.5) 0.2 (0.2)
$r_g$=0.31
MDD & AN
MDD | AN
Observed -log10(p_MDD)
Expected -log10(p_MDD)
All SNPs
P_AN ≤ 0.1
P_AN ≤ 0.01
P_AN ≤ 0.001
AN | MDD
Observed -log10(p_AN)
Expected -log10(p_AN)
All SNPs
P_MDD ≤ 0.1
P_MDD ≤ 0.01
P_MDD ≤ 0.001
Log-likelihood
-log(L) + const
Shared variant number [k]
2.0 (1.9) 11.6 (1.9) 0.7 (1.2)
$r_g$=0.39
MDD & ASD
MDD | ASD
Observed -log10(p_MDD)
Expected -log10(p_MDD)
All SNPs
P_ASD ≤ 0.1
P_ASD ≤ 0.01
P_ASD ≤ 0.001
ASD | MDD
Observed -log10(p_ASD)
Expected -log10(p_ASD)
All SNPs
P_MDD ≤ 0.1
P_MDD ≤ 0.01
P_MDD ≤ 0.001
Log-likelihood
-log(L) + const
Shared variant number [k]
12.6 (0.4) 1.0 (0.0) 0.3 (0.1)
$r_g$=0.24
MDD & CAD
MDD | CAD
Observed -log10(p_MDD)
Expected -log10(p_MDD)
All SNPs
P_CAD ≤ 0.1
P_CAD ≤ 0.01
P_CAD ≤ 0.001
CAD | MDD
Observed -log10(p_CAD)
Expected -log10(p_CAD)
All SNPs
P_MDD ≤ 0.1
P_MDD ≤ 0.01
P_MDD ≤ 0.001
Log-likelihood
-log(L) + const
Shared variant number [k]

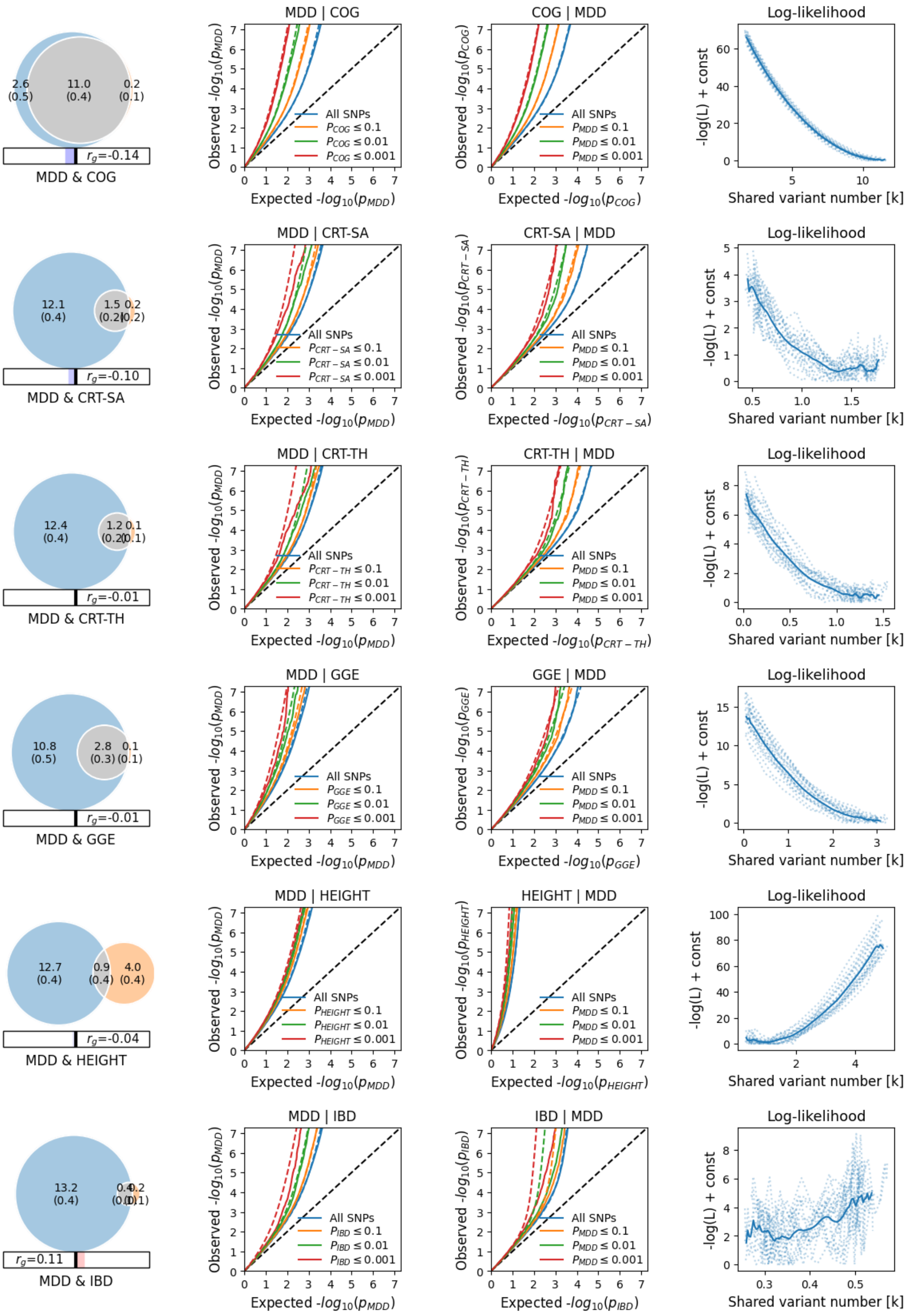

2.6 (0.5)
11.0 (0.4)
0.2 (0.1)
$r_g$=-0.14
MDD & COG
MDD | COG
COG | MDD
Log-likelihood
12.1 (0.4)
1.5 (0.2)
0.2 (0.2)
$r_g$=-0.10
MDD & CRT-SA
MDD | CRT-SA
CRT-SA | MDD
12.4 (0.4)
1.2 (0.2)
0.1 (0.1)
$r_g$=-0.01
MDD & CRT-TH
MDD | CRT-TH
CRT-TH | MDD
10.8 (0.5)
2.8 (0.3)
0.1 (0.1)
$r_g$=-0.01
MDD & GGE
MDD | GGE
GGE | MDD
12.7 (0.4)
0.9 (0.4)
4.0 (0.4)
$r_g$=-0.04
MDD & HEIGHT
MDD | HEIGHT
HEIGHT | MDD
13.2 (0.4)
$r_g$=0.11
MDD & IBD
MDD | IBD
IBD | MDD
Observed -$log_{10}(p)$
Expected -$log_{10}(p)$
All SNPs
$\le$ 0.1
$\le$ 0.01
$\le$ 0.001
-log(L) + const
Shared variant number [k]

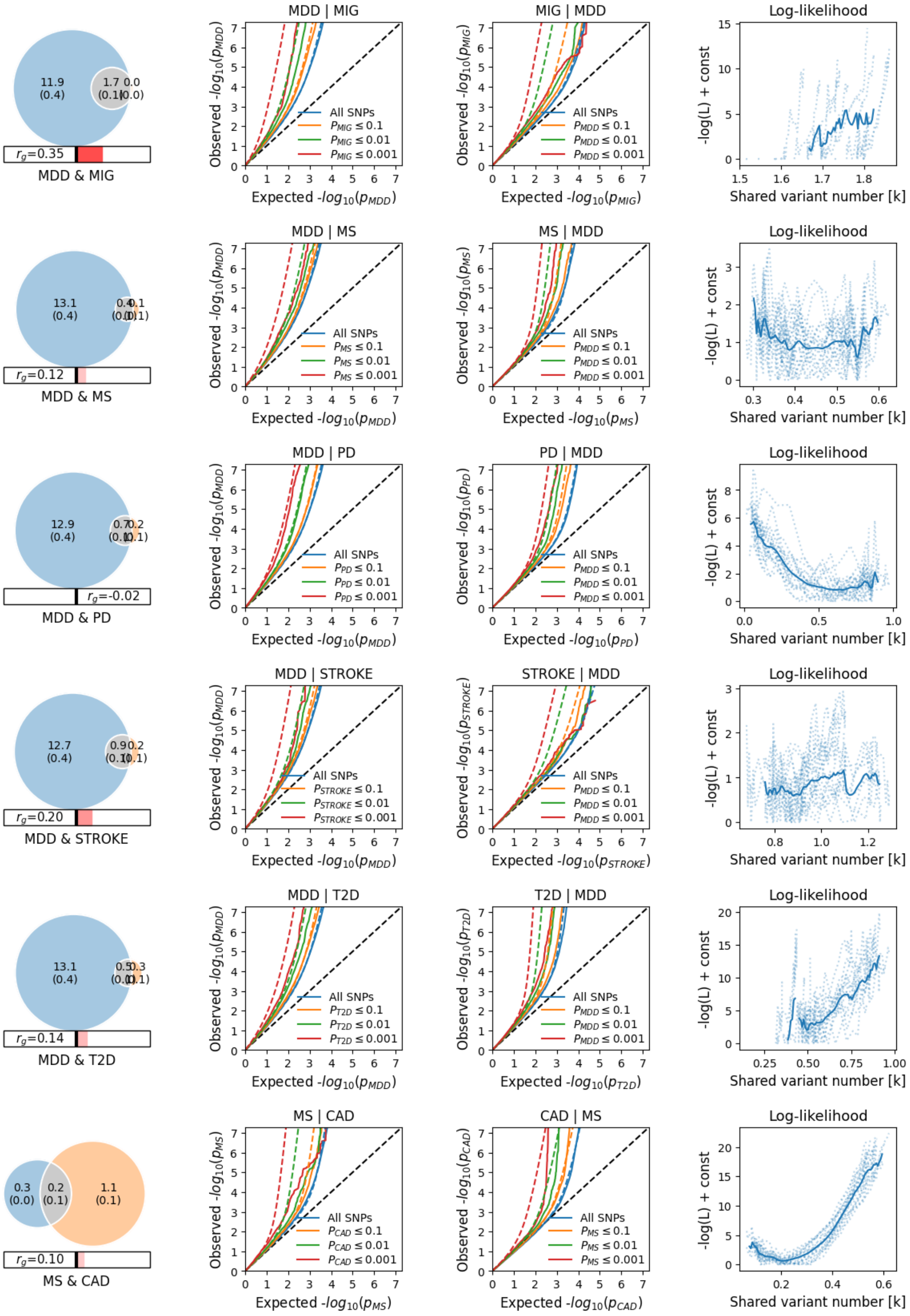
MDD & MIG
$r_g$=0.35
MDD | MIG
MIG | MDD
Log-likelihood
MDD & MS
$r_g$=0.12
MDD | MS
MS | MDD
MDD & PD
$r_g$=-0.02
MDD | PD
PD | MDD
MDD & STROKE
$r_g$=0.20
MDD | STROKE
STROKE | MDD
MDD & T2D
$r_g$=0.14
MDD | T2D
T2D | MDD
MS & CAD
$r_g$=0.10
MS | CAD
CAD | MS
All SNPs
Expected -$log_{10}(p)$
Observed -$log_{10}(p)$
-log(L) + const
Shared variant number [k]

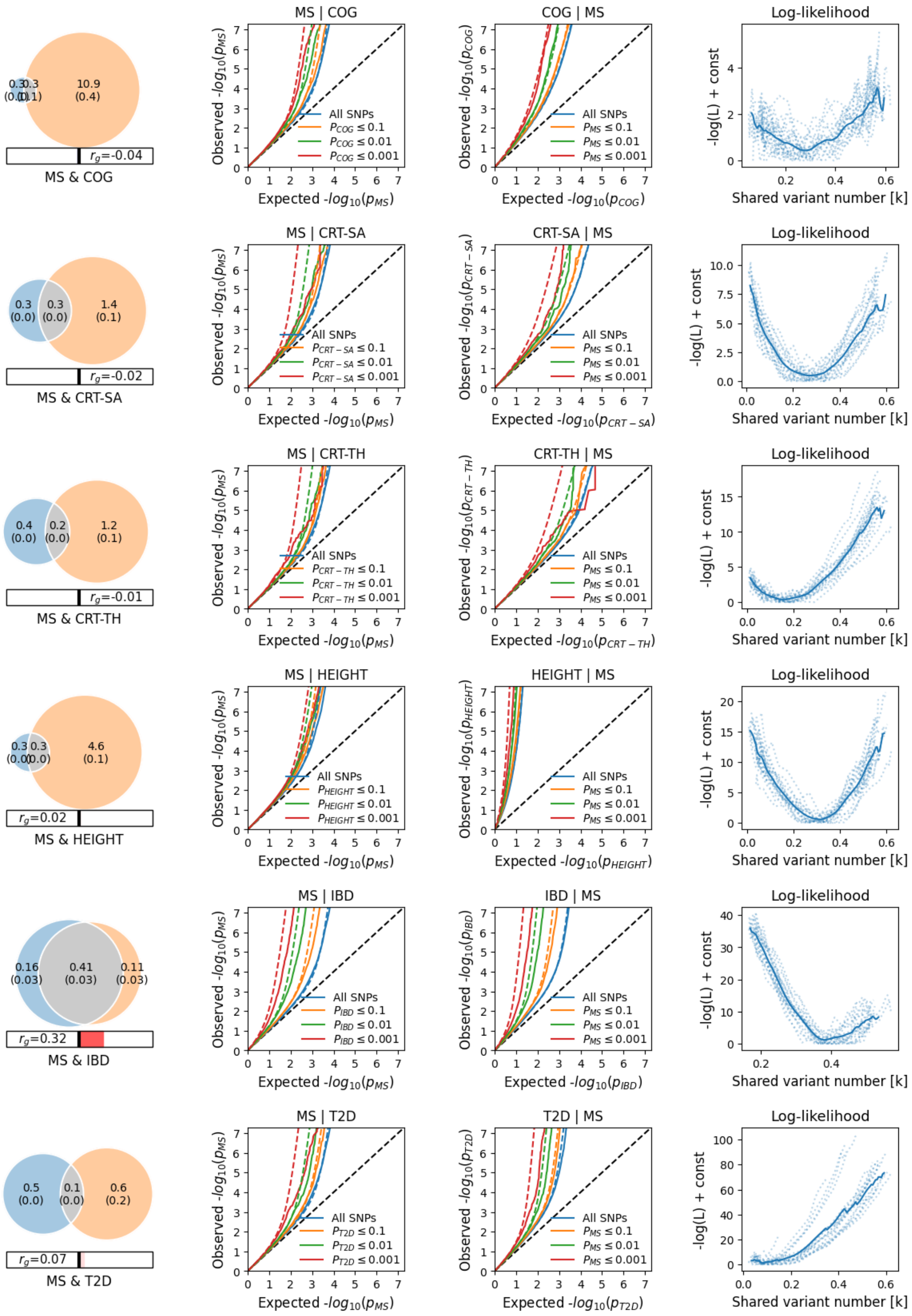

MS & COG
0.3 (0.0)
0.3 (0.1)
10.9 (0.4)
$r_g$=-0.04
MS | COG
COG | MS
Log-likelihood
All SNPs
$P_{COG} \le 0.1$
$P_{COG} \le 0.01$
$P_{COG} \le 0.001$
$P_{MS} \le 0.1$
$P_{MS} \le 0.01$
$P_{MS} \le 0.001$
Observed -$log_{10}(p_{MS})$
Observed -$log_{10}(p_{COG})$
Expected -$log_{10}(p_{MS})$
Expected -$log_{10}(p_{COG})$
-log(L) + const
Shared variant number [k]
MS & CRT-SA
0.3 (0.0)
0.3 (0.0)
1.4 (0.1)
$r_g$=-0.02
MS | CRT-SA
CRT-SA | MS
$P_{CRT-SA} \le 0.1$
$P_{CRT-SA} \le 0.01$
$P_{CRT-SA} \le 0.001$
Observed -$log_{10}(p_{CRT-SA})$
Expected -$log_{10}(p_{CRT-SA})$
MS & CRT-TH
0.4 (0.0)
0.2 (0.0)
1.2 (0.1)
$r_g$=-0.01
MS | CRT-TH
CRT-TH | MS
$P_{CRT-TH} \le 0.1$
$P_{CRT-TH} \le 0.01$
$P_{CRT-TH} \le 0.001$
Observed -$log_{10}(p_{CRT-TH})$
Expected -$log_{10}(p_{CRT-TH})$
MS & HEIGHT
0.3 (0.0)
0.3 (0.0)
4.6 (0.1)
$r_g$=0.02
MS | HEIGHT
HEIGHT | MS
$P_{HEIGHT} \le 0.1$
$P_{HEIGHT} \le 0.01$
$P_{HEIGHT} \le 0.001$
Observed -$log_{10}(p_{HEIGHT})$
Expected -$log_{10}(p_{HEIGHT})$
MS & IBD
0.16 (0.03)
0.41 (0.03)
0.11 (0.03)
$r_g$=0.32
MS | IBD
IBD | MS
$P_{IBD} \le 0.1$
$P_{IBD} \le 0.01$
$P_{IBD} \le 0.001$
Observed -$log_{10}(p_{IBD})$
Expected -$log_{10}(p_{IBD})$
MS & T2D
0.5 (0.0)
0.1 (0.0)
0.6 (0.2)
$r_g$=0.07
MS | T2D
T2D | MS
$P_{T2D} \le 0.1$
$P_{T2D} \le 0.01$
$P_{T2D} \le 0.001$
Observed -$log_{10}(p_{T2D})$
Expected -$log_{10}(p_{T2D})$

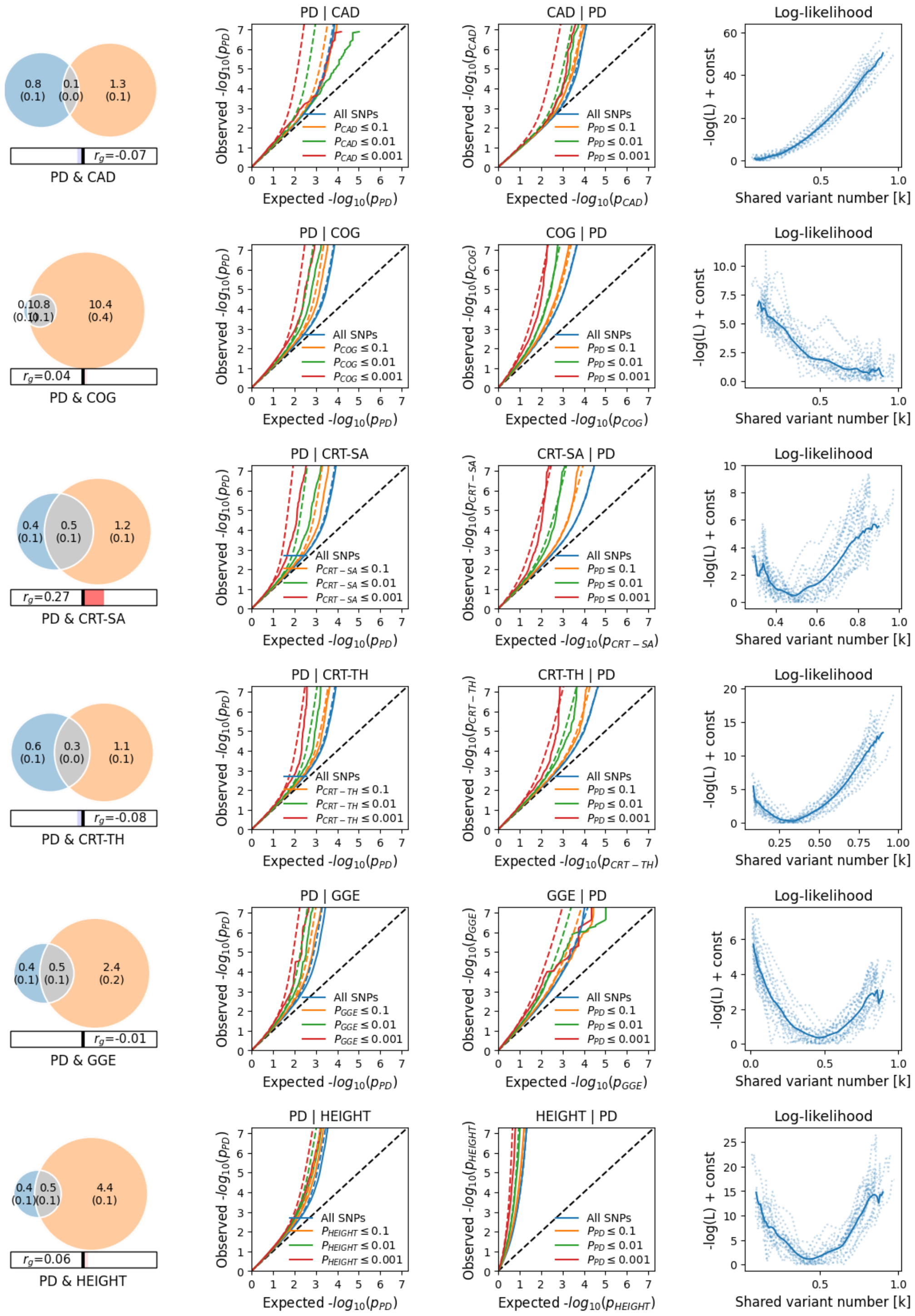
0.8 (0.1)
0.1 (0.0)
1.3 (0.1)
$r_g$=-0.07
PD & CAD
PD | CAD
CAD | PD
Log-likelihood
All SNPs
$p_{CAD} \leq 0.1$
$p_{CAD} \leq 0.01$
$p_{CAD} \leq 0.001$
$p_{PD} \leq 0.1$
$p_{PD} \leq 0.01$
$p_{PD} \leq 0.001$
Observed -$log_{10}(p_{PD})$
Expected -$log_{10}(p_{PD})$
Observed -$log_{10}(p_{CAD})$
Expected -$log_{10}(p_{CAD})$
-log(L) + const
Shared variant number [k]
0.1 0.8 (0.1) (0.1)
10.4 (0.4)
$r_g$=0.04
PD & COG
PD | COG
COG | PD
$p_{COG} \leq 0.1$
$p_{COG} \leq 0.01$
$p_{COG} \leq 0.001$
Observed -$log_{10}(p_{COG})$
Expected -$log_{10}(p_{COG})$
0.4 (0.1)
0.5 (0.1)
1.2 (0.1)
$r_g$=0.27
PD & CRT-SA
PD | CRT-SA
CRT-SA | PD
$p_{CRT-SA} \leq 0.1$
$p_{CRT-SA} \leq 0.01$
$p_{CRT-SA} \leq 0.001$
Observed -$log_{10}(p_{CRT-SA})$
Expected -$log_{10}(p_{CRT-SA})$
0.6 (0.1)
0.3 (0.0)
1.1 (0.1)
$r_g$=-0.08
PD & CRT-TH
PD | CRT-TH
CRT-TH | PD
$p_{CRT-TH} \leq 0.1$
$p_{CRT-TH} \leq 0.01$
$p_{CRT-TH} \leq 0.001$
Observed -$log_{10}(p_{CRT-TH})$
Expected -$log_{10}(p_{CRT-TH})$
0.4 (0.1)
0.5 (0.1)
2.4 (0.2)
$r_g$=-0.01
PD & GGE
PD | GGE
GGE | PD
$p_{GGE} \leq 0.1$
$p_{GGE} \leq 0.01$
$p_{GGE} \leq 0.001$
Observed -$log_{10}(p_{GGE})$
Expected -$log_{10}(p_{GGE})$
0.4 (0.1)
0.5 (0.1)
4.4 (0.1)
$r_g$=0.06
PD & HEIGHT
PD | HEIGHT
HEIGHT | PD
$p_{HEIGHT} \leq 0.1$
$p_{HEIGHT} \leq 0.01$
$p_{HEIGHT} \leq 0.001$
Observed -$log_{10}(p_{HEIGHT})$
Expected -$log_{10}(p_{HEIGHT})$

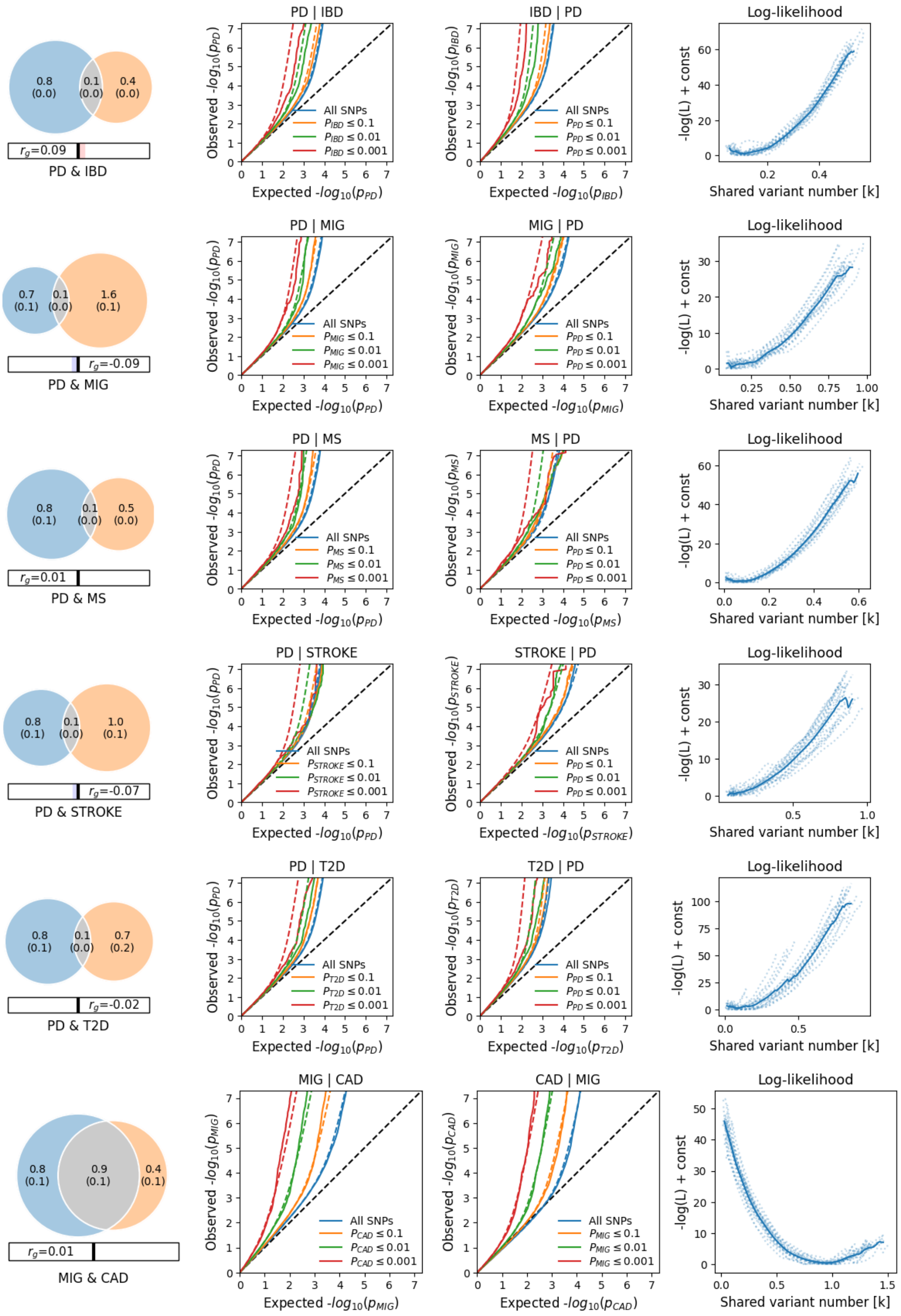
0.8 (0.0)
0.1 (0.0)
0.4 (0.0)
$r_g$=0.09
PD & IBD
PD | IBD
Observed -$log_{10}(p_{PD})$
Expected -$log_{10}(p_{PD})$
All SNPs
$P_{IBD} \leq 0.1$
$P_{IBD} \leq 0.01$
$P_{IBD} \leq 0.001$
IBD | PD
Observed -$log_{10}(p_{IBD})$
Expected -$log_{10}(p_{IBD})$
All SNPs
$P_{PD} \leq 0.1$
$P_{PD} \leq 0.01$
$P_{PD} \leq 0.001$
Log-likelihood
-log(L) + const
Shared variant number [k]
0.7 (0.1)
0.1 (0.0)
1.6 (0.1)
$r_g$=-0.09
PD & MIG
PD | MIG
Observed -$log_{10}(p_{PD})$
Expected -$log_{10}(p_{PD})$
All SNPs
$P_{MIG} \leq 0.1$
$P_{MIG} \leq 0.01$
$P_{MIG} \leq 0.001$
MIG | PD
Observed -$log_{10}(p_{MIG})$
Expected -$log_{10}(p_{MIG})$
All SNPs
$P_{PD} \leq 0.1$
$P_{PD} \leq 0.01$
$P_{PD} \leq 0.001$
Log-likelihood
-log(L) + const
Shared variant number [k]
0.8 (0.1)
0.1 (0.0)
0.5 (0.0)
$r_g$=0.01
PD & MS
PD | MS
Observed -$log_{10}(p_{PD})$
Expected -$log_{10}(p_{PD})$
All SNPs
$P_{MS} \leq 0.1$
$P_{MS} \leq 0.01$
$P_{MS} \leq 0.001$
MS | PD
Observed -$log_{10}(p_{MS})$
Expected -$log_{10}(p_{MS})$
All SNPs
$P_{PD} \leq 0.1$
$P_{PD} \leq 0.01$
$P_{PD} \leq 0.001$
Log-likelihood
-log(L) + const
Shared variant number [k]
0.8 (0.1)
0.1 (0.0)
1.0 (0.1)
$r_g$=-0.07
PD & STROKE
PD | STROKE
Observed -$log_{10}(p_{PD})$
Expected -$log_{10}(p_{PD})$
All SNPs
$P_{STROKE} \leq 0.1$
$P_{STROKE} \leq 0.01$
$P_{STROKE} \leq 0.001$
STROKE | PD
Observed -$log_{10}(p_{STROKE})$
Expected -$log_{10}(p_{STROKE})$
All SNPs
$P_{PD} \leq 0.1$
$P_{PD} \leq 0.01$
$P_{PD} \leq 0.001$
Log-likelihood
-log(L) + const
Shared variant number [k]
0.8 (0.1)
0.1 (0.0)
0.7 (0.2)
$r_g$=-0.02
PD & T2D
PD | T2D
Observed -$log_{10}(p_{PD})$
Expected -$log_{10}(p_{PD})$
All SNPs
$P_{T2D} \leq 0.1$
$P_{T2D} \leq 0.01$
$P_{T2D} \leq 0.001$
T2D | PD
Observed -$log_{10}(p_{T2D})$
Expected -$log_{10}(p_{T2D})$
All SNPs
$P_{PD} \leq 0.1$
$P_{PD} \leq 0.01$
$P_{PD} \leq 0.001$
Log-likelihood
-log(L) + const
Shared variant number [k]
0.8 (0.1)
0.9 (0.1)
0.4 (0.1)
$r_g$=0.01
MIG & CAD
MIG | CAD
Observed -$log_{10}(p_{MIG})$
Expected -$log_{10}(p_{MIG})$
All SNPs
$P_{CAD} \leq 0.1$
$P_{CAD} \leq 0.01$
$P_{CAD} \leq 0.001$
CAD | MIG
Observed -$log_{10}(p_{CAD})$
Expected -$log_{10}(p_{CAD})$
All SNPs
$P_{MIG} \leq 0.1$
$P_{MIG} \leq 0.01$
$P_{MIG} \leq 0.001$
Log-likelihood
-log(L) + const
Shared variant number [k]

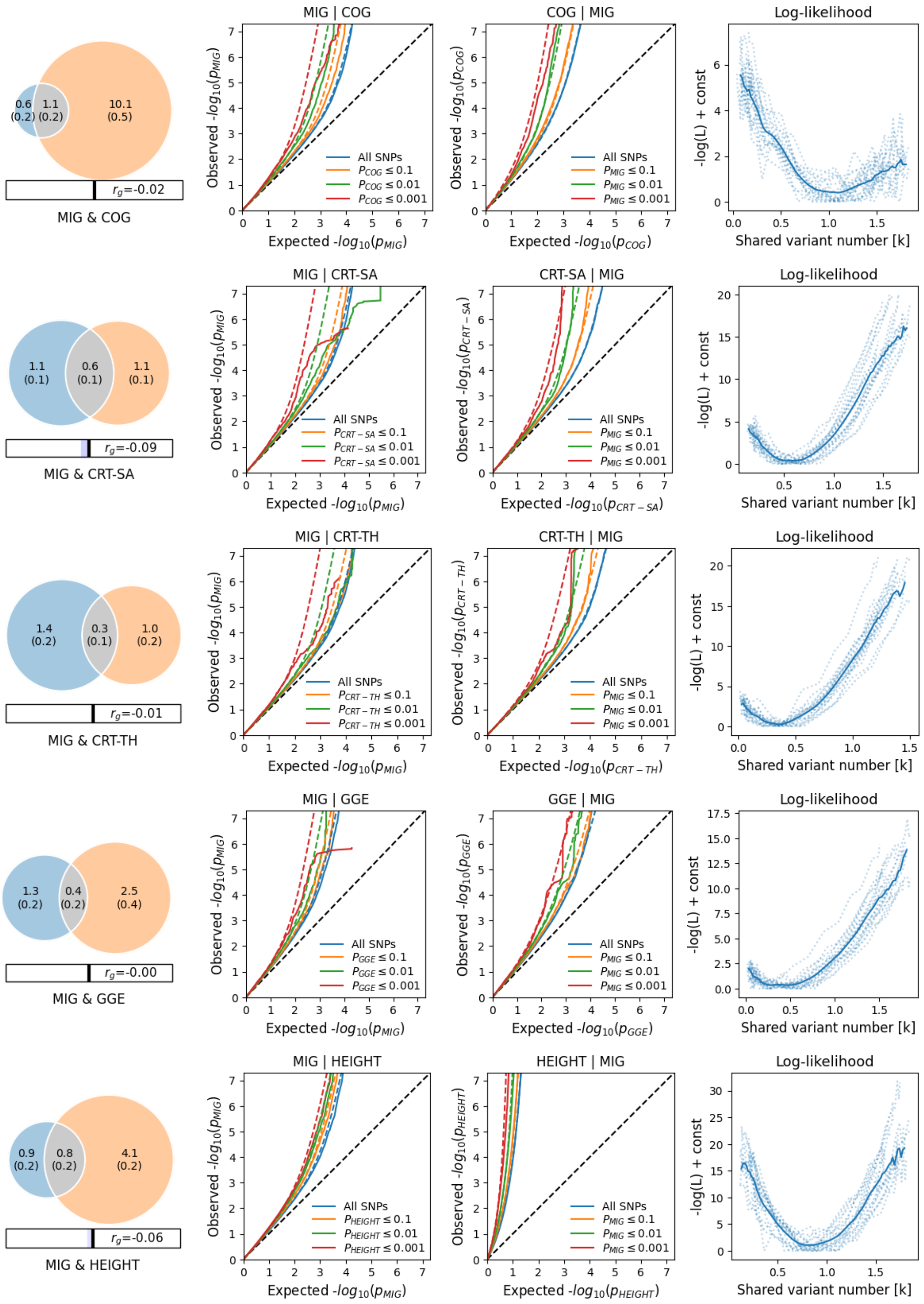
0.6 (0.2)
1.1 (0.2)
10.1 (0.5)
$r_g$=-0.02
MIG & COG
MIG | COG
COG | MIG
Log-likelihood
1.1 (0.1)
0.6 (0.1)
1.1 (0.1)
$r_g$=-0.09
MIG & CRT-SA
MIG | CRT-SA
CRT-SA | MIG
Log-likelihood
1.4 (0.2)
0.3 (0.1)
1.0 (0.2)
$r_g$=-0.01
MIG & CRT-TH
MIG | CRT-TH
CRT-TH | MIG
Log-likelihood
1.3 (0.2)
0.4 (0.2)
2.5 (0.4)
$r_g$=-0.00
MIG & GGE
MIG | GGE
GGE | MIG
Log-likelihood
0.9 (0.2)
0.8 (0.2)
4.1 (0.2)
$r_g$=-0.06
MIG & HEIGHT
MIG | HEIGHT
HEIGHT | MIG
Log-likelihood
All SNPs
Observed -$log_{10}(p)$
Expected -$log_{10}(p)$
-log(L) + const
Shared variant number [k]

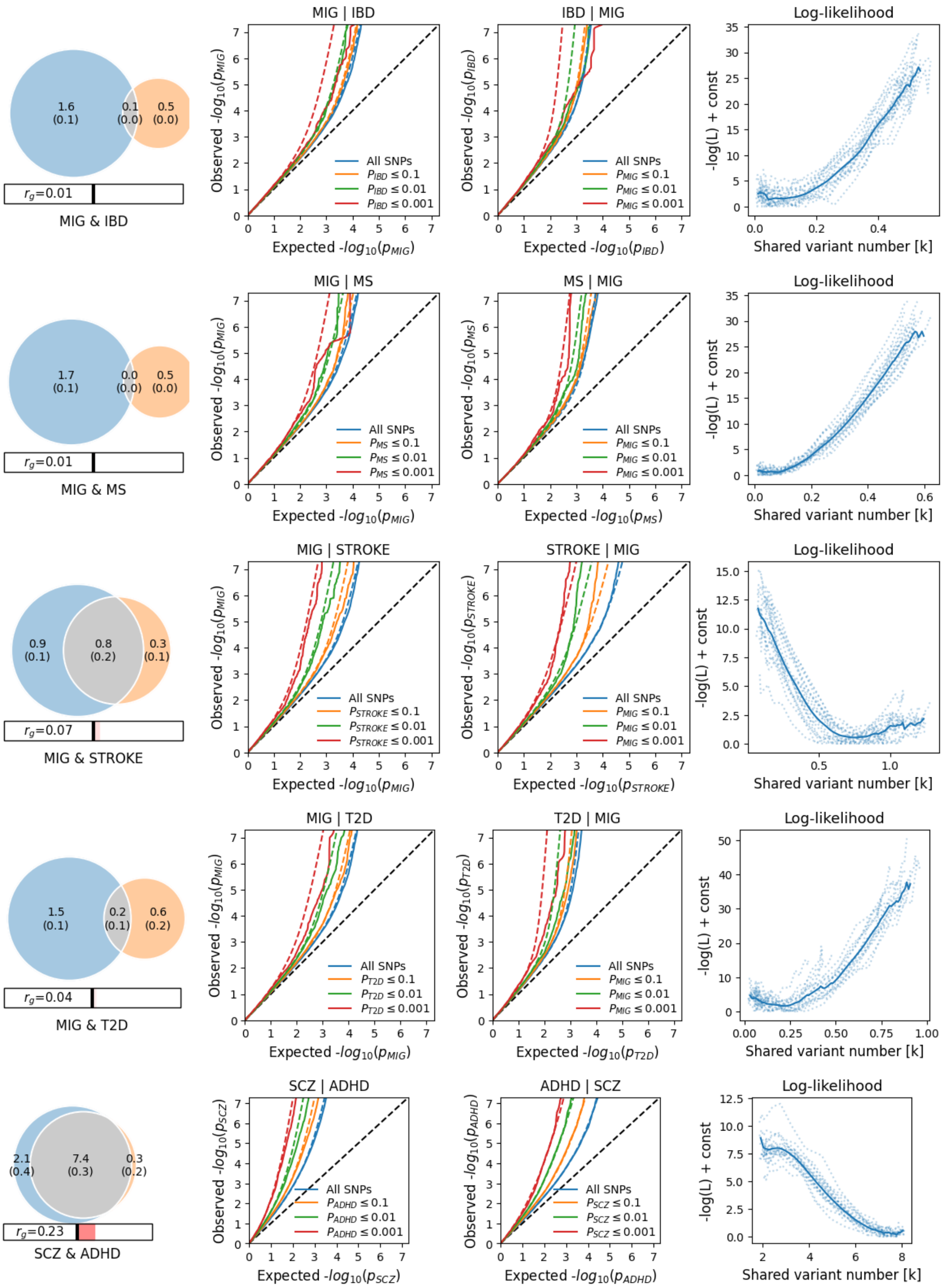
1.6 (0.1)
0.1 (0.0)
0.5 (0.0)
$r_g$=0.01
MIG & IBD
MIG | IBD
Observed -$log_{10}(p_{MIG})$
Expected -$log_{10}(p_{MIG})$
All SNPs
$P_{IBD} \leq 0.1$
$P_{IBD} \leq 0.01$
$P_{IBD} \leq 0.001$
IBD | MIG
Observed -$log_{10}(p_{IBD})$
Expected -$log_{10}(p_{IBD})$
All SNPs
$P_{MIG} \leq 0.1$
$P_{MIG} \leq 0.01$
$P_{MIG} \leq 0.001$
Log-likelihood
-log(L) + const
Shared variant number [k]
1.7 (0.1)
0.0 (0.0)
0.5 (0.0)
$r_g$=0.01
MIG & MS
MIG | MS
Observed -$log_{10}(p_{MIG})$
Expected -$log_{10}(p_{MIG})$
All SNPs
$P_{MS} \leq 0.1$
$P_{MS} \leq 0.01$
$P_{MS} \leq 0.001$
MS | MIG
Observed -$log_{10}(p_{MS})$
Expected -$log_{10}(p_{MS})$
All SNPs
$P_{MIG} \leq 0.1$
$P_{MIG} \leq 0.01$
$P_{MIG} \leq 0.001$
Log-likelihood
-log(L) + const
Shared variant number [k]
0.9 (0.1)
0.8 (0.2)
0.3 (0.1)
$r_g$=0.07
MIG & STROKE
MIG | STROKE
Observed -$log_{10}(p_{MIG})$
Expected -$log_{10}(p_{MIG})$
All SNPs
$P_{STROKE} \leq 0.1$
$P_{STROKE} \leq 0.01$
$P_{STROKE} \leq 0.001$
STROKE | MIG
Observed -$log_{10}(p_{STROKE})$
Expected -$log_{10}(p_{STROKE})$
All SNPs
$P_{MIG} \leq 0.1$
$P_{MIG} \leq 0.01$
$P_{MIG} \leq 0.001$
Log-likelihood
-log(L) + const
Shared variant number [k]
1.5 (0.1)
0.2 (0.1)
0.6 (0.2)
$r_g$=0.04
MIG & T2D
MIG | T2D
Observed -$log_{10}(p_{MIG})$
Expected -$log_{10}(p_{MIG})$
All SNPs
$P_{T2D} \leq 0.1$
$P_{T2D} \leq 0.01$
$P_{T2D} \leq 0.001$
T2D | MIG
Observed -$log_{10}(p_{T2D})$
Expected -$log_{10}(p_{T2D})$
All SNPs
$P_{MIG} \leq 0.1$
$P_{MIG} \leq 0.01$
$P_{MIG} \leq 0.001$
Log-likelihood
-log(L) + const
Shared variant number [k]
2.1 (0.4)
7.4 (0.3)
0.3 (0.2)
$r_g$=0.23
SCZ & ADHD
SCZ | ADHD
Observed -$log_{10}(p_{SCZ})$
Expected -$log_{10}(p_{SCZ})$
All SNPs
$P_{ADHD} \leq 0.1$
$P_{ADHD} \leq 0.01$
$P_{ADHD} \leq 0.001$
ADHD | SCZ
Observed -$log_{10}(p_{ADHD})$
Expected -$log_{10}(p_{ADHD})$
All SNPs
$P_{SCZ} \leq 0.1$
$P_{SCZ} \leq 0.01$
$P_{SCZ} \leq 0.001$
Log-likelihood
-log(L) + const
Shared variant number [k]

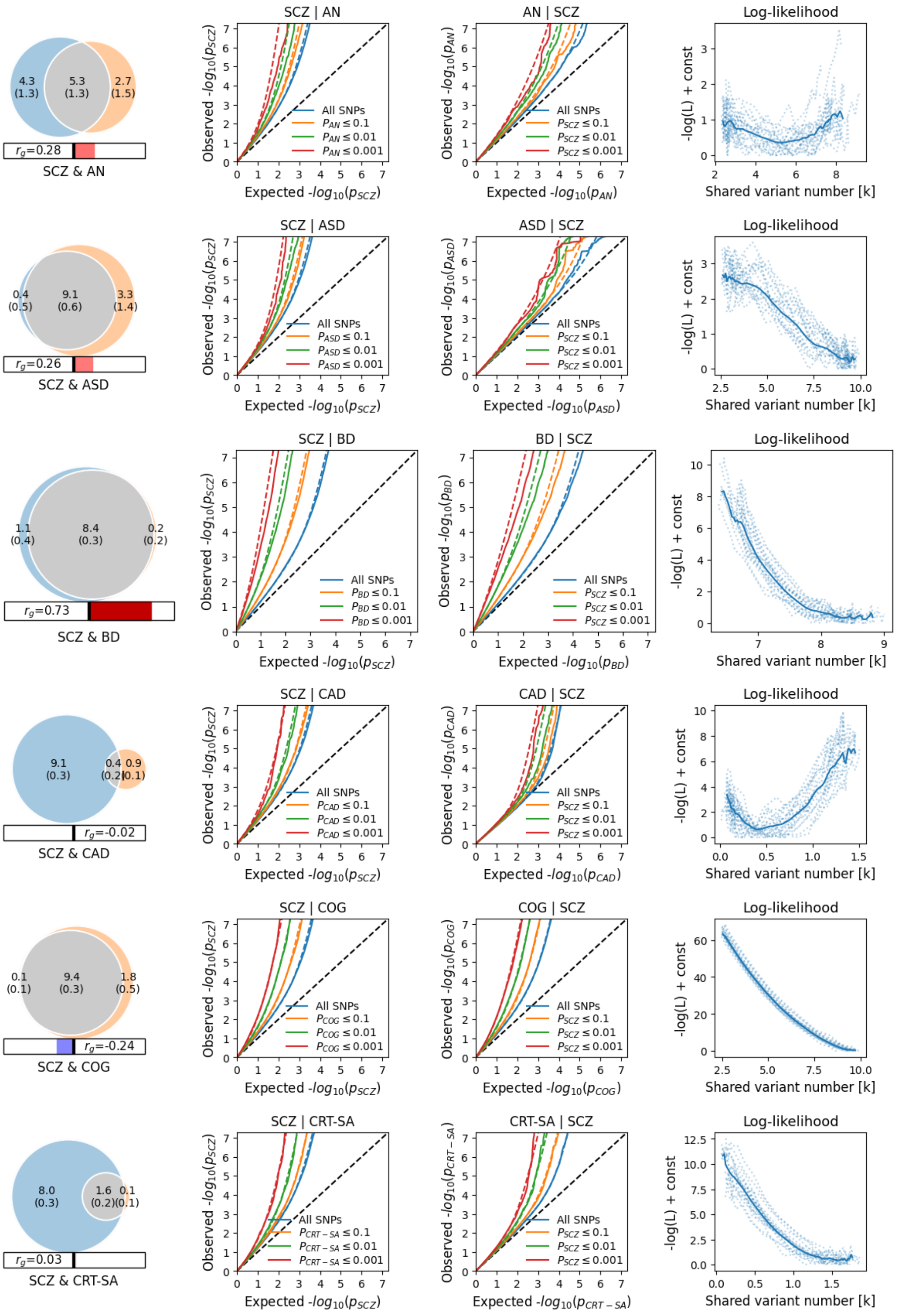

4.3 (1.3)
5.3 (1.3)
2.7 (1.5)
$r_g$=0.28
SCZ & AN
SCZ | AN
AN | SCZ
Log-likelihood
Observed $-log_{10}(p_{SCZ})$
Expected $-log_{10}(p_{SCZ})$
Observed $-log_{10}(p_{AN})$
Expected $-log_{10}(p_{AN})$
All SNPs
$P_{AN} \leq 0.1$
$P_{AN} \leq 0.01$
$P_{AN} \leq 0.001$
$P_{SCZ} \leq 0.1$
$P_{SCZ} \leq 0.01$
$P_{SCZ} \leq 0.001$
-log(L) + const
Shared variant number [k]
0.4 (0.5)
9.1 (0.6)
3.3 (1.4)
$r_g$=0.26
SCZ & ASD
SCZ | ASD
ASD | SCZ
Observed $-log_{10}(p_{ASD})$
Expected $-log_{10}(p_{ASD})$
$P_{ASD} \leq 0.1$
$P_{ASD} \leq 0.01$
$P_{ASD} \leq 0.001$
1.1 (0.4)
8.4 (0.3)
0.2 (0.2)
$r_g$=0.73
SCZ & BD
SCZ | BD
BD | SCZ
Observed $-log_{10}(p_{BD})$
Expected $-log_{10}(p_{BD})$
$P_{BD} \leq 0.1$
$P_{BD} \leq 0.01$
$P_{BD} \leq 0.001$
9.1 (0.3)
0.4 (0.2)
0.9 (0.1)
$r_g$=-0.02
SCZ & CAD
SCZ | CAD
CAD | SCZ
Observed $-log_{10}(p_{CAD})$
Expected $-log_{10}(p_{CAD})$
$P_{CAD} \leq 0.1$
$P_{CAD} \leq 0.01$
$P_{CAD} \leq 0.001$
0.1 (0.1)
9.4 (0.3)
1.8 (0.5)
$r_g$=-0.24
SCZ & COG
SCZ | COG
COG | SCZ
Observed $-log_{10}(p_{COG})$
Expected $-log_{10}(p_{COG})$
$P_{COG} \leq 0.1$
$P_{COG} \leq 0.01$
$P_{COG} \leq 0.001$
8.0 (0.3)
1.6 (0.2)
0.1 (0.1)
$r_g$=0.03
SCZ & CRT-SA
SCZ | CRT-SA
CRT-SA | SCZ
Observed $-log_{10}(p_{CRT-SA})$
Expected $-log_{10}(p_{CRT-SA})$
$P_{CRT-SA} \leq 0.1$
$P_{CRT-SA} \leq 0.01$
$P_{CRT-SA} \leq 0.001$

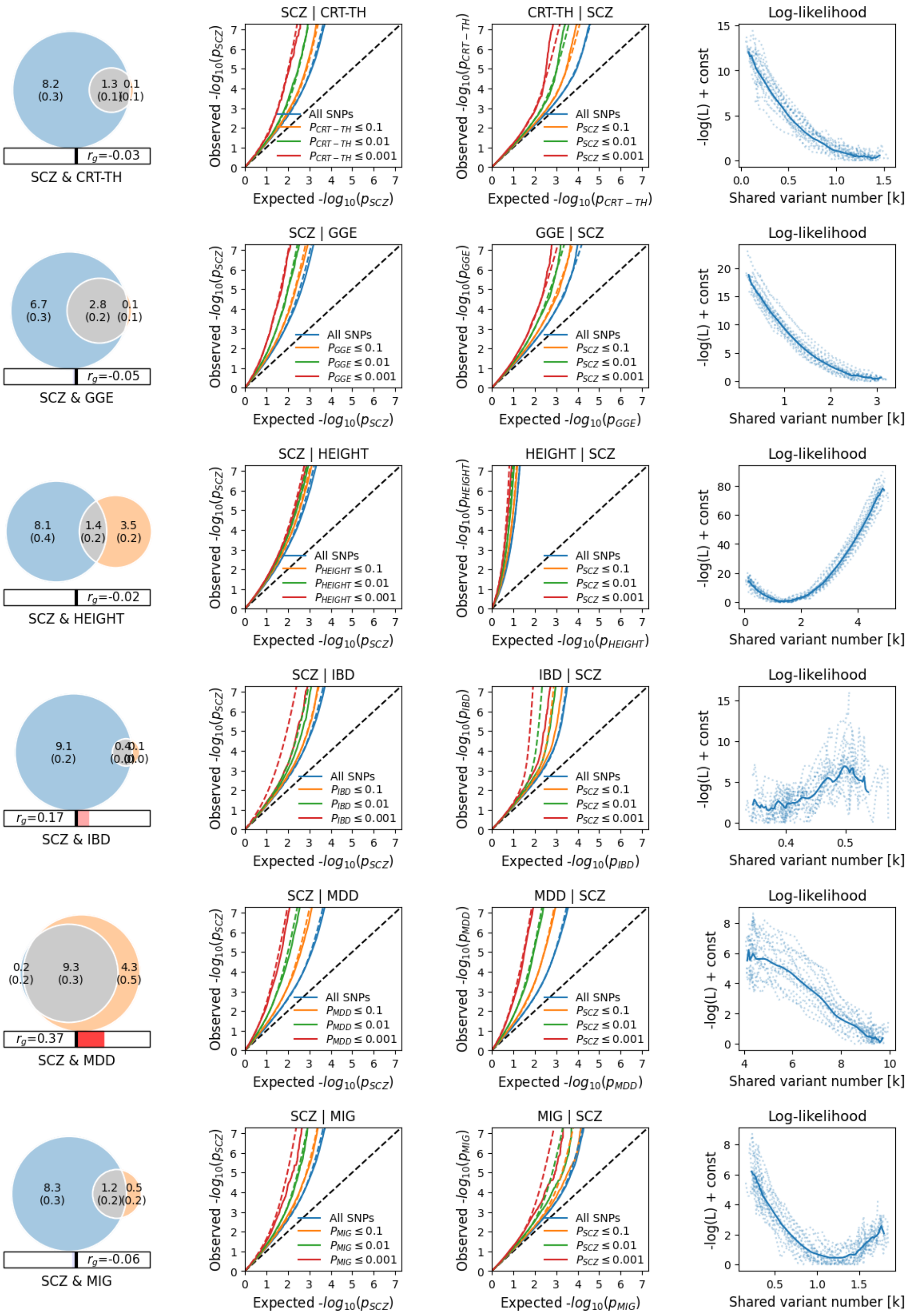
SCZ & CRT-TH
SCZ | CRT-TH
CRT-TH | SCZ
Log-likelihood
SCZ & GGE
SCZ | GGE
GGE | SCZ
SCZ & HEIGHT
SCZ | HEIGHT
HEIGHT | SCZ
SCZ & IBD
SCZ | IBD
IBD | SCZ
SCZ & MDD
SCZ | MDD
MDD | SCZ
SCZ & MIG
SCZ | MIG
MIG | SCZ
All SNPs
-log(L) + const
Shared variant number [k]
$r_g$=-0.03
$r_g$=-0.05
$r_g$=-0.02
$r_g$=0.17
$r_g$=0.37
$r_g$=-0.06

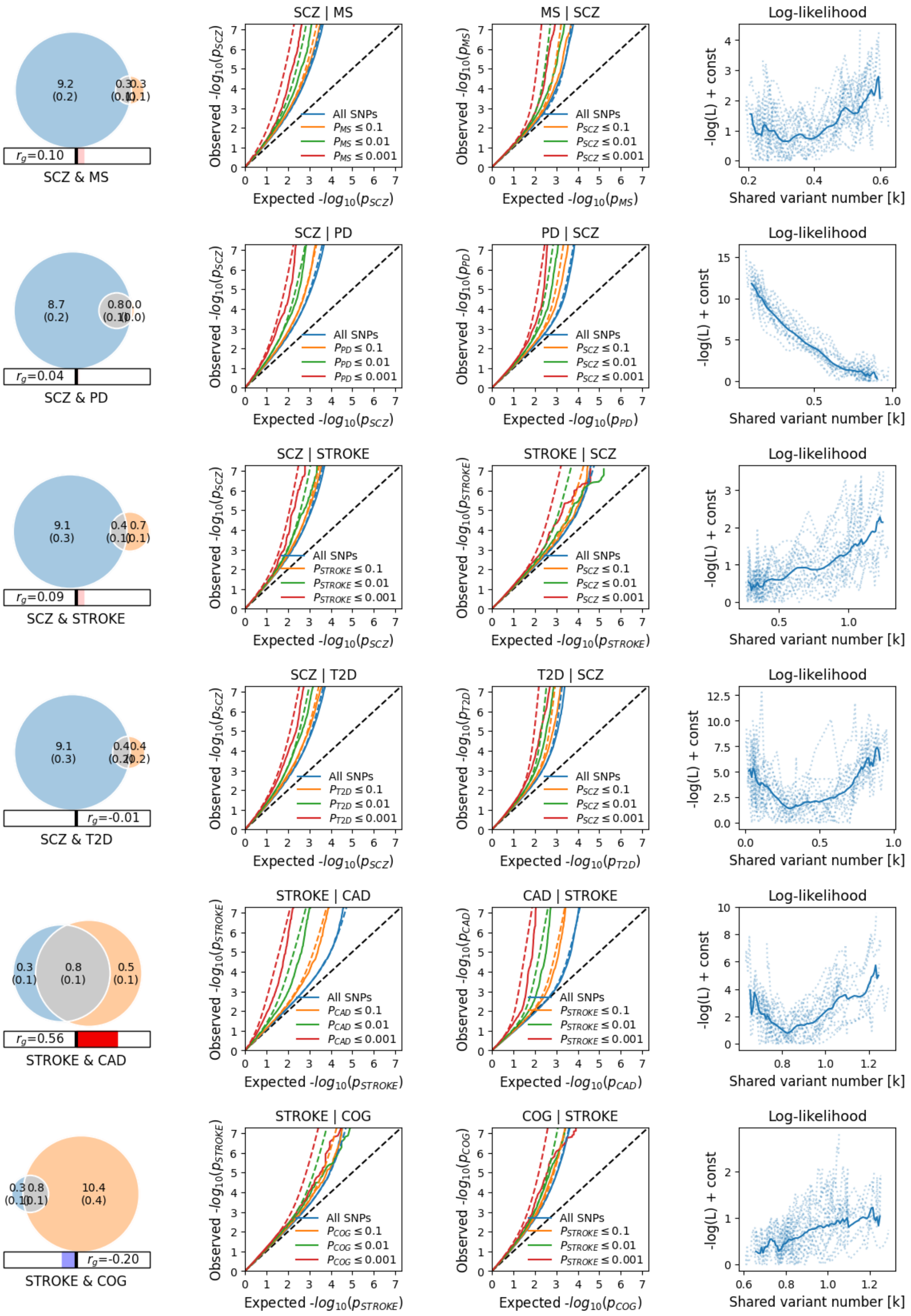
SCZ & MS
$r_g$=0.10
SCZ | MS
MS | SCZ
Log-likelihood
SCZ & PD
$r_g$=0.04
SCZ | PD
PD | SCZ
Log-likelihood
SCZ & STROKE
$r_g$=0.09
SCZ | STROKE
STROKE | SCZ
Log-likelihood
SCZ & T2D
$r_g$=-0.01
SCZ | T2D
T2D | SCZ
Log-likelihood
STROKE & CAD
$r_g$=0.56
STROKE | CAD
CAD | STROKE
Log-likelihood
STROKE & COG
$r_g$=-0.20
STROKE | COG
COG | STROKE
Log-likelihood
All SNPs
Shared variant number [k]
-log(L) + const

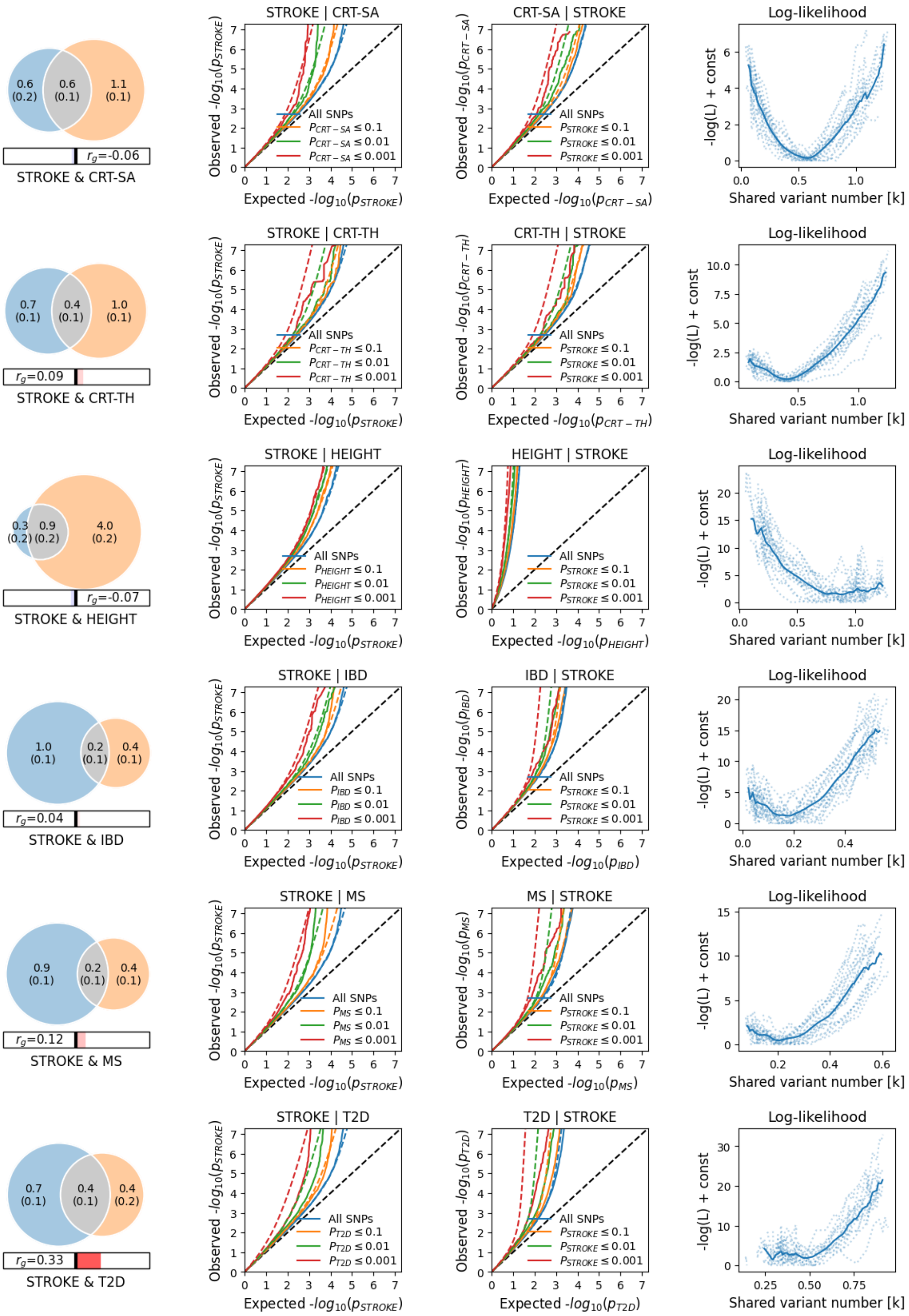

0.6 (0.2)
0.6 (0.1)
1.1 (0.1)
$r_g$=-0.06
STROKE & CRT-SA
STROKE | CRT-SA
CRT-SA | STROKE
Log-likelihood
0.7 (0.1)
0.4 (0.1)
1.0 (0.1)
$r_g$=0.09
STROKE & CRT-TH
STROKE | CRT-TH
CRT-TH | STROKE
Log-likelihood
0.3 (0.2)
0.9 (0.2)
4.0 (0.2)
$r_g$=-0.07
STROKE & HEIGHT
STROKE | HEIGHT
HEIGHT | STROKE
Log-likelihood
1.0 (0.1)
0.2 (0.1)
0.4 (0.1)
$r_g$=0.04
STROKE & IBD
STROKE | IBD
IBD | STROKE
Log-likelihood
0.9 (0.1)
0.2 (0.1)
0.4 (0.1)
$r_g$=0.12
STROKE & MS
STROKE | MS
MS | STROKE
Log-likelihood
0.7 (0.1)
0.4 (0.1)
0.4 (0.2)
$r_g$=0.33
STROKE & T2D
STROKE | T2D
T2D | STROKE
Log-likelihood
All SNPs
$p \leq 0.1$
$p \leq 0.01$
$p \leq 0.001$
Observed -$log_{10}(p)$
Expected -$log_{10}(p)$
-log(L) + const
Shared variant number [k]

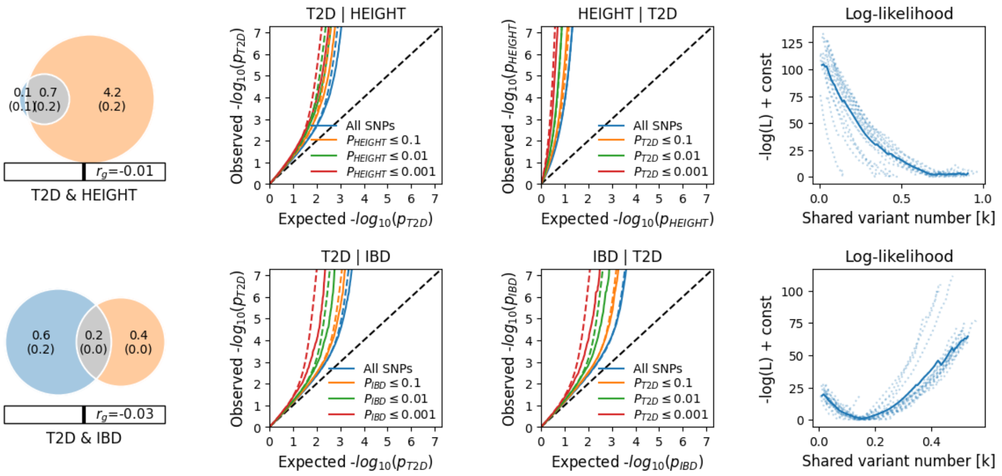

**Supplementary Fig. 3 |** Bivariate MiXeR estimates of genetic overlap. For each pair of phenotypes, the unique and overlapping genetic architectures estimated using MiXeR47, excluding phenotypes with poor model fit due to insufficient GWAS power (ANX, PTSD, TS, OCD, FE, ET and LBD). MiXeR-modeled Venn diagrams displaying the number (in thousands) of shared and phenotype-specific "causal" variants for each pair of phenotypes, conditional QQ plots showing real-life vs modelled cross trait enrichment and log-likelihood plots, plotting the adjusted negative log-likelihood function against the number of shared 'causal' variants, constrained to minimum and maximum possible overlap.

#### 3.4 Supplementary Fig. 4 | Tissue specific expression of mapped genes

##### Alzheimer's disease

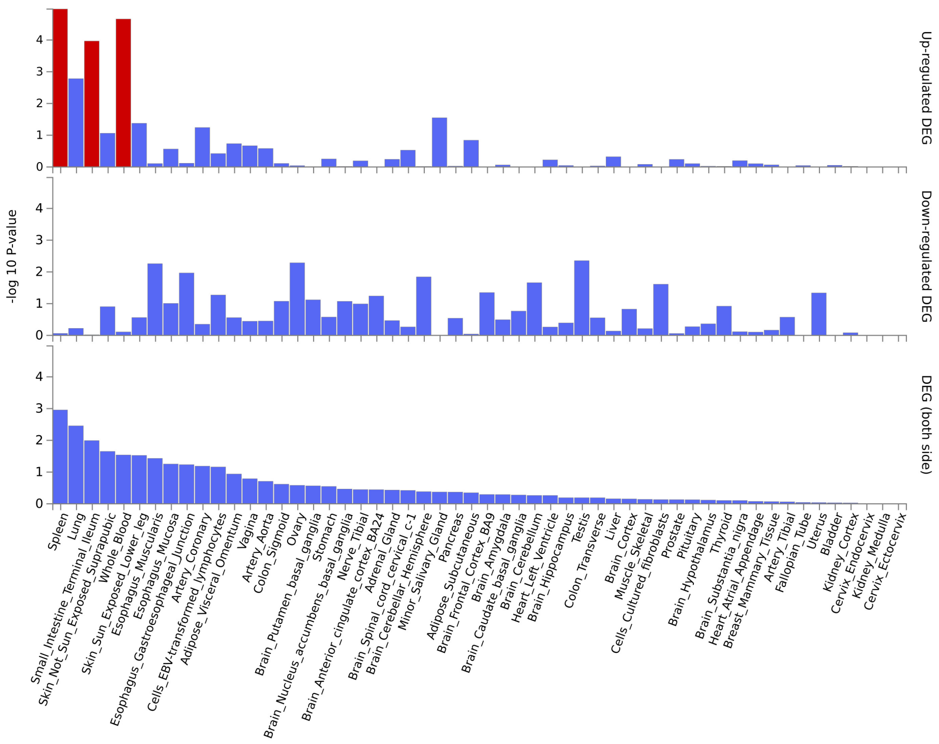

##### Attention-deficit/hyperactivity disorder

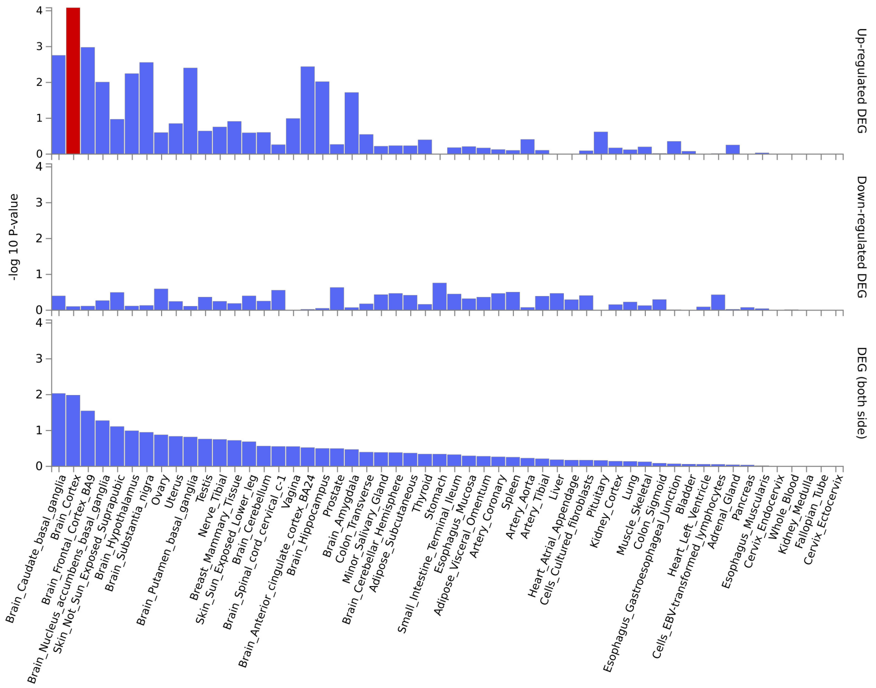

### Anorexia nervosa

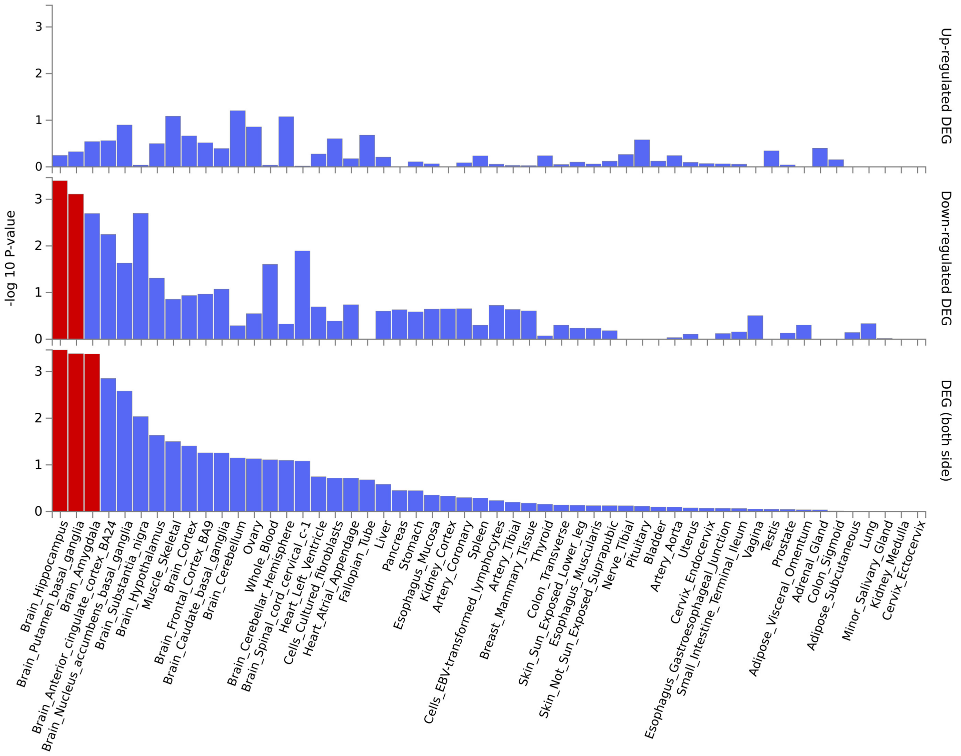

### Cortical thickness

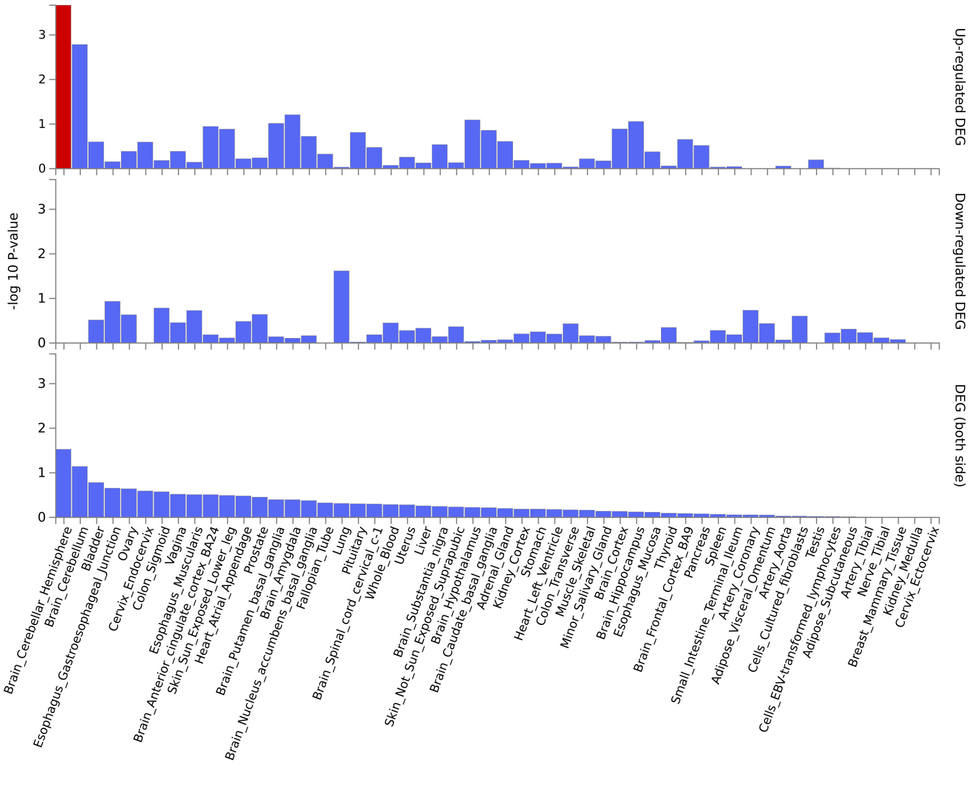

### General cognitive ability

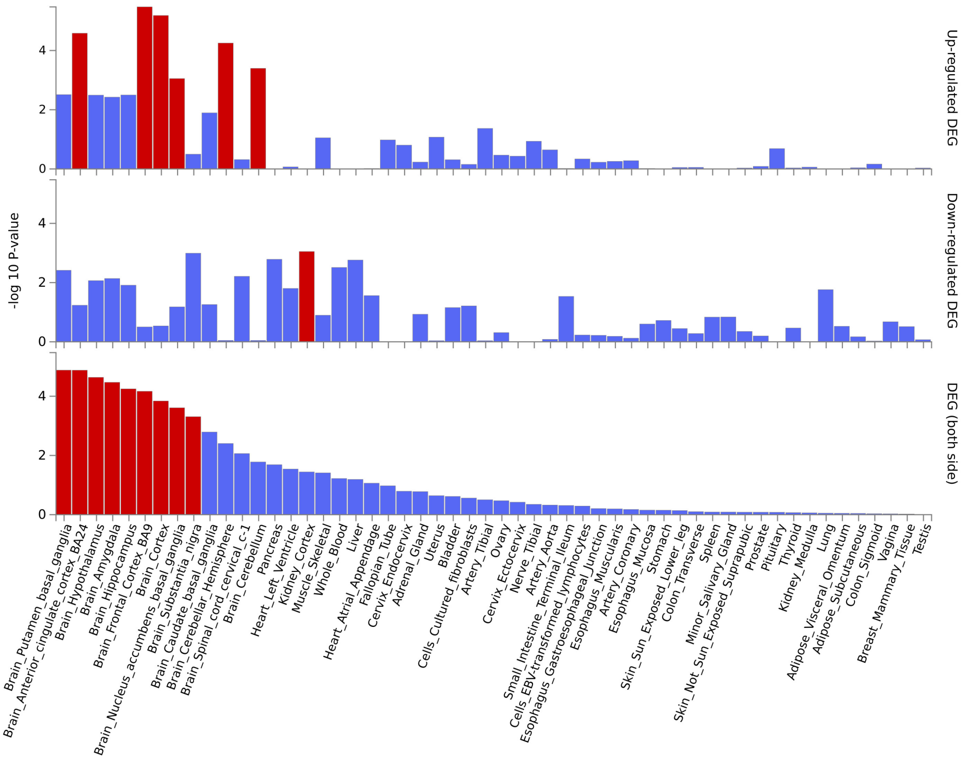

### Height

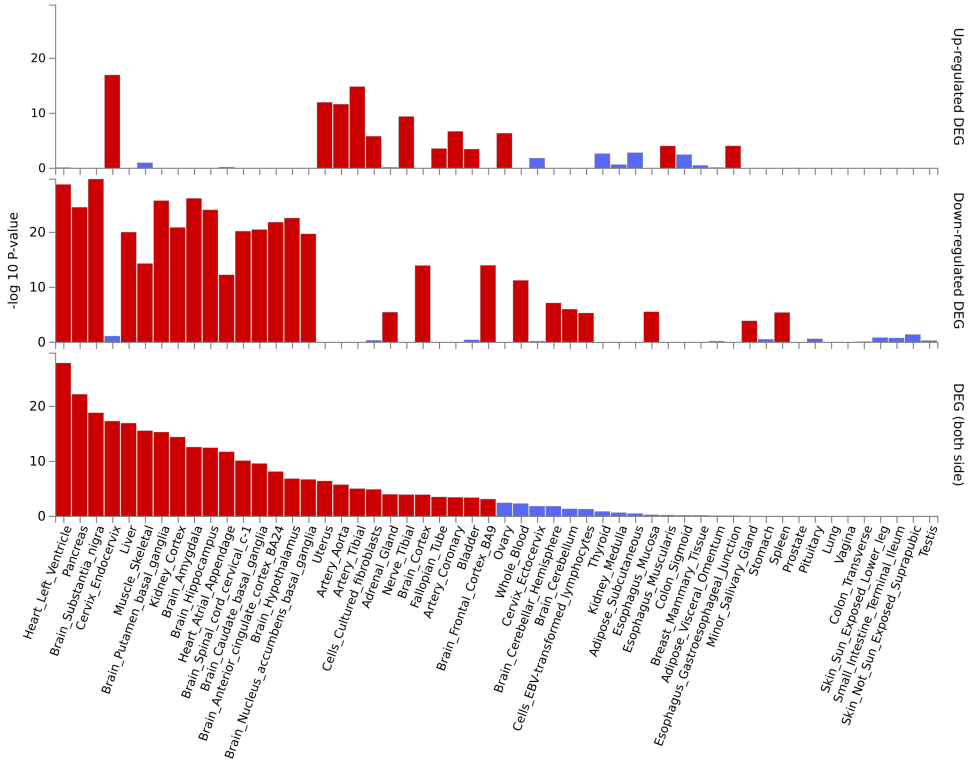

### Inflammatory bowel disease

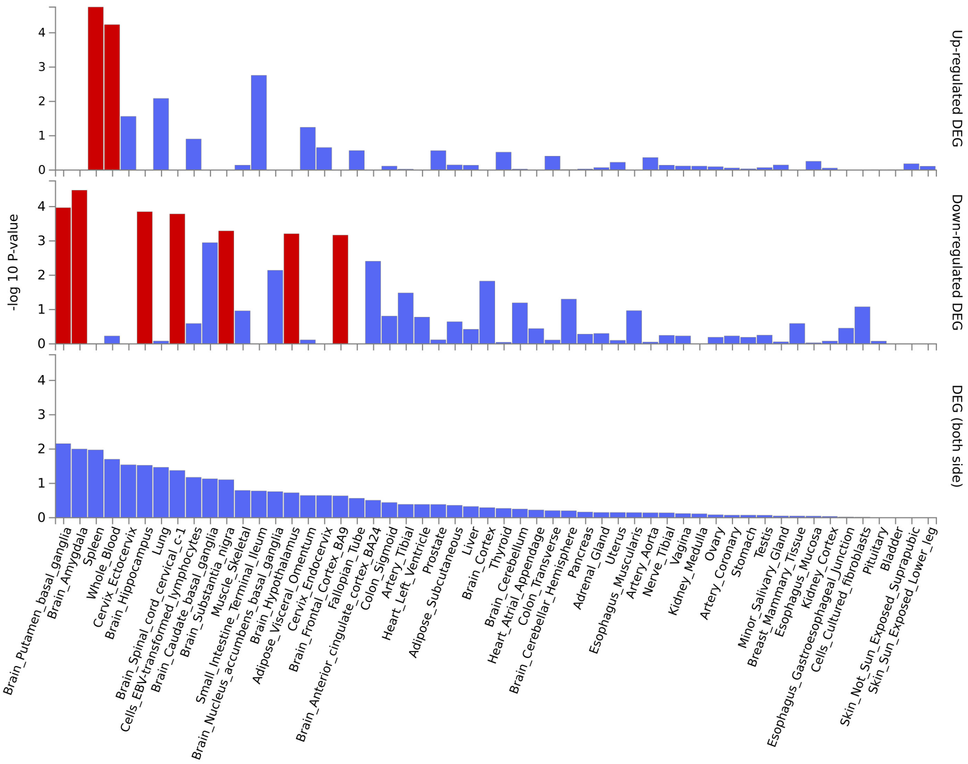

### Major depressive disorder

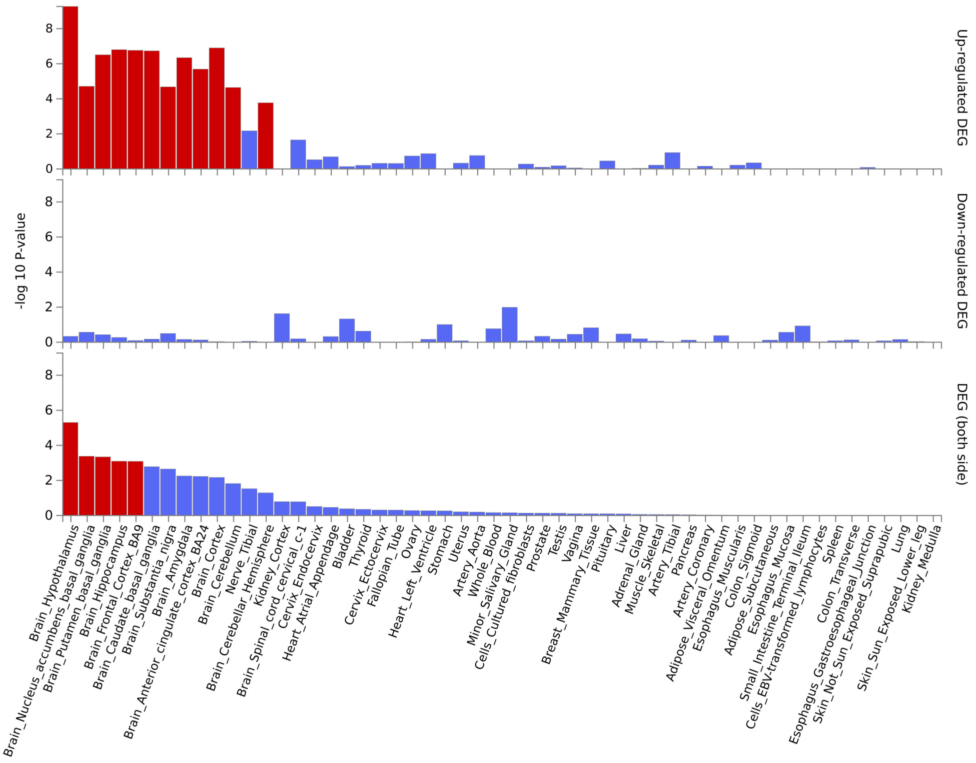

#### Multiple sclerosis

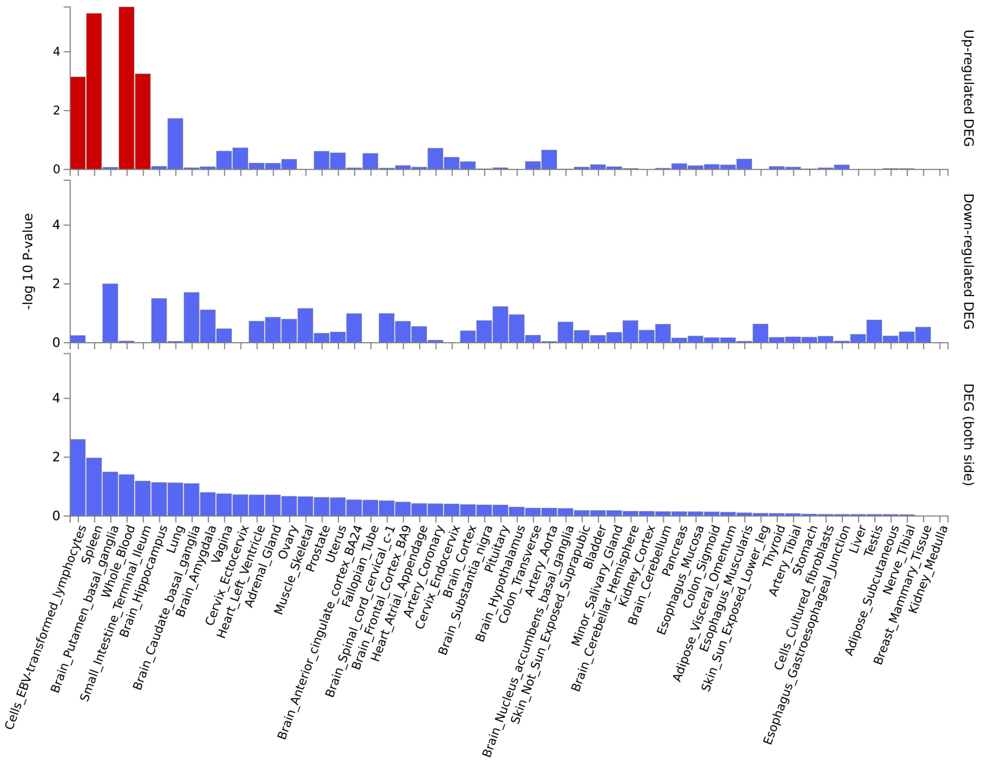

#### Parkinson's disease

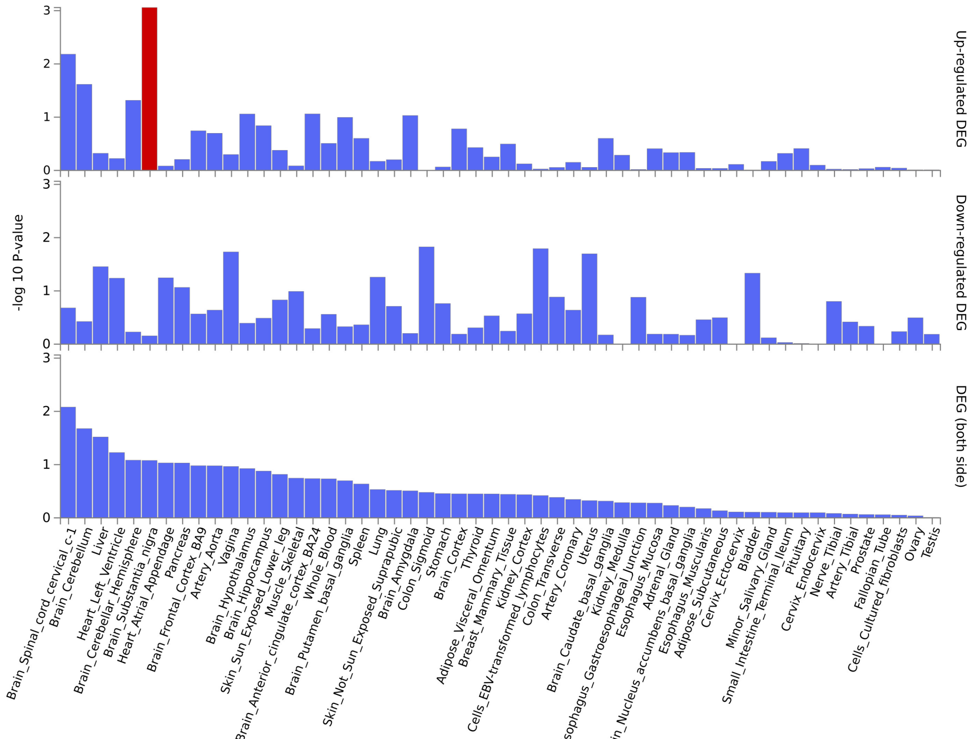

### Schizophrenia

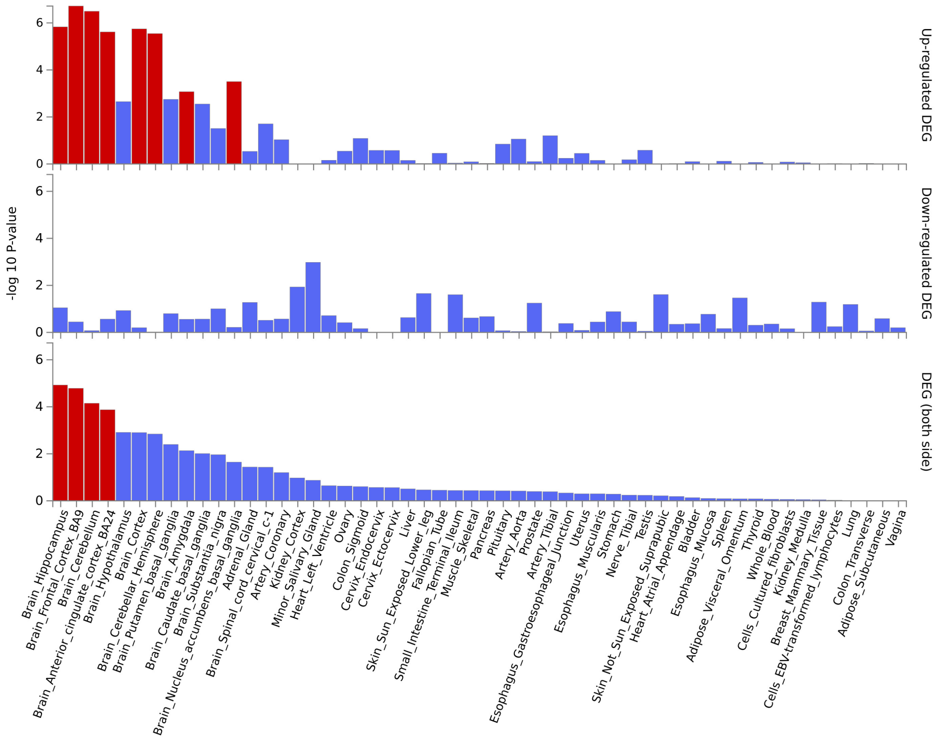

### Type 2 Diabetes

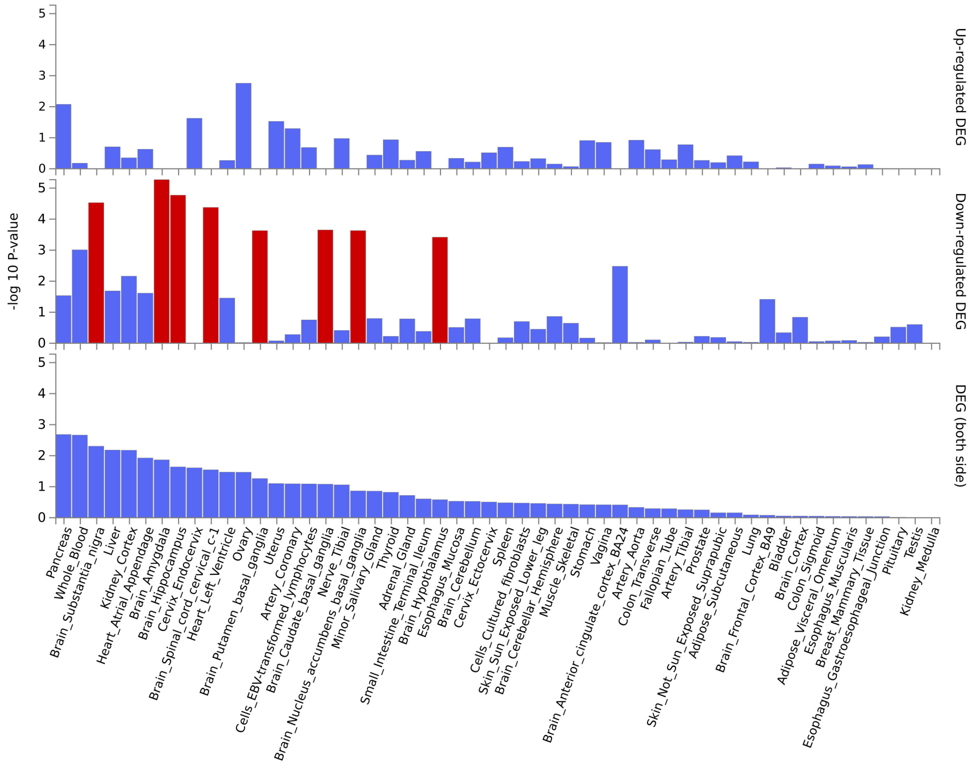

**Supplementary Fig. 4 | Tissue specific enrichment.** Results from MAGMA tissue specificity analysis across 54 different tissue types based on GTEx v.8 data[44], as implemented in FUMA[45]. Red bars indicate significant results after Bonferroni correction..

#### 3.5 Supplementary Fig. 5 | Cell type specific expression of mapped genes

##### Alzheimer's disease

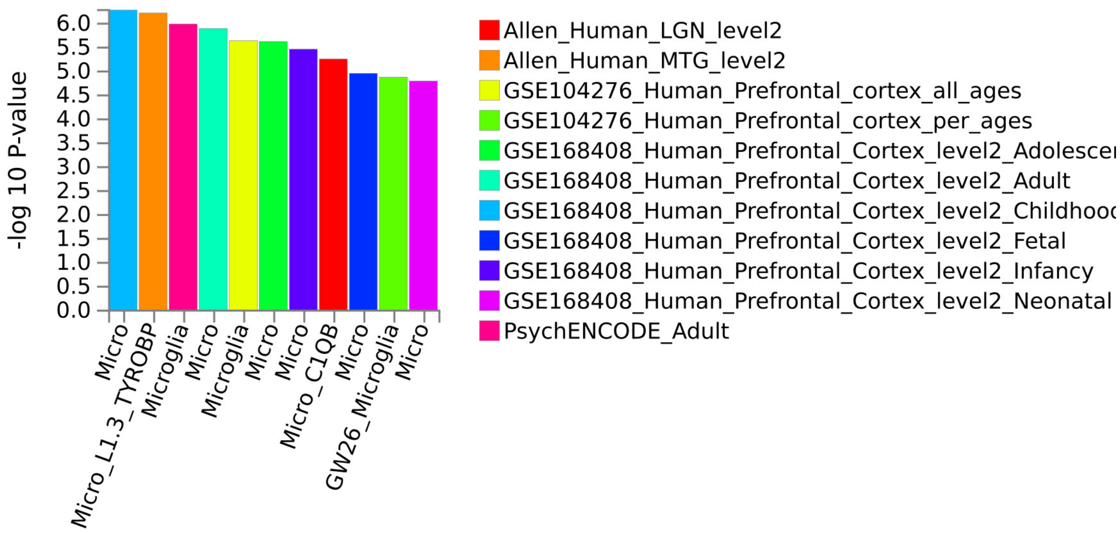

##### Genetic generalized epilepsy

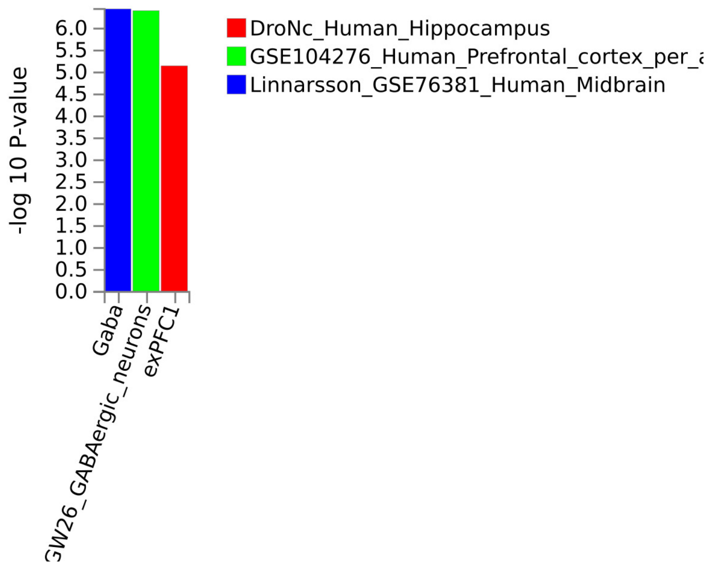

**Multiple sclerosis**

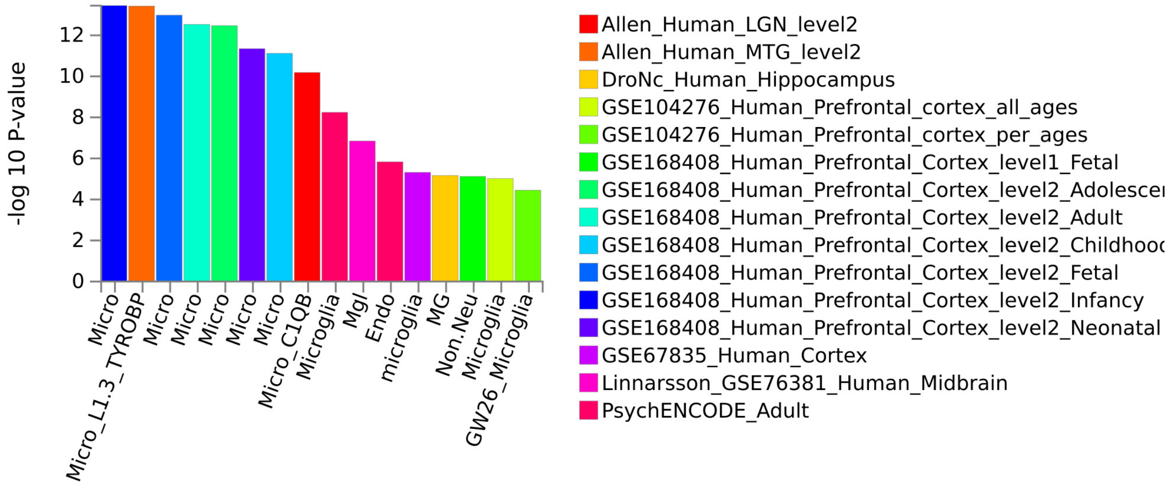

**Attention-deficit/hyperactivity disorder**

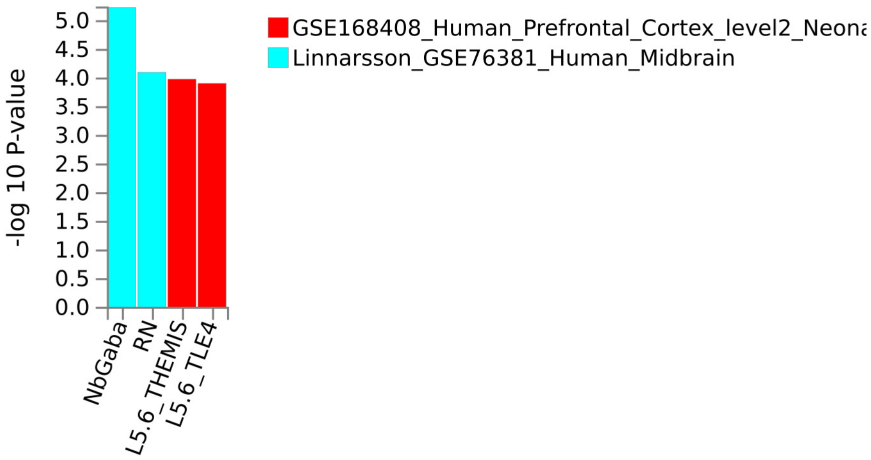

#### Bipolar disorder

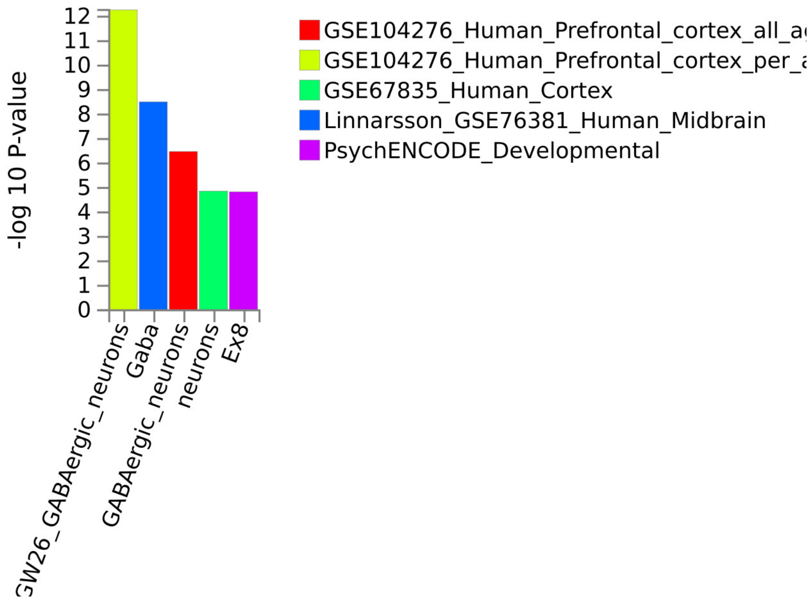

#### Major depressive disorder

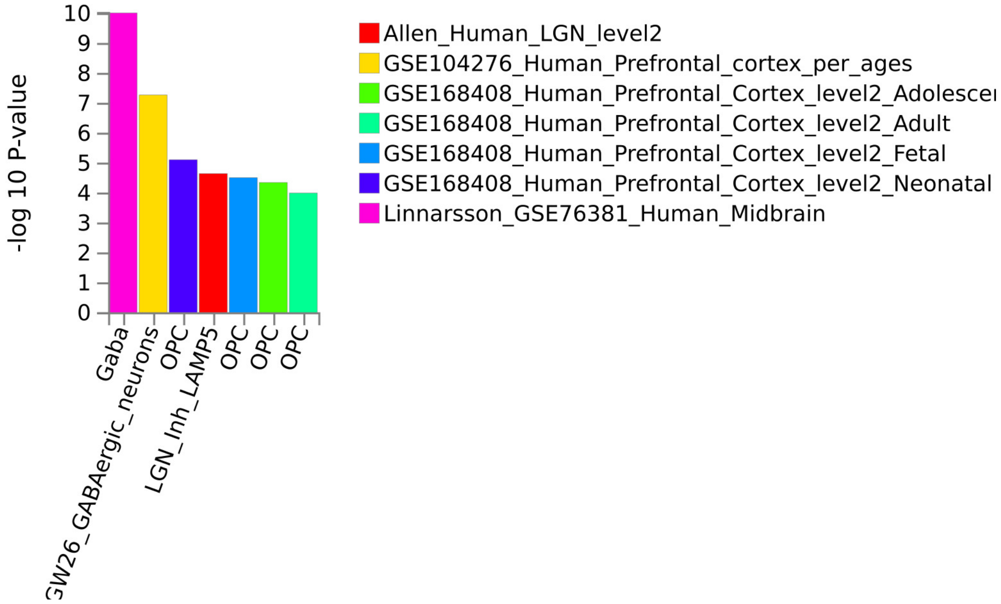

#### Schizophrenia

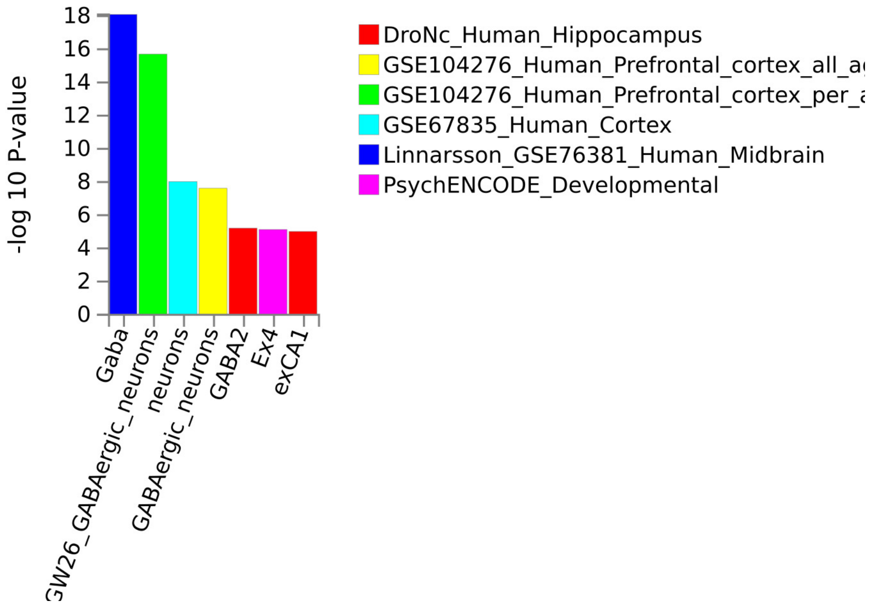

#### Coronary artery disease

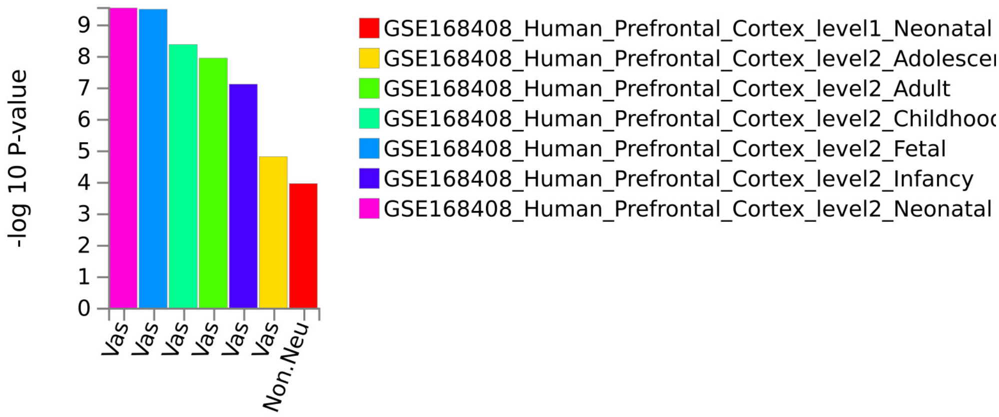

#### Cortical surface area

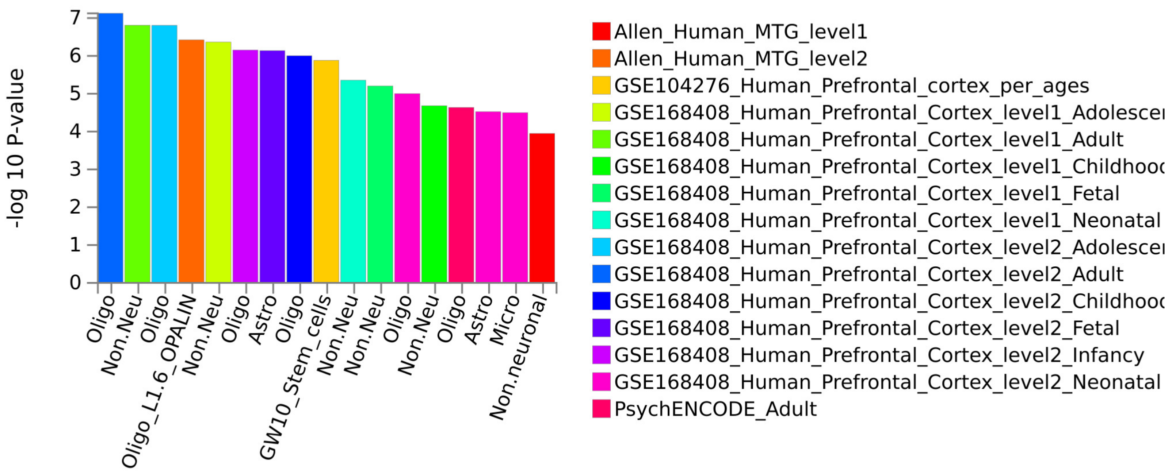

#### General cognitive ability

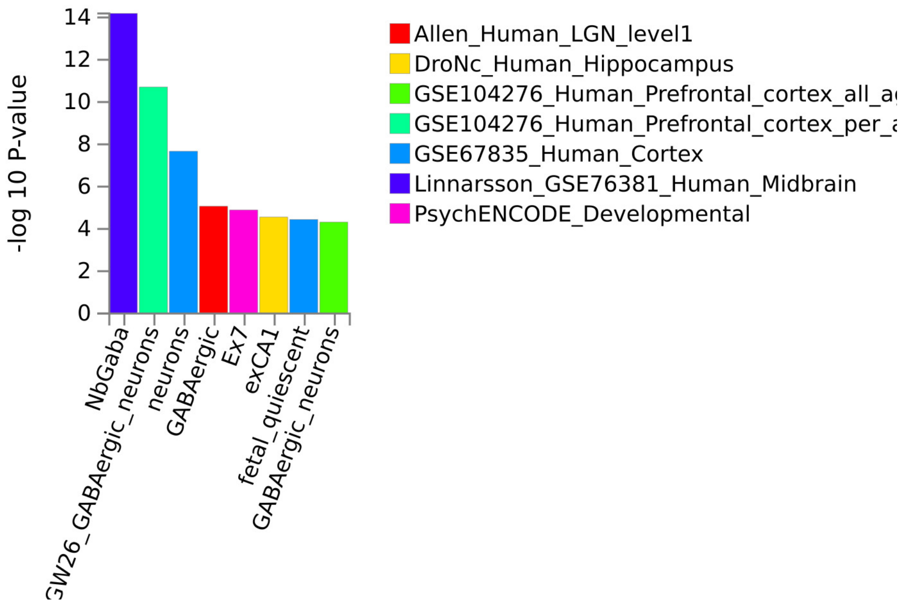

#### Inflammatory bowel disease

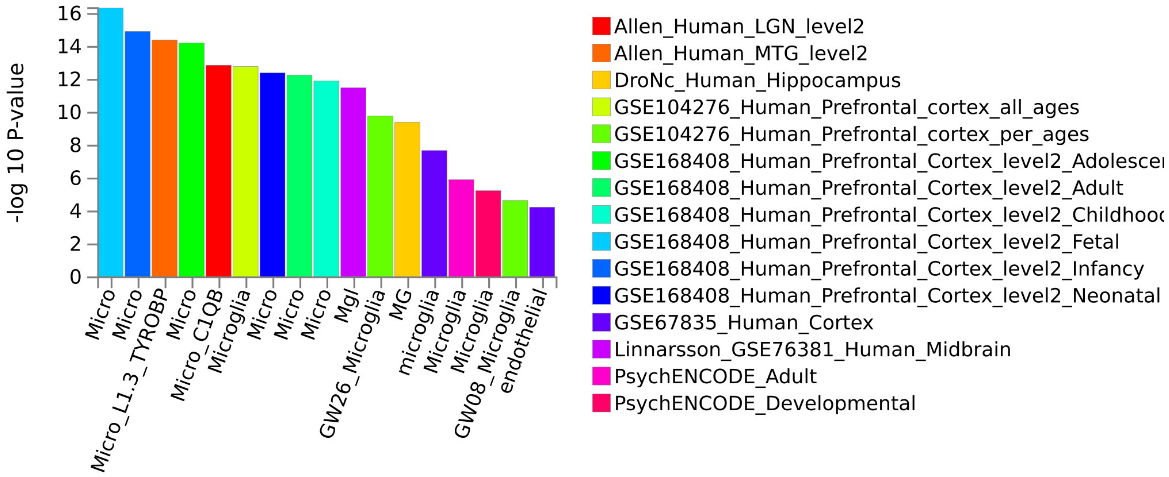

#### Type 2 diabetes

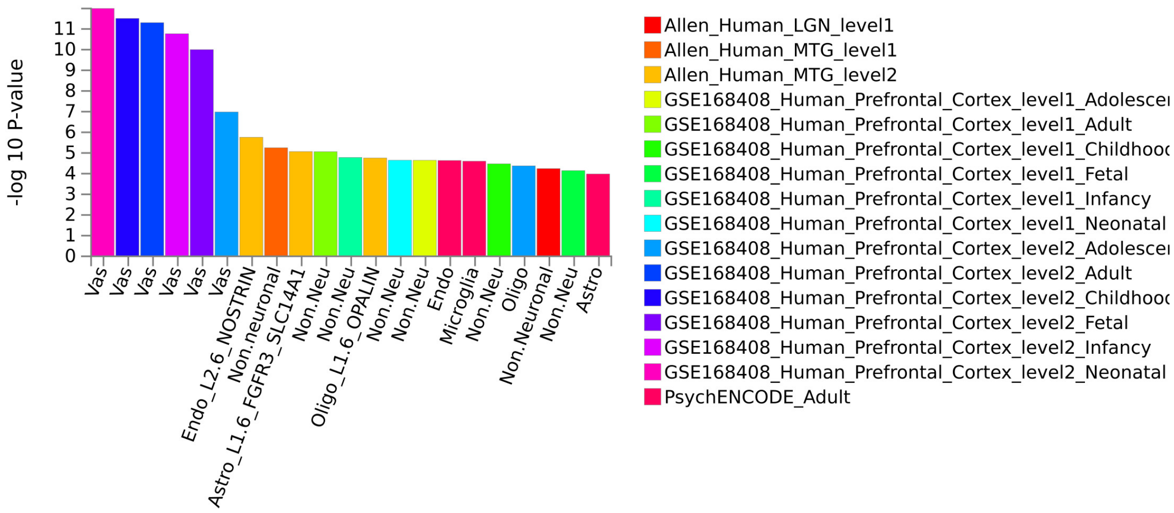

**Height**

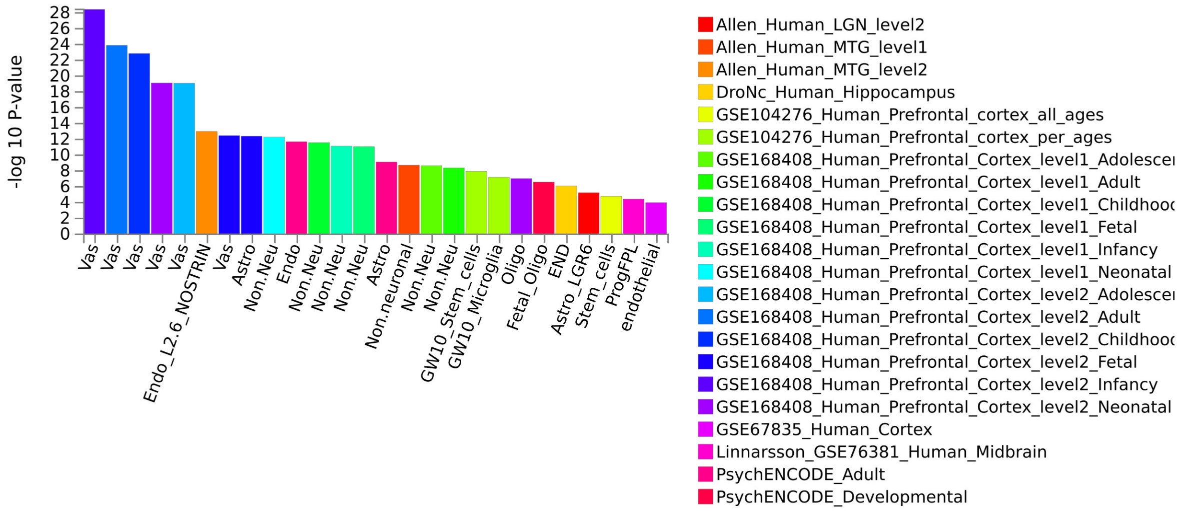

**Supplementary Fig. 5 | Cell type specificity analysis**. Results from MAGMA cell type specificity analysis using 24 single-cell RNA sequencing datasets from the developing and adult human brain, as implemented in FUMA[48]. Significant cell type associations were determined after Bonferroni correction, followed by conditional analysis within each dataset to account for correlated cell type expression profiles.
